# Geographic variation of mutagenic exposures in kidney cancer genomes

**DOI:** 10.1101/2023.06.20.23291538

**Authors:** Sergey Senkin, Sarah Moody, Marcos Díaz-Gay, Behnoush Abedi-Ardekani, Thomas Cattiaux, Aida Ferreiro-Iglesias, Jingwei Wang, Stephen Fitzgerald, Mariya Kazachkova, Raviteja Vangara, Anh Phuong Le, Erik N. Bergstrom, Azhar Khandekar, Burçak Otlu, Saamin Cheema, Calli Latimer, Emily Thomas, Joshua Ronald Atkins, Karl Smith-Byrne, Ricardo Cortez Cardoso Penha, Christine Carreira, Priscilia Chopard, Valérie Gaborieau, Pekka Keski-Rahkonen, David Jones, Jon W. Teague, Sophie Ferlicot, Mojgan Asgari, Surasak Sangkhathat, Worapat Attawettayanon, Beata Świątkowska, Sonata Jarmalaite, Rasa Sabaliauskaite, Tatsuhiro Shibata, Akihiko Fukagawa, Dana Mates, Viorel Jinga, Stefan Rascu, Mirjana Mijuskovic, Slavisa Savic, Sasa Milosavljevic, John M.S. Bartlett, Monique Albert, Larry Phouthavongsy, Patricia Ashton-Prolla, Mariana R. Botton, Brasil Silva Neto, Stephania Martins Bezerra, Maria Paula Curado, Stênio de Cássio Zequi, Rui Manuel Reis, Eliney Faria, Nei Soares Menezes, Renata Spagnoli Ferrari, Rosamonde E. Banks, Naveen S. Vasudev, David Zaridze, Anush Mukeriya, Oxana Shangina, Vsevolod Matveev, Lenka Foretova, Marie Navratilova, Ivana Holcatova, Anna Hornakova, Vladimir Janout, Mark Purdue, Nathaniel Rothman, Stephen J. Chanock, Per Magne Ueland, Mattias Johansson, James McKay, Ghislaine Scelo, Estelle Chanudet, Laura Humphreys, Ana Carolina de Carvalho, Sandra Perdomo, Ludmil B. Alexandrov, Michael R. Stratton, Paul Brennan

**Affiliations:** Genomic Epidemiology Branch, International Agency for Research on Cancer (IARC/WHO), Lyon, France; Cancer, Ageing and Somatic Mutation, Wellcome Sanger Institute, Cambridge, UK; Department of Cellular and Molecular Medicine, University of California San Diego, La Jolla, USA; Department of Bioengineering, University of California San Diego, La Jolla, USA; Moores Cancer Center, University of California San Diego, La Jolla, USA; Biomedical Sciences Graduate Program, University of California San Diego, La Jolla, USA; Department of Health Informatics, Graduate School of Informatics, Middle East Technical University, Ankara, Turkey; Cancer Epidemiology Unit, The Nuffield Department of Population Health, University of Oxford, Oxford, UK; Evidence Synthesis and Classification Branch, International Agency for Research on Cancer (IARC/WHO), Lyon, France; Nutrition and Metabolism Branch, International Agency for Research on Cancer (IARC/WHO), Lyon, France; Service d’Anatomie Pathologique, Assistance Publique-Hôpitaux de Paris, Univeristé Paris-Saclay, Le Kremlin-Bicêtre, France; Oncopathology Research Center, Iran University of Medical Sciences, Tehran, Iran; Translational Medicine Research Center, Faculty of Medicine, Prince of Songkla University, Hat Yai, Thailand; Department of Surgery, Urology, Faculty of Medicine, Prince of Songkla University, Hat Yai, Thailand; Department of Environmental Epidemiology, Nofer Institute of Occupational Medicine, Łódź, Poland; Laboratory of Genetic Diagnostic, National Cancer Institute, Vilnius, Lithuania; Department of Botany and Genetics, Institute of Biosciences, Vilnius University, Vilnius, Lithuania; Laboratory of Molecular Medicine, The Institute of Medical Science, The University of Tokyo, Minato-ku, Japan; Division of Cancer Genomics, National Cancer Center Research Institute, Chuo-ku, Japan; Department of Pathology, Graduate School of Medicine, The University of Tokyo, Bunkyo-ku, Japan; Occupational Health and Toxicology, National Center for Environmental Risk Monitoring, National Institute of Public Health, Bucharest, Romania; Urology Department, “Carol Davila” University of Medicine and Pharmacy - “Prof. Dr. Th. Burghele” Clinical Hospital, Bucharest, Romania; Clinic of Nefrology, Faculty of Medicine, Military Medical Academy, Belgrade, Serbia; Department of Urology, University Hospital “Dr D. Misovic” Clinical Center, Belgrade, Serbia; International Organization for Cancer Prevention and Research, Belgrade, Serbia; Cancer Research UK Edinburgh Centre, Institute of Genetics and Cancer, University of Edinburgh, Edinburgh, Scotland; Centre for Biodiversity Genomics, University of Guelph, Guelph, Canada; Ontario Tumour Bank, Ontario Institute for Cancer Research, Toronto, Canada; Experimental Research Center, Genomic Medicine Laboratory, Hospital de Clínicas de Porto Alegre, Porto Alegre, Brazil; Post-Graduate Program in Genetics and Molecular Biology, Universidade Federal do Rio Grande do Sul, Porto Alegre, Brazil; Diagnostic Laboratory Service, Personalized Medicine, Hospital de Clínicas de Porto Alegre, Porto Alegre, Brazil; Service of Urology, Hospital de Clínicas de Porto Alegre, Porto Alegre, Brazil; Post-Graduate Program in Medicine: Surgical Sciences, Universidade Federal do Rio Grande do Sul, Porto Alegre, Brazil; Department of Anatomic Pathology, A.C. Camargo Cancer Center, São Paulo, Brazil; Department of Epidemiology, A.C. Camargo Cancer Center, São Paulo, Brazil; Department of Urology, A.C. Camargo Cancer Center, São Paulo, Brazil; National Institute for Science and Technology in Oncogenomics and Therapeutic Innovation, A.C. Camargo Cancer Center, São Paulo, Brazil; Latin American Renal Cancer Group – LARCG, São Paulo, Brazil; Department of Surgery, Division of Urology, Sao Paulo Federal University - UNIFESP, São Paulo, Brazil; Molecular Oncology Research Center, Barretos Cancer Hospital, Brazil; Life and Health Sciences Research Institute (ICVS), School of Medicine, Minho University, Braga, Portugal; Department of Urology, Barretos Cancer Hospital, Brazil; Department of Pathology, Barretos Cancer Hospital, Brazil; Leeds Institute of Medical Research at St James’s, University of Leeds, Leeds, UK; Clinical Epidemiology, N.N.Blokhin National Medical Research Centre of Oncology, Moscow, Russia; Department of Urology, N.N.Blokhin National Medical Research Centre of Oncology, Moscow, Russia; Department of Cancer Epidemiology and Genetics, Masaryk Memorial Cancer Institute, Brno, Czech Republic; Institute of Public Health & Preventive Medicine, 2nd Faculty of Medicine, Charles University, Prague, Czech Republic; Department of Oncology, 2nd Faculty of Medicine, Charles University and Motol University Hospital, Prague, Czech Republic; Institute of Hygiene & Epidemiology, 1nd Faculty of Medicine, Charles University, Prague, Czech Republic; Faculty of Health Sciences, Palacky University, Olomouc, Czech Republic; Division of Cancer Epidemiology and Genetics, National Cancer Institute, Rockville, USA; Bevital AS, Bergen, Norway; Observational & Pragmatic Research Institute Pte Ltd, Singapore, Singapore; Department of Pathology, Radboud University Medical Centre, Nijmegen, Netherlands

**Author notes:** Corresponding author: Paul Brennan. These authors contributed equally: Sergey Senkin, Sarah Moody.

## Abstract

International differences in the incidence of many cancer types indicate the existence of carcinogen exposures that have not been identified by conventional epidemiology yet potentially make a substantial contribution to cancer burden^1^. This pertains to clear cell renal cell carcinoma (ccRCC), for which obesity, hypertension, and tobacco smoking are risk factors but do not explain its geographical variation in incidence^2^. Some carcinogens generate somatic mutations and a complementary strategy for detecting past exposures is to sequence the genomes of cancers from populations with different incidence rates and infer underlying causes from differences in patterns of somatic mutations. Here, we sequenced 962 ccRCC from 11 countries of varying incidence. Somatic mutation profiles differed between countries. In Romania, Serbia and Thailand, mutational signatures likely caused by extracts of Aristolochia plants were present in most cases and rare elsewhere. In Japan, a mutational signature of unknown cause was found in >70% cases and <2% elsewhere. A further mutational signature of unknown cause was ubiquitous but exhibited higher mutation loads in countries with higher kidney cancer incidence rates (p-value <6 × 10^−18^). Known signatures of tobacco smoking correlated with tobacco consumption, but no signature was associated with obesity or hypertension suggesting non-mutagenic mechanisms of action underlying these risk factors. The results indicate the existence of multiple, geographically variable, mutagenic exposures potentially affecting 10s of millions of people and illustrate the opportunities for new insights into cancer causation through large-scale global cancer genomics.

## INTRODUCTION

The incidence rates of most adult cancers vary substantially between geographical regions and many such differences are unexplained by known risk factors^1^. Together with unexplained trends in incidence over time, this indicates the likely presence of unknown environmental or lifestyle causes for many cancer types^1^. Traditional epidemiological studies have identified many important lifestyle, environmental and infectious risk factors for cancer. However, they have had limited success in recent decades suggesting that alternative study designs are required if further risk factors are to be identified.

Characterization of mutational signatures within cancer genomes^3^ is an approach complementary to conventional epidemiology for investigating unknown causes of cancer. Most cancers contain thousands of somatic mutations that have occurred over the lifetime of the individual. These can be caused by endogenous cellular processes, such as imperfect DNA replication and repair, or by exposure to exogenous environmental or lifestyle mutagens such as ultraviolet radiation in sunlight and compounds in cigarette smoke. Mutational signatures are the patterns of somatic mutation imprinted on genomes by individual mutational processes. Analysis of thousands of cancer genome sequences from most cancer types has established a set of reference mutational signatures including 71 single base substitution (SBS) or doublet base substitution (DBS) signatures, and 18 small insertion and deletion (ID) signatures^4^. A possible etiology has been suggested for 47 SBS/DBS signatures and nine ID signatures.

Kidney cancer has particularly high incidence rates in Central and Northern Europe, notably in the Czech Republic and Lithuania, and has shown increasing incidence in high income countries in recent decades (**Fig. 1**)^2^. Most kidney cancers are clear cell renal cell carcinomas (ccRCC)^3^ for which obesity, hypertension and tobacco smoking are known risk factors^2^. However, these account for <50% of the global ccRCC burden and do not explain geographical or temporal incidence trends. Previous ccRCC genome sequencing studies have included relatively modest numbers of individuals from a small number of countries with limited variation in ccRCC incidence^5–9^ and have not comprehensively examined associations between ccRCC risk factors and mutational signatures. To detect the activity of unknown carcinogens involved in ccRCC development and to investigate the mechanisms of action of known risk factors, we generated and analyzed epidemiological and whole genome sequencing data from a large international series of ccRCC^10^.

**Fig. 1:**
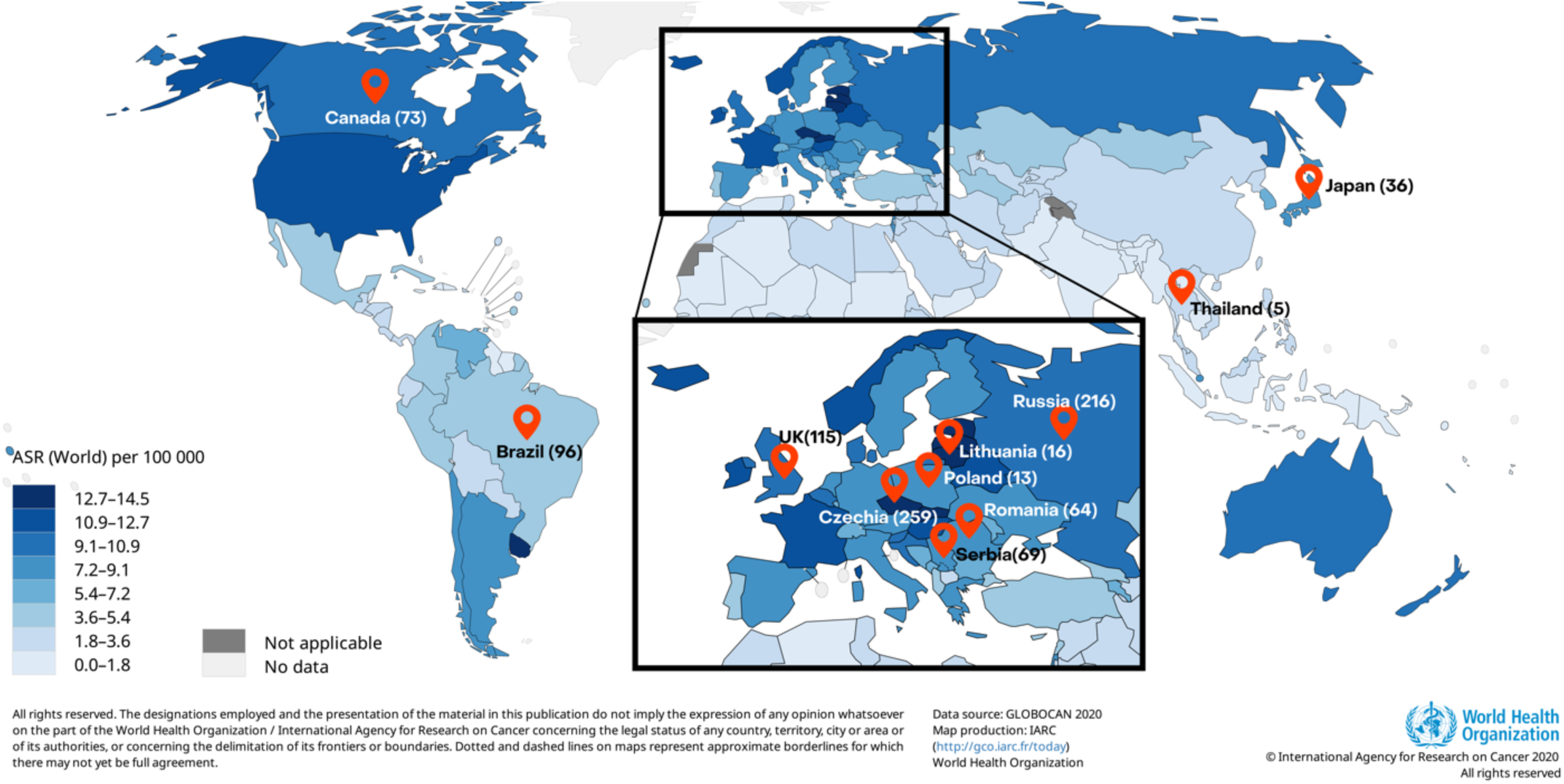
Eleven participating countries and estimated age-standardized incidence rates of clear cell renal cell carcinomas. Incidence of clear cell renal cell carcinomas (ccRCC), men and women combined, age-standardized incidence rates (ASR) per 100,000, data from GLOBOCAN 2020. Markers indicate countries included in this study (number of participating ccRCC patients per country).

## RESULTS

A total of 962 ccRCC cases from 11 countries in four continents were studied, including Czech Republic (*n*=259), Russia (*n*=216), United Kingdom (*n*=115), Brazil (*n*=96), Canada (*n*=73), Serbia (*n*=69), Romania (*n*=64), Japan (*n*=36), Lithuania (*n*=16), Poland (*n*=13), and Thailand (*n*=5; **Fig. 1**; **Table 1**; **Methods**). These encompass a broad range of ccRCC incidence, from the highest global age-standardized rates (ASRs) of Lithuania and Czech Republic (ASRs of 14.5 and 14.4/100,000 respectively) to the relatively low rates of Brazil and Thailand (ASRs of 4.5 and 1.8/100,000 respectively)^11^. Epidemiological questionnaire data were available on sex, age at diagnosis, and important risk factors including body mass index (BMI), hypertension, and tobacco smoking (**Table 1**, **Supplementary Table 1**). DNAs from ccRCCs and blood from the same individuals were extracted and whole genome sequenced to average coverage of 54-fold and 31-fold, respectively.

**Table 1.** Summary of clear cell renal cell carcinomas risk factors included in this study.

| Country<br>(ASR/100,000) |  | Brazil<br>(4.5) | Canada<br>(10.4) | Czechia<br>(14.4) | Japan<br>(7.6) | Lithuania<br>(14.5) | Poland<br>(8.1) | Romania<br>(7.7) | Russia<br>(10.3) | Serbia<br>(7.4) | Thailand<br>(1.8) | UK<br>(10.3) | Total<br>(4.6) |
| --- | --- | --- | --- | --- | --- | --- | --- | --- | --- | --- | --- | --- | --- |
| Number of cases |  | 96 | 73 | 259 | 36 | 16 | 13 | 64 | 216 | 69 | 5 | 115 | 962 |
| Sex | Female | 44 | 22 | 93 | 8 | 9 | 5 | 25 | 98 | 30 | 4 | 42 | 380 |
|  | Male | 52 | 51 | 166 | 28 | 7 | 8 | 39 | 118 | 39 | 1 | 73 | 582 |
| Age at diagnosis<br>(years) | 0-45 | 15 | 6 | 27 | 3 | 1 | 2 | 6 | 43 | 16 | 0 | 6 | 125 |
|  | 45-55 | 20 | 17 | 51 | 5 | 0 | 6 | 10 | 44 | 11 | 0 | 22 | 186 |
|  | 55-65 | 30 | 17 | 77 | 8 | 9 | 1 | 20 | 91 | 27 | 2 | 41 | 323 |
|  | 65-75 | 24 | 27 | 72 | 13 | 4 | 4 | 20 | 32 | 9 | 2 | 31 | 238 |
|  | 75+ | 7 | 6 | 32 | 7 | 2 | 0 | 8 | 6 | 6 | 1 | 15 | 90 |
| Year of<br>recruitment | 1999-2005 |  |  | 93 |  |  | 13 | 14 | 18 |  |  |  | 138 |
|  | 2005-2010 |  |  | 111 |  |  |  | 19 | 70 | 1 |  | 31 | 232 |
|  | 2010-2015 |  | 9 | 55 | 28 |  |  | 31 | 116 | 68 |  | 41 | 348 |
|  | 2015-2020 | 96 | 64 |  | 8 | 16 |  |  | 12 |  | 5 | 43 | 244 |
| Stage | I | 28 | 3 | 123 | 24 | 6 | 0 | 33 | 94 | 32 |  | 53 | 396 |
|  | II | 2 | 0 | 42 | 1 | 0 | 6 | 12 | 24 | 4 |  | 8 | 99 |
|  | III | 16 | 23 | 46 | 6 | 5 | 5 | 18 | 65 | 26 |  | 38 | 248 |
|  | IV | 7 | 10 | 38 | 5 | 2 | 2 | 1 | 33 | 7 |  | 16 | 121 |
|  | Missing | 43 | 37 | 10 |  | 3 |  |  |  |  | 5 |  | 98 |
| Body mass index | <20 | 3 | 2 | 5 | 2 | 0 | 2 | 2 | 9 | 8 | 0 | 6 | 39 |
|  | 20-25 | 21 | 10 | 100 | 25 | 2 | 3 | 17 | 84 | 28 | 3 | 23 | 316 |
|  | 25-30 | 35 | 24 | 85 | 7 | 6 | 6 | 30 | 40 | 20 | 1 | 45 | 299 |
|  | >30 | 37 | 37 | 69 | 2 | 8 | 2 | 14 | 83 | 13 | 1 | 41 | 307 |
|  | Missing |  |  |  |  |  |  | 1 |  |  |  |  | 1 |
| Hypertension | No | 45 | 28 | 129 | 16 | 5 | 9 | 39 | 125 | 28 | 2 | 58 | 484 |
|  | Yes | 51 | 44 | 130 | 20 | 10 | 4 | 24 | 91 | 41 | 3 | 56 | 474 |
|  | Missing |  | 1 |  |  | 1 |  | 1 |  |  |  | 1 | 4 |
| Diabetes | No | 76 | 55 | 130 | 29 | 9 |  | 45 | 186 | 61 | 3 | 95 | 689 |
|  | Yes | 20 | 16 | 36 | 7 | 7 |  | 4 | 12 | 8 | 2 | 20 | 132 |
|  | Missing |  | 2 | 93 |  |  | 13 | 15 | 18 |  |  |  | 141 |
| Family history of<br>ccRCC | No | 90 | 42 | 165 | 35 | 16 |  | 54 | 192 | 67 | 5 | 102 | 726 |
|  | Yes | 5 | 4 | 22 | 1 | 0 |  | 1 | 6 | 2 | 0 | 3 | 43 |
|  | Missing | 1 | 27 | 72 |  |  | 13 | 9 | 18 |  |  | 10 | 193 |
| Tobacco status | Current smoker | 23 | 21 | 66 | 9 | 4 | 6 | 11 | 52 | 18 | 1 | 28 | 239 |
|  | Ex-smoker | 21 | 30 | 62 | 15 | 3 | 3 | 15 | 27 | 15 | 0 | 44 | 235 |
|  | Never | 52 | 22 | 131 | 11 | 9 | 4 | 37 | 137 | 36 | 4 | 43 | 486 |
|  | Missing |  |  |  | 1 |  |  | 1 |  |  |  |  | 2 |
| PFOA (ng/mL) | Mean<br>(st. dev.) | 0.7<br>(0.5) | 1.6<br>(1.1) | 3.4<br>(2.1) |  | 1.3<br>(0.6) | 5.4<br>(4.1) | 1.3<br>(0.9) | 1.5<br>(1.4) | 1.3<br>(0.6) | 2.2<br>(2.2) | 3.3<br>(1.7) | 2.2<br>(1.9) |

Somatic mutation burdens in the 962 ccRCC genomes ranged from 803 to 45,376 (median 5,093) for single base substitutions (SBS), 2 to 240 (median 53) for doublet base substitutions (DBS), and 10 to 14,770 (median 695) for small insertions and deletions (**Supplementary Table 2**). The average burden of all three mutation types differed between the 11 countries (p-value <2 × 10^−23^, p-value <2 × 10^−14^, p-value <6 × 10^−14^, for SBSs, DBSs, and IDs, respectively). In particular, the burden of all mutation types was elevated in Romania compared to other countries (**Extended Data Fig. 1**). Principal Component Analysis (PCA) performed on the proportions of the six primary SBS mutation classes (C>A, C>G, C>T, T>A, T>C, T>G) in each sample identified a distinct cluster of mainly Romanian and Serbian cases and a further cluster of mainly Japanese cases (**Extended Data Fig. 2**). The results, therefore, clearly demonstrate geographical variation of somatic mutation loads and patterns in ccRCC.

To investigate the mutational processes contributing to the geographical variation in mutation burdens we extracted mutational signatures and estimated the contribution of each signature to each ccRCC genome (**Supplementary Tables 3-7**). Ten signatures with strong similarity to a reference signature in the Catalogue of Somatic Mutations in Cancer (COSMIC) database were extracted: SBS1, due to deamination of 5-methylcytosine^12^; SBS2 and SBS13, due to cytosine deamination by Apolipoprotein B mRNA-editing enzyme, catalytic polypeptide-like (APOBEC) DNA editing enzymes^12^; SBS4, due to tobacco smoke mutagens^13^; SBS5, due to an endogenous mutational process in which mutations accumulate with age^13^; SBS12, of unknown cause; SBS18, due to DNA damage by reactive oxygen species^13^; SBS21 and SBS44, due to defective DNA mismatch repair^13,14^; and SBS22, due to Aristolochic acid exposure^15,16^.

Five further SBS signatures were identified which were not well described by the COSMIC catalogue (**Fig. 2**; **Supplementary Table 8**). SBS40a, SBS40b and SBS40c were present in most ccRCC, accounting for, on average, ∼30%, ∼20%, and ∼3% of mutations respectively (**Fig. 2b**). Combined, they closely resemble the previously reported SBS40 (0.96 cosine similarity), suggesting that the large number of ccRCC whole genomes analyzed here provides the power to separate the constituent component signatures of SBS40. SBS40 was previously reported frequently, and at high levels, in kidney cancer, but also in other cancers, and is of unknown etiology. Like the composite SBS40, SBS40a is present in multiple cancer types. However, SBS40b and SBS40c are largely restricted to ccRCC (**Supplementary Note, Extended Data Fig. 3**). SBS_H was found in a single case and SBS_I is related to Aristolochic acid exposure (see below; SBS_I has been renamed as SBS22b). Analysis of all other types of mutational signatures, including doublet bases substitutions, small insertion and deletions, copy number variants and structural variants, is presented in Supplementary results.

**Fig. 2:**
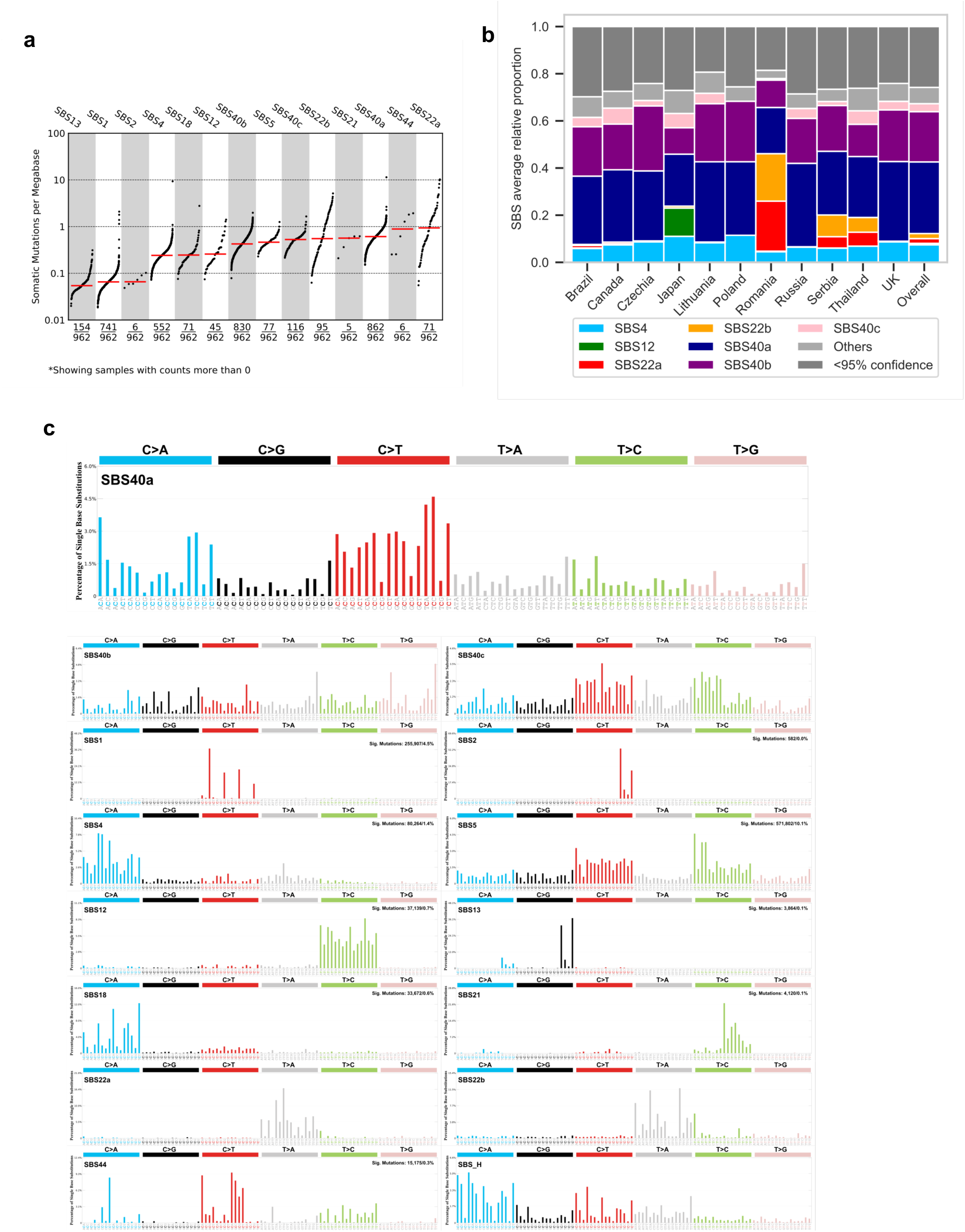
Single base substitution signature operative in clear cell renal cell carcinomas. **(a)** TMB plot showing the frequency and mutations per Mb for each of the decomposed SBS signatures. **(b)** Average relative attribution for single base substitution (SBS) signatures across countries. Signatures contributing less than 5% on average are grouped in the ‘Others’ category, apart from SBS12 and AA-related signatures SBS22a and SBS22b. ‘<95% confidence’ category accounts for the proportion of mutation burden which could not be assigned to any signature with confidence level of at least 95%. **(c)** Decomposed signatures, including reference COSMIC signatures as well as *de novo* signatures not decomposed into COSMIC reference signatures.

The mutation burdens of multiple SBS mutational signatures varied between the 11 countries. SBS22 is thought to be caused by Aristolochic acids, mutagenic derivatives of plants of the Aristolochia genus which are carcinogenic and also cause Balkan endemic nephropathy (BEN), a kidney disease prevalent in areas adjacent to the Danube in Southeastern Europe^17^. SBS22 has previously been found in ccRCC, other urothelial tract cancers, and hepatocellular carcinomas from Romania^5,18^ and various countries in East and South-East Asia^15,16,19^. In this study, SBS22 was present in high proportions of ccRCC from Romania (45/64, 70%), Serbia (16/69, 23%), and Thailand (3/5, 60%), often with very high mutation burdens. Of note, given the limited number of cases in Thailand, they may not be representative of ccRCC in that region. The presence of SBS22 was strongly correlated with that of new signatures SBS_I, DBS_D, and ID_C (**Extended Data Fig. 4-6**) which are, therefore, also probably due to Aristolochic acid exposure. SBS_I, like SBS22, is composed predominantly of T>A mutations. The signature identified previously as SBS22, has therefore been renamed SBS22a, and SBS_I has been named SBS22b. The mutation burden of both SBS22a and SBS22b differed between Serbia and Romania, with higher levels being detected in Romania, and away from recognized BEN zones (**Fig 3a-c**). The two signatures may be due to different subsets of Aristolochic acids, and/or to different metabolites, which induce slightly different mutational patterns. Only five ccRCC cases were known to reside within recognized BEN zones, suggesting no clear link between the two diseases. While the source of this exposure is uncertain, these results indicate that a substantial proportion of the population over a wide geographical area of Eastern Europe, possibly numbering in the 10s of millions, has been exposed to Aristolochic acid containing compounds, the public health consequences of which are uncertain.

**Fig. 3:**
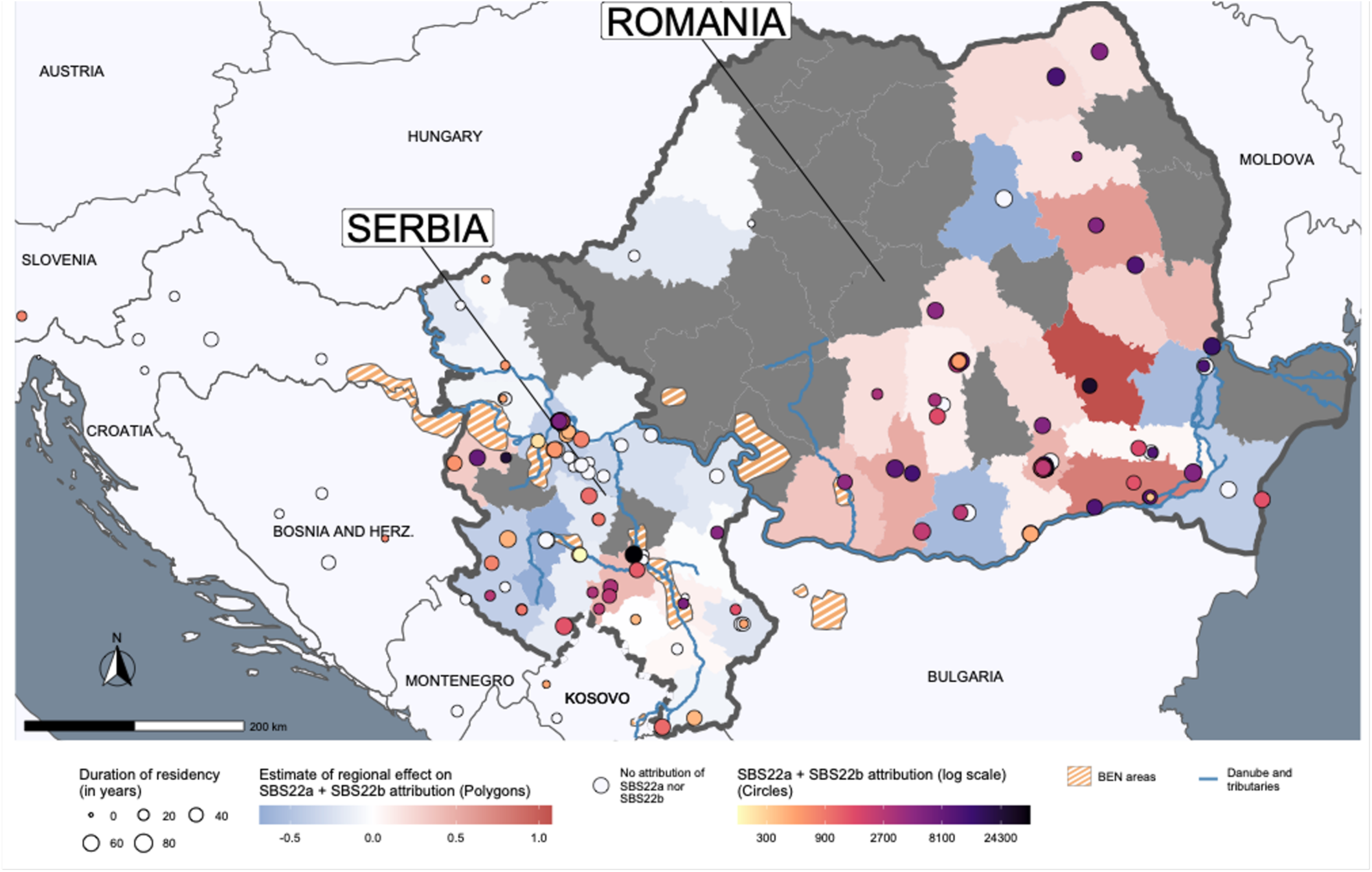
Geospatial analysis of Aristolochic acid-related SBS signatures. Distribution of Romanian and Serbian cases with known residential history, along with the summed levels of SBS22a and SBS22b attributions (per-case and regional estimate), with respect to the Balkan endemic nephropathy (BEN) areas. White circles represented cases with no detected activity of SBS22a and SBS22b.

SBS12 was present in 72% of Japanese and 2% of non-Japanese ccRCC (p-value=4.7 × 10^−78^) (**Extended Data Fig. 7h**). Compared to the mutation burdens imposed by Aristolochic acid in ccRCC, SBS12 contributed modest mutation loads. SBS12 is composed predominantly of T>C substitutions and exhibits strong transcriptional strand bias with more T>C mutations on the transcribed than untranscribed strands of protein coding genes. Transcriptional strand bias is typically caused by activity of transcription-coupled nucleotide excision repair acting on bulky DNA adducts due to exogenous mutagenic exposures such as tobacco smoke chemicals^13^, ultraviolet light^13^, Aristolochic acids^15^, and aflatoxins^20^. Assuming that transcription-coupled repair of DNA adducts is responsible for the SBS12 strand bias, the adducts are likely on adenine. Alternatively, transcriptional strand bias can also be caused by transcription-coupled damage^21,22^. The presence of SBS12 was replicated in two further series of whole genome sequenced ccRCC from Japan including 14 cases from an independent study group who undertook a broad genomic analysis of ccRCC but without detailed mutational signature analysis^23^, and a more recent unpublished series of 61 cases from an additional cohort of ccRCC sequenced by the same center as the initial cohort (**Supplementary Note**). SBS12 was present in 12/14 cases (85%) cases and 46/61 (75%) of cases, respectively. SBS12 was previously reported in hepatocellular carcinomas^4,13^ and additional analysis of existing datasets revealed strong SBS12 enrichment in hepatocellular carcinomas from Japan compared to other countries (p-value=3.8 × 10^−15^; **Supplementary Note**). These results, therefore, indicate that exposure to an agent contributing SBS12 mutations to kidney and liver cancer is common in Japan and rare in the other 10 countries included in this study. The agent responsible for SBS12 is unknown although the precedents provided by other mutational signatures with strong transcriptional strand bias suggest that it is likely of exogenous origin^21,22^. A polymorphism in aldehyde dehydrogenase 2 known to impair metabolism of alcohol to aldehydes and common in Japan did not associate with levels of SBS12, and neither did any other common germline variants (**Supplementary Note**).

SBS40a, SBS40b, and SBS40c were present in ccRCC from all 11 countries. The country-specific average mutation burdens of SBS40a and SBS40b positively associated with country-specific ASRs of kidney cancer incidence (p-value=0.0022 and p-value=5.1 × 10^−18^, respectively; **Extended Data Fig. 8a**; **Fig. 4a**, **Supplementary Table 9**), with the highest mutation loads in the Czech Republic and Lithuania. Kidney cancer incidence rates also vary between the regions of the Czech Republic and SBS40b mutation burdens differed significantly between these (p-value=0.011; **Fig 4b,c**, **Supplementary Table 10**), with the highest attribution in the highest risk region. SBS40b exhibits modest transcriptional strand bias and, assuming that transcription-coupled repair of DNA adducts is responsible, the adducts underlying SBS40b are likely on pyrimidines. Insertion and deletion (indel) signatures ID5 and ID8, which together contributed ∼60% of the indel mutation burden on average, were also strongly associated with country-specific kidney cancer ASR (p-value=1.3 × 10^−10^ and p-value=7.1 × 10^−5^, respectively, **Extended Data Fig. 8b,c**). Signatures ID5 and ID8 correlated with each other (Spearman’s r=0.78), as well as with SBS40b (r=0.79 and r=0.74, respectively) indicating that they likely all constitute products of the same underlying mutational process. Thus, the burdens of the full complement of mutation types generated by this mutational process correlate with age-adjusted kidney cancer incidence rates. The overall mutational burden did not, however, associate significantly with kidney cancer incidence rates (**Extended Data Fig. 9**).

**Fig. 4:**
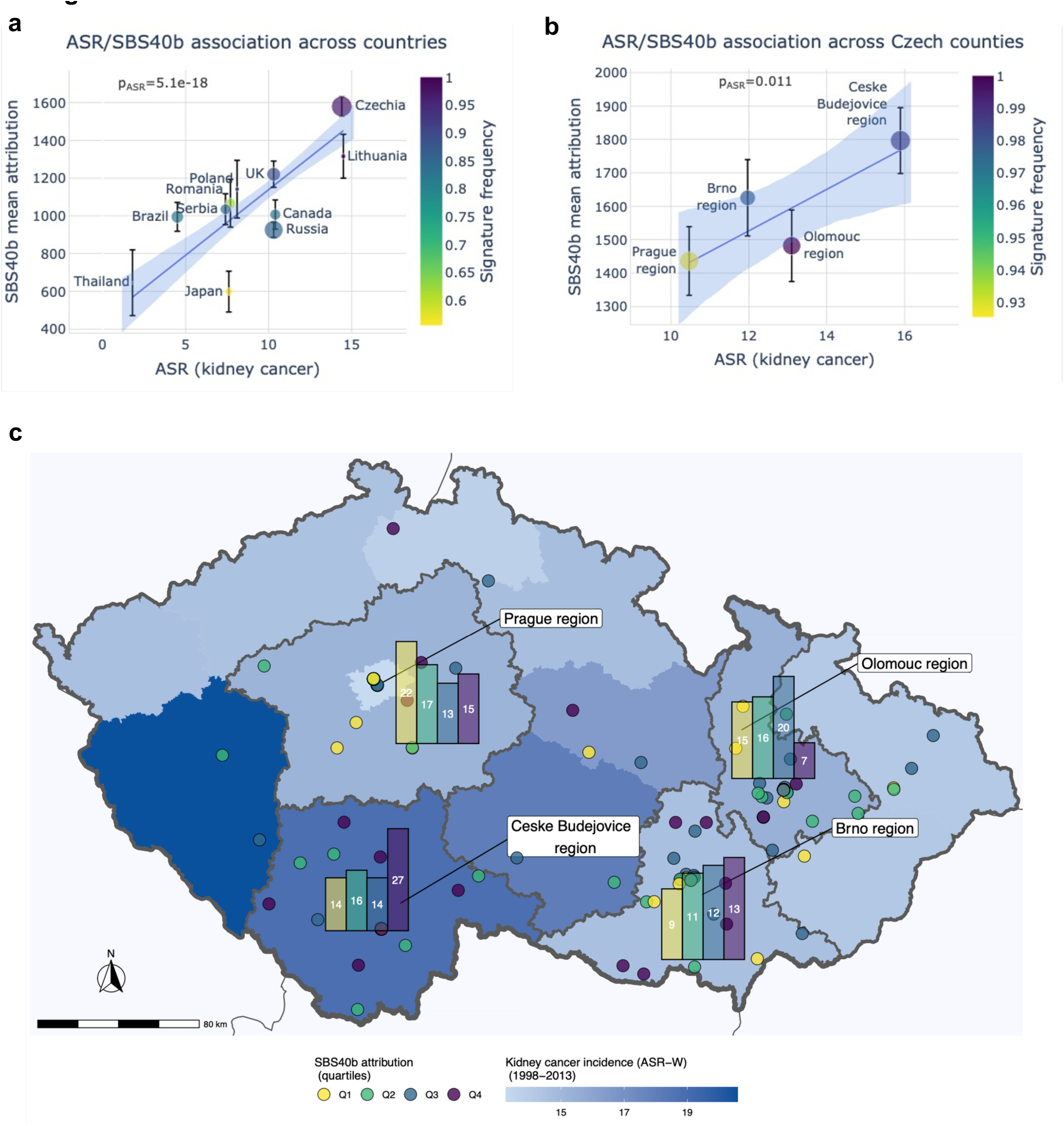
Association of SBS40b signature attribution with incidence of kidney cancer. **(a)** Number of mutations attributed to signature SBS40b against age-standardized incidence rates (ASR) of kidney cancer in each of the eleven countries represented in the cohort. Error bars represent standard errors of the mean. **(b)** Number of mutations attributed to signature SBS40b in four regions of Czech Republic against ASR of kidney cancer in each region. Error bars represent standard errors of the mean. In **(a)** and **(b)**, the p-values are shown for the ASR variable in linear regressions across all cases, adjusted for sex and age of diagnosis. **(c)** Levels of attribution of SBS40b signature within Czech Republic, with bar plots showing the number of cases for each quartile of SBS40b attribution across Prague, Olomouc, Ceske Budejovice, and Brno regions.

To investigate potential mutagenic agents underlying these geographically variable signatures, an untargeted metabolomics screen of plasma was conducted on 901 individuals in the study, from all countries except Japan (**Methods**). 2,392 metabolite features were obtained, including 944 independent peaks (r<0.85). Three features were associated with SBS4 (**Supplementary Table 11**), with two identified as hydroxycotinine (p-value=2.9 × 10^-9^) and cotinine (p-value=1.9 × 10^-5^), two major metabolites of nicotine^24^. Eight features were associated with SBS40b (**Supplementary Table 11**). One feature was identified as N,N,N-trimethyl-L-alanyl-L-proline betaine (TMAP; p-value=1.2 × 10^-5^, **Supplementary Table 12**), increased levels of which correlate strongly with reduced kidney function^25^. Other established measures of kidney function, including cystatin C and creatinine, were correlated with TMAP (p-value = 2.5 x 10^-30^ and 1.7 × 10^-69^, respectively) and also showed evidence of positive association with SBS40b (p-value=0.023 and 0.058, respectively). Thus, exposure to the mutagenic agent responsible for SBS40b is associated with reduced kidney function. No recognized metabolome features were significantly associated with any other signatures.

A total of 1962 “driver” mutations were found in 136 genes including *VHL*, *PBRM1*, *SETD2* and *BAP1*, the known frequently mutated cancer genes in ccRCC (Methods) (**Fig. 5a**, **Supplementary Table 13**)^9,26^. The frequencies of mutations in these genes were consistent across countries (**Fig. 5b**). The spectrum of all driver mutations in ccRCC with Aristolochic acid exposure (**Methods**) was enriched in T>A mutations compared to non-exposed cases (25% vs 13%, p-value=0.0062, **Fig. 5c,d**) with similar enrichment specifically in VHL mutations (30% vs 16%; **Fig. 5e,f**), and in the whole exomes (27% in exposed compared to 12% in unexposed cases). Thus genome-wide Aristolochic acid mutagenesis has contributed in a proportionate fashion to generation of driver mutations in Aristolochic acid-exposed ccRCC. The driver mutation spectrum did not show statistically significant enrichment of T>C mutations in SBS12 exposed cases (20% vs 12%, p = 0.069), but was consistent with the level of enrichment in the exome (21% in exposed compared to 15% in unexposed cases). SBS40b also did not show statistically significant enrichment possibly due to the ubiquitous exposure and its relatively flat and featureless mutation profile.

**Fig. 5:**
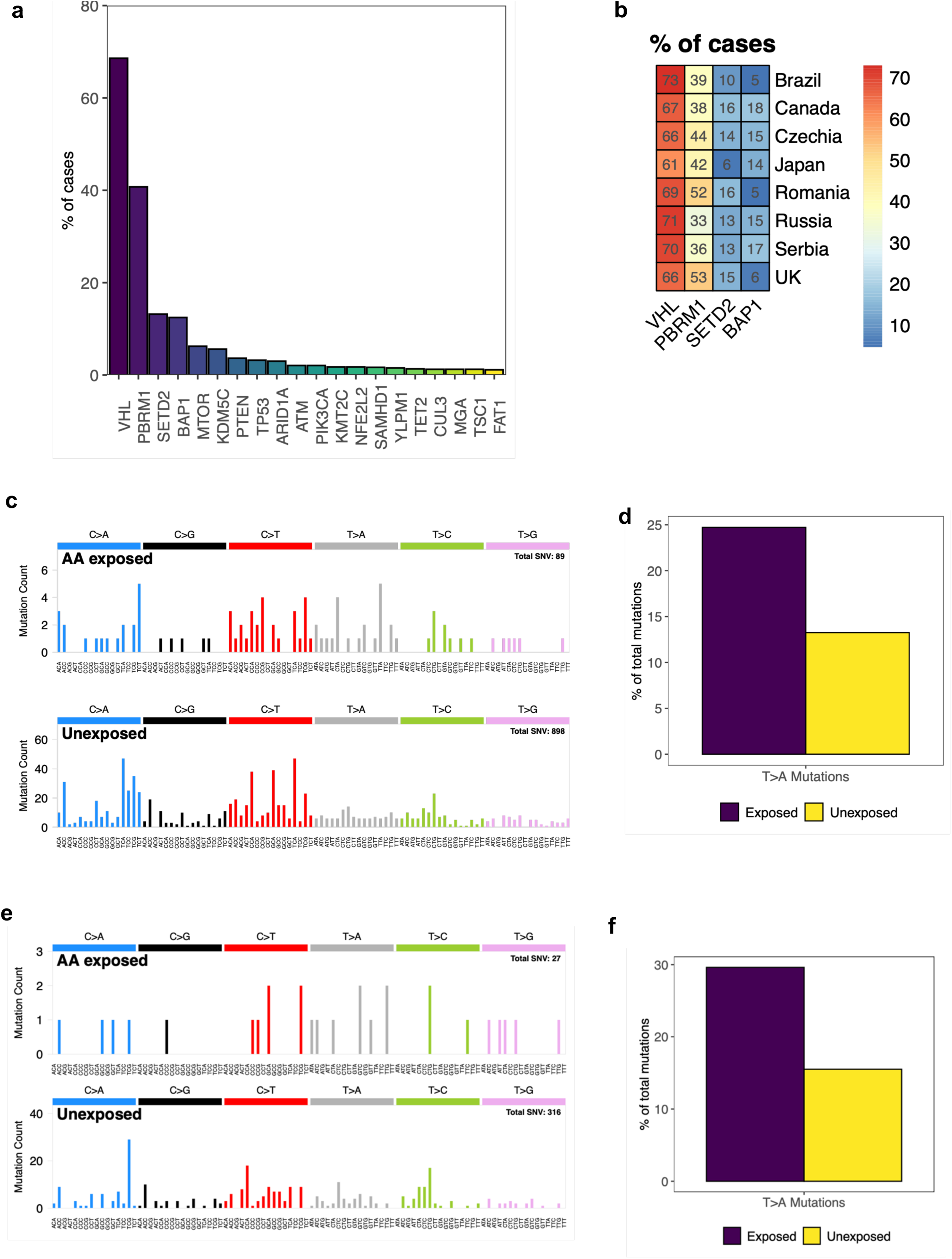
Driver mutation analysis in clear cell renal cell carcinomas. **(a)** Frequency of driver genes in the cohort. Only genes mutated in at least 10 cases are shown. **(b)** Frequency of driver genes across countries. Thailand, Poland and Lithuania are not shown due to low sample numbers. **(c)** SBS-96 mutational spectra of all driver mutations in ccRCC for Aristolochic acid (AA)-exposed and unexposed cases. **(d)** Percentage of T>A driver mutations in AA-exposed and unexposed cases. **(e)** SBS-96 mutational spectra of VHL mutations in ccRCC for AA-exposed and unexposed cases. **(f)** Percentage of T>A VHL mutations in AA-exposed and unexposed cases.

Exogenous mutagenic exposures that ultimately cause cancer may be present during the early stages of evolution of cancer clones. To time mutagenic exposures, the contribution of each mutational signature to mutations in the primary clone (relatively early) and to mutations in subclones (relatively late) were estimated^27,28^ (**Methods**). All signatures of the putative exogenous mutagenic exposures observed in ccRCC were present at relatively early stages of cancer development, consistent with exposures to normal cells. SBS12, SBS22b, and SBS40b showed higher activities in main clones compared to subclones (q-value=0.04, q-value=0.02, q-value=2.3 × 10^−5^, respectively) (**Extended Data Fig. 10**) and SBS22a showed no significant difference^15,16^. By contrast, signatures due to endogenous mutational processes including APOBEC DNA editing (SBS13) and oxidative damage (SBS18), were enriched in subclones (q-value=1.6 × 10^−4^, q-value=3.2 × 10^−7^, respectively).

Established or suspected risk factors for ccRCC include age, tobacco smoking, obesity, hypertension, diabetes, and environmental exposure to PFAS compounds^29^. Total SBS, DBS, and ID mutation burdens associated with age, as did SBS1, SBS4, SBS5, SBS40a, SBS40b, SBS22a, SBS22b, DBS2, ID1, ID5, and ID8. Total SBS (p-value=3.1 × 10^−5^), DBS (p-value=3.7 × 10^−3^) and ID (p-value=1.3 × 10^-4^) mutation burdens also associated with sex, with males having higher mutation burdens than females, and with SBS40b showing a similar association (p-value=9.3 × 10^−5^). Associations with tobacco smoking were observed for SBS4 (p-value=5.3 × 10^−6^) and DBS2 (p-value=2.4 × 10^−7^), both known to be caused by tobacco carcinogens^30,31^. These results suggest that the known increased risk of ccRCC with tobacco smoking is due to direct exposure of the kidney to tobacco related mutagens (**Supplementary Note**). Associations of particular mutational signatures with other ccRCC risk factors were not observed (**Supplementary Tables 14, 15**). To complement this analysis of observational data, associations between polygenic risk scores for known ccRCC risk factors and mutational signatures^32,33^ were examined (**Methods**). Consistent with the observational data, no associations were found between genetically inferred risk factors and mutational signatures except for tobacco smoking and DBS2 (p-value=0.01; **Supplementary Table 16**).

## DISCUSSION

Somatic mutations in the genomes of 962 ccRCC patients from 11 countries indicate the existence of multiple, widespread mutational processes exhibiting substantial geographical variation in their contributions to ccRCC mutation loads. The results contrast with those from 552 esophageal squamous carcinomas from eight countries with widely different esophageal carcinoma incidence rates in which geographical differences in mutation burdens or signatures were not observed^34^. Together the studies implicate both geographically variable mutagenic and non-mutagenic carcinogenic exposures contributing to global cancer incidence. Indeed, the presence of mutational signatures associated with tobacco smoking but absence of signatures associated with other known ccRCC risk factors, such as obesity and hypertension, suggests that the latter may be mediated by non-mutagenic processes and, therefore, that both classes of carcinogen contribute to the development of ccRCC.

The existence, identity, and carcinogenic effect of some of the agents underlying these mutational processes are known. Aristolochic acids are believed to cause SBS22a/b, and its associated signatures, but this study suggests that the geographical extent and proportion of the population acquiring mutations in South-Eastern Europe is far greater than previously anticipated, possibly affecting 10s of millions of individuals. The sources of the Aristolochic acid exposure, the manner by which it is ingested and whether the exposure continues today are uncertain, and further definition of the source and extent of this exposure is required in order to provide a foundation for public health action.

The existence of the mutagenic exposures underlying SBS12 and SBS40b were not previously suspected, and their causative agents are unknown. Based on current information, the exposure causing SBS12 is restricted to Japan. However, larger studies are now indicated to explore the geographical extent of exposure in Japan and neighboring countries, and the proportions of their populations that have been exposed. Studies of Japanese migrants to other countries are also likely to be informative regarding the potential source of exposure. In the first instance this will be achievable by further sequencing of kidney and hepatocellular cancer genomes. However, studies of normal tissues, using recently reported sequencing methods allowing detection of somatic mutations in normal cells^35^, and particularly relatively accessible ones such as cells in urine that can be prospectively collected, may enable large population-based studies providing better characterization of the exposure and its consequences. As with exposure to Aristolochic acid in South-Eastern Europe, it is possible that 10s of millions of individuals in East Asia are exposed to a potent mutagen, and identification of the source and extent of exposure would seem to be a public health priority.

In contrast to Aristolochic acid and the agent causing SBS12, the exposure underlying SBS40b appears to be globally ubiquitous. It predominantly causes mutations in ccRCC, with much lower burdens in other cancer types, and generates mutation loads correlating strongly with age and sex. There are few clues as to its origin or nature.

The incidence rates of ccRCC vary ∼eightfold across the eleven countries from which ccRCCs were sequenced. A strong positive correlation (p-value=5.5 × 10^−18^) was found between the average mutation loads attributable to SBS40b in each country (and also those of ID5 and ID8 which are correlated with SBS40b) and incidence of kidney cancer within each country. This correlation reflects approximately a tripling of average country-specific SBS40b mutation loads (a difference of ∼1000 mutations) in parallel with the eightfold increase of country-specific ASR.

SBS40b mutation burdens also positively correlated with biomarkers of impaired kidney function, reminiscent of the nephrotoxic effects of Aristolochic acids in Balkan endemic nephropathy. It is possible that the increased SBS40b somatic mutation load itself engenders this reduction in renal function. However, studies of other normal tissues suggest that they are generally tolerant of elevated mutation burdens, except for manifesting a higher incidence of neoplasia^36,37^. It is also possible that the agent underlying SBS40b is directly nephrotoxic, for example by engendering DNA damage and a response to it, and that the mutations it generates are immaterial to kidney function. It is also conceivable, however, that impaired renal function, potentially due to many different causes, results in a metabolic state which itself causes the elevated SBS40b mutation load. Whatever the mutational process underlying SBS40b, it is plausible that it contributes to the geographical variation in the age standardized rates of kidney cancer incidence rates. It is of public health interest to determine the cause of SBS40b and, hence, to consider whether the exposure can be mitigated, potentially with concomitant reduction in global ccRCC incidence rates.

The absence of any association between several known risk factors for ccRCC and mutation burden, in particular for obesity and hypertension, supports a model of cancer development where mutations are essential but additional factors affect the expansion of a mutated clone and thus the chance of it progressing into cancer^38^. Further efforts at defining how lifestyle and environmental exposures contribute to cancer development will therefore require a greater understanding of both the causes of the mutations in cell clones in normal tissue, and the further promotion of such mutant clones by non-mutagenic processes.

Finally, the substantial geographical variability of SBS12 and SBS22a/b, with most countries not showing evidence of exposure, raises the possibility that additional mutational signature studies of ccRCC involving more countries may reveal further mutagenic exposures. Furthermore, the results relating to SBS40b highlight the prospect that a significant proportion of global cancer burden may be caused by relatively ubiquitous exposures that are not readily detectable by classical cancer epidemiology studies. The conduct of large scale whole genome sequencing for other cancer sites across high and low risk populations around the world would seem to be an appropriate strategy for detecting such novel cancer causing agents.

## Data Availability

Whole genome sequencing data and patient metadata are deposited in the European Genome-phenome Archive (EGA) associated with study EGAS00001003542. Aligned BAM files for all ccRCC cases included in the final analysis were deposited in dataset EGAD00001012102, variant calling files in dataset EGAD00001012222 and patient metadata in dataset EGAD00001012223. The metabolomics data are available upon request. All other data are provided in the accompanying Supplementary Tables.

https://ega-archive.org/studies/EGAS00001003542

https://ega-archive.org/datasets/EGAD00001012102

https://ega-archive.org/datasets/EGAD00001012222

https://ega-archive.org/datasets/EGAD00001012223

## ONLINE METHODS

### Recruitment of cases and informed consent

The International Agency for Research on Cancer (IARC/WHO) coordinated case recruitment through an international network of over 40 collaborators from the 11 participating countries (**Table1**; **Supplementary Table 17**). The inclusion criteria for patients were >=18 years of age (ranging from 23 to 87, with a mean of 60 and a standard deviation of 12), confirmed diagnosis of primary ccRCC and no prior cancer treatment. Informed consent was obtained for all participants. Patients were excluded if they had any condition that could interfere with their ability to provide informed consent or if there were no means of obtaining adequate tissues or associated data as per the protocol requirements. Ethical approvals were first obtained from each Local Research Ethics Committee and Federal Ethics Committee when applicable, as well as from the IARC Ethics Committee.

### Bio-samples, data collection, and expert pathology review

Dedicated standard operating procedures, following guidelines from the International Cancer Genome Consortium (ICGC), were designed by IARC/WHO to select appropriate case series with complete biological samples and exposure information as described previously^1^ (**Supplementary Table 17**). In brief, for all case series included, anthropometric measures were taken, together with relevant information regarding medical and familial history. Comparable smoking and alcohol history was available from all centers. Detailed epidemiological information on residential history was collected in Czech Republic, Romania, and Serbia. Potential limitations of using retrospective clinical data collected using different protocols from different populations were addressed by a central data harmonization to ensure a comparable group of exposure variables (**Supplementary Table 17**). All patient related data as well as clinical, demographical, lifestyle, pathological and outcome data were pseudonymized locally through the use a dedicated alpha-numerical identifier system before being transferred to IARC/WHO central database.

Original diagnostic pathology departments provided diagnostic histological details of contributing cases through standard abstract forms. IARC/WHO centralized the entire pathology workflow and coordinated a centralized digital pathology examination of the frozen tumor tissues collected for the study as well as formalin-fixed, paraffin-embedded (FFPE) sections when available, via a web-based report approach and dedicated expert panel following standardized procedures as described previously^1^. A minimum of 50% viable tumor cells was required for eligibility to whole genome sequencing.

In summary, frozen tumor tissues were first examined to confirm the morphological type and the percentage of viable tumor cells. A random selection of tumor tissues was independently evaluated by a second pathologist. Enrichment of tumor component was performed by dissection of non-tumoral part, if necessary. 90 cases overlapped with a previously published cohort recruited under the Cancer Genomics of the Kidney (CAGEKID) project^2^, which were also part of the Pan-Cancer Analysis of Whole Genomes (PCAWG) project^3^.

### DNA extraction

Extraction of DNA from fresh frozen tumor and matched blood samples was centrally conducted at IARC/WHO except for Japan, which performed DNA extractions at the local center following a similarly standardized DNA extraction procedure. Of the cases which proceeded to the final analysis (*n*=962), germline DNA was extracted from either buffy-coat, whole blood, or from adjacent normal tissue (*viz.*, samples from Japan) using previously described protocols and methods^1^.

### Whole genome sequencing

In total, 1583 renal cell carcinoma cases were evaluated, with 1267 confirmed as ccRCC cases. 116 (9%) were excluded due to insufficient viable tumor cells (pathology level), or inadequate DNA (tumor or germline). DNA from 1151 cases was received at the Wellcome Sanger Institute for whole genome sequencing. Fluidigm SNP genotyping with a custom panel was performed to ensure that each pair of tumor and matched normal samples originated from the same individual. Whole genome sequencing (150bp paired end) was performed on the Illumina NovaSeq 6000 platform with target coverage of 40X for tumors and 20X for matched normal tissues. All sequencing reads were aligned to the GRCh38 human reference genome using Burrows-Wheeler-MEM (v0.7.16a and v0.7.17). Post-sequencing QC metrics were applied for total coverage, evenness of coverage and contamination. Cases were excluded if coverage was below 30X for tumor or 15X for normal tissue. For evenness of coverage, the median over mean coverage (MoM) score was calculated. Tumors with MoM scores outside the range of values determined by previous studies to be appropriate for whole genome sequencing (0.92 – 1.09) were excluded. Conpair^4^ (https://github.com/nygenome/Conpair) was used to detect contamination, cases were excluded if the result was greater than 3%^5^. A total of 962 cases passed all criteria and were included in subsequent analysis.

### Somatic variant calling

Variant calling was performed using the standard Sanger bioinformatics analysis pipeline (https://github.com/cancerit). Copy number profiles were determined first using the algorithms ASCAT^6^ and BATTENBERG^7^, where tumor purity allowed. SNV were called with cgpCaVEMan^8^, indels were called with cgpPINDEL^9^, and structural rearrangements were called using BRASS. CaVEMan and BRASS were run using the copy number profile and purity values determined from ASCAT where possible (complete pipeline, n=857). Where tumor purity was insufficient to determine an accurate copy number profile (partial pipeline, n=105), CaVEMan and BRASS were run using copy number defaults and an estimate of purity obtained from ASCAT/BATTENBERG. For SNV additional filters (ASRD >= 140 and CLPM ==0) were applied to remove potential false positive calls. A second variant caller, Strelka2, was run for SNVs and indels as consensus variant calling was previously shown to eliminate algorithm specific artefacts and to generate highly reproducible mutational spectra compared to using a single variant calling algorithm^1,10^. Only variants called by both the Sanger variant calling pipeline and Strelka2 were included in subsequent analysis.

### Validation of Japanese sequencing

The matched normal tissue sequenced was blood for all countries with the exception of Japan, where adjacent normal kidney was used. As Japan displayed an enrichment of SBS12, matched blood was obtained from 28 of the 36 patients to confirm that the source of the matched normal tissue was not influencing the result. In all cases, the mutational spectra of Japanese ccRCC generated using either blood or adjacent normal kidney matched each other with a cosine similarity of >0.99.

### Generation of mutational matrices

Mutational matrices for single base substitutions (SBS), doublet base substitutions (DBS) and small insertions and deletions (ID) were generated using SigProfilerMatrixGenerator (https://github.com/AlexandrovLab/SigProfilerMatrixGenerator) with default options (v1.2.12)^11^.

### Mutational signature analysis

Mutational signatures were extracted using two algorithms, SigProfilerExtractor (https://github.com/AlexandrovLab/SigProfilerExtractor), based on nonnegative matrix factorization, and mSigHdp^12^ (https://github.com/steverozen/mSigHdp), based on the Bayesian hierarchical Dirichlet process. For SigProfilerExtractor, *de novo* mutational signatures were extracted from each mutational matrix using SigProfilerExtractor with nndsvd_min initialization (NMF_init=“nndsvd_min”) and default parameters (v1.1.9)^13^. Briefly, SigProfilerExtractor deciphers mutational signatures by first performing Poisson resampling of the original matrix with additional renormalization (based on a generalized mixture model approach) of hypermutators to reduce their effect on the overall factorization^13^. Nonnegative matrix factorization (NMF) was performed using initialization with nonnegative singular value decomposition and by applying the multiplicative update algorithm using the Kullback–Leibler divergence as an objective function^13^. NMF was applied with factorizations between *k*=1 and *k*=20 signatures; each factorization was repeated 500 times^13^. *De novo* single base substitution mutational signatures were extracted with SigProfilerExtractor for both SBS-288 and SBS-1536 contexts^11^. The results were largely concordant with the SBS-1536 *de novo* signatures allowing additional separation of mutational processes, therefore the SBS-1536 *de novo* signatures were taken forward for further analysis (**Supplementary Table 3**). Mutational signatures for DBS and ID were extracted in DBS-78 and ID-83 contexts respectively (**Supplementary Tables 4, 5**). Where possible, SigProfilerExtractor matched each *de novo* extracted mutational signature to a set of previously identified COSMIC signatures^14^, for SBS-1536 signatures this requires collapsing the 1536 classification into the standard 96 substitution type classification with six mutation classes having single 3’ and 5’ sequence contexts (**Supplementary Table 8**). This step makes it possible to distinguish between *de novo* signatures which can be explained by a combination of the known catalog of mutational process (which have not been completely separated during the extraction), and those which have not been previously identified. mSigHdp extraction of SBS-96 and ID-83 signatures was performed using the suggested parameters and using the country of origin to construct the hierarchy. SigProfilerExtractor’s decomposition module was subsequently used to match mSigHdp *de novo* signatures to previously identified COSMIC signatures^14^. Further details on the comparison of results between SigProfilerExtractor and mSigHdp and decomposition of *de novo* signatures into COSMIC reference signatures can be found in the **Supplementary Note.**

### Attribution of activities of mutational signatures

The *de novo* (SigProfiler) and COSMIC signature (SigProfiler and mSigHdp) activities were attributed for each sample using the MSA signature attribution tool (v2.0, https://gitlab.com/s.senkin/MSA)^15^. For COSMIC attributions, only COSMIC reference signatures, which were identified in the decomposition of *de novo* signatures, were included in the panel for attribution, in addition to *de novo* signatures which could not be decomposed into COSMIC reference. At its core, the tool utilizes the nonnegative least squares (NNLS) approach minimizing the L2 distance between the input sample and the one reconstructed using available signatures. To limit false positive attributions, an automated optimization procedure was applied by repeated removal of all signatures that do not increase the L2 similarity of a sample by >0.008 for SBS, >0.014 for DBS, and >0.03 for ID mutation types, as suggested by simulations. These optimal penalties were derived using an optional parameter (params.no_CI_for_penalties = false) utilizing a conservative approach in calculation of penalties. Finally, a parametric bootstrap approach was applied to extract 95% confidence intervals for each attributed mutational signature activity.

### Driver mutations

A dNdS approach was used to identify genes under positive selection in ccRCC^16^. The analysis was performed both for the whole genome (q-value<0.01), and with restricted hypothesis testing (RHT) for a panel of 369 known cancer genes^16^. Variants in any gene identified as under positive selection in global dNdS or in the 369-cancer gene panel were assessed as potential drivers^16^. Candidate driver mutations were annotated with the mode of action using the Cancer Gene Census (https://cancer.sanger.ac.uk/census) and the Cancer Genome Interpreter tool (https://www.cancergenomeinterpreter.org). Missense mutations were assessed using the MutationMapper tool (http://www.cbioportal.org/mutation_mapper). Variants were considered likely drivers if they met any of the following criteria: *(i)* Truncating mutations in genes annotated as tumor suppressors; *(ii)* mutations annotated as likely or known oncogenic in MutationMapper; (iii) truncating variants in genes with selection (q-value<0.05) for truncating mutations assumed to be tumor suppressors and thus likely drivers; *(iv)* missense variants in all genes under positive selection and with dN/dS ratios for missense mutations above 5 (assuming 4 of every 5 missense mutations are drivers) labelled as likely drivers; or *(v)* in-frame indels in genes under significant positive selection for in-frame indels.

### Evolutionary analysis

Subclonal architecture reconstruction was performed using the DPClust R package v2.2.8^7,17^, after obtaining cancer cell fraction (CCF) estimates by dpclust3p v1.0.8 (https://github.com/Wedge-lab/dpclust3p) based on the variant allele frequency provided by the somatic variant callers and the copy number profiles determined by the BATTENBERG algorithm. Only tumors with at least 40% purity according to BATTENBERG were considered for further evolutionary analysis. For each tumor with at least one subclone, the respective somatic mutations were split into clonal and subclonal mutations using the most probable cluster assignment for each mutation as per the DPClust output. Mutations not assigned to a cluster by DPClust were removed from further analysis. Clusters centered at a CCF>1.5 and ones where chromosome X contributed the highest number of mutations were deemed artifactual, and the respective mutations were removed. Samples with a total number of clonal or subclonal mutations below 256 were also removed. Additionally, samples with poor separation between the clonal and subclonal distributions (*e.g.*, subclone centered at a CCF>0.80) were removed. Finally, only samples that had both a clone and at least one subclone post-filtering were retained for further analysis. This yielded a total of 223 samples, each with clonal and subclonal mutations. SigProfilerAssignment (v0.0.13) (https://github.com/AlexandrovLab/SigProfilerAssignment) was used to identify the activity of each mutational signature in each clone/subclone, and these activities were then normalized by the total number of mutations belonging to the clone/subclone (*i.e.*, clonal mutations were not included in the subclone). A two-sided Wilcoxon Signed-Rank Test^18^ was used to assess the differences in the relative activity of each mutational signature between the clones and their respective subclones. P-values were corrected using the Benjamini-Hochberg procedure^19^ and reported as q-values in the manuscript.

### Regressions

Signature attributions were dichotomized into presence and absence using confidence intervals, with presence defined as both lower and upper limits being positive, and absence as the lower limit being zero. If a signature was present in at least 75% of cases (SBS1, SBS40a, SBS40b, ID1, and ID5), it was dichotomized into above and below the median of attributed mutation counts. The binary attributions served as dependent variables in logistic regressions, and relevant risk factors were used as factorized independent variables. To adjust for confounding factors, sex, age of diagnosis, country, and tobacco status were added as covariates in regressions. The Bonferroni method was used to test for significant p-values (*i.e.,* a total of 224 comparisons for regressions with signatures, and a total of 24 comparisons for regressions with mutation burden). P-values reported are raw (not corrected). Regressions with incidence of renal cancer were performed as linear regressions with mutation burdens or signature attributions (those present in at least 75% of cases) with confidence intervals not consistent with zero as a dependent variable, and age-standardized rates (ASR) of renal cancer obtained from Global Cancer Observatory (GLOBOCAN)^20^, sex and age of diagnosis as independent variables. ASR of renal cancer for regions of Czech Republic were obtained from SVOD web portal^21^. Signatures present in less than 75% of cases were dichotomized into presence and absence as previously mentioned and analyzed using the logistic regressions. All regressions were performed on a sample basis.

### Polygenic risk score (PRS) analysis of lifestyle risk factors

In this analysis, we used the genome-wide association studies (GWAS) summary statistics estimated in European populations for well-established risk factors for ccRCC. For tobacco smoking status, we used results from the GSCAN consortium meta-analysis of smoking initiation (ever vs never status)^22^. For body mass index (BMI), the results of UK biobank (UKBB) and GIANT consortium meta-analysis of continuous BMI were used^23^. GWAS summary statistics related to hypertension, namely systolic blood pressure and diastolic blood pressure, as well as the ones related to diabetes^24^, such as fasting glucose and fasting insulin were also obtained using UKBB studies^25^.

Since all the GWAS summary statistics used in the current work were based on European populations, we used ADMIXTURE tool (v1.3.0)^26^ and principal component analysis (PCA) to infer the unsupervised cluster of individuals with European genetic background within ccRCC cases. Hapmap SNPs (n=1,176,821 variants) were extracted from the ccRCC whole-genome sequence genotype data. After basic quality control using PLINK (v1.9b, www.cog-genomics.org/plink/1.9/), 333 variants were removed due to missing genotype rate > 5%, 1,236 variants failed Hardy-Weinberg equilibrium test (p-values<10^-8^), and 18,702 variants had MAF<1% in our cohort. Additionally, 3 ambiguous variants and 21,358 variants within regions of long-range, high linkage disequilibrium (LD) in the human genome (hg38) were excluded. After pruning for linkage disequilibrium, 143,727 variants remained in ccRCC genotype data. The 1000 genome reference population genotype data (phase 3) for Europeans (N=489), Africans (YRI, N=108) and East Asians (N=103 from China and 104 from Japan) (https://www.internationalgenome.org/data/) were filtered and merged with ccRCC genotype data based on the pruned set of variants present in both datasets. ADMIXTURE was run on the merged genotype data with *k*=3, which would correspond to the three ancestral continental population groups that likely reflect the participants of our study. The ccRCC cases with European genetic fraction greater than 80% by the ADMIXTURE analysis were selected for the polygenic risk scores (PRS) analyses. To complement the ADMIXTURE analysis, PCA was run on the same samples.

The initial genotype data based on whole-genome sequence from 849 ccRCC cases with European genetic background consisted of biallelic SNPs with MAF >0.01% (to exclude ultra-rare variants; N ∼ 30 million variants). After basic quality control, variants with missing genotype rate of greater than 5% (N=7,519,196 variants) with strong deviation from Hardy-Weinberg equilibrium (p-values<10^-8^, N=220,862) were excluded. For each GWAS trait, we restricted our analyses to the biallelic SNPs with minor allele frequency (MAF) greater than 1% in the 1000 genomes reference for European populations. For the selection of the independent genome-wide significant hits (p-values<5 × 10^-8^) of each GWAS summary statistic used to generate the PRS, SNPs were clumped (r^2^=0.1 within a LD window of 10 MB) using PLINK (v1.9b, www.cog-genomics.org/plink/1.9/) based on the 1000 genomes European reference population genotype data (N=489; ∼ 10 million variants). Where a selected GWAS hit was not found in ccRCC genotype data, we extracted proxies (r^2^>0.8 in 1000 genomes) also present in ccRCC dataset where possible (**Supplementary Table 18**). The variance of each genetic trait explained by the genetic variants were calculated as previously suggested^27^. PRS was subsequently calculated as the sum of the individual’s beta-weighted genotypes using PRSice-2 software^28^. Associations were estimated per standard deviation increase in the PRS, which was normalized to have a mean of zero across ccRCC cases of European genetic ancestry.

### Untargeted metabolomics association with signatures

Of the 962 subjects from the main analysis, 901 subjects were included in this sub-study – all Japanese samples (*n*=36) as well as few cases from Czech Republic (*n*=13), Romania (*n*=5) and Russia (*n*=3) were not included due to lack of available plasma samples. Samples were randomized and analyzed as two independent analytical batches. Analysis was performed with a UHPLC-QTOF-MS system that consisted of a 1,290 Binary LC and a 6,550 QTOF mass spectrometer equipped with Jet Stream electrospray ionization source (Agilent Technologies), using previously described methods^29^. Pre-processing was performed using Profinder 10.0.2.162 and Mass Profiler Professional B.14.9.1 software (Agilent Technologies). A “Batch recursive feature extraction (small molecules)” process was employed for samples and blanks to find [M+H]^+^ ions. The two batches were processes separately and the resulting features were aligned in Mass Profiler Professional. Chromatographic peak areas were used as a measurement of intensity. No normalization or transformation of raw data was performed prior to the downstream data analysis.

A total of 2,392 features were detectable in at least one of the 901 samples. Features present in only one of the two batches were filtered out. Recursive filtering elimination was applied to decrease redundancy from highly correlated variables (r≥0.85, Pearson’s r calculated before any transformation/imputation) by selecting the features with least missing data within clusters of features. A total of 944 features were included in the statistical analysis. Features were pre-processed: missing values were replaced with 1/5 of the minimal value of the feature before applying mean centering and Pareto scaling. Each feature was regressed against both *de novo* and COSMIC signatures, adjusting for sex and age of diagnosis, as well as body mass index (BMI) and technical factors (batch, acquisition order) that could impact chromatographic peak area. Models for SBS22a and SBS22b were restricted to Romanian and Serbian samples to find potential pathways of Aristolochic acid exposure in the Balkan region. Logistic models were used for zero-inflated signatures (≥30% zeros) while quasi-Poisson regressions were used for the least zero-inflated signatures (SBS1, SBS40a, and SBS40b). To derive specific false detection rates, random variables were created from permutations of the initial features and regressed against signatures in the same fashion as true features. Maximum p-value thresholds from regressions with random features were compared to adjusted p-value thresholds according to Bonferroni’s procedure. The more conservative approach was used in selecting features of interest. Random forest models were also used as cross-checking multivariate models to assess the relative importance of each feature in explaining the signature attribution. As with univariate models, regression models were used for the least zero-inflated signatures (<30% of zeros) while classification models were used for all other signatures, with restriction to Romanian and Serbian samples for SBS22a and SBS22b. Importance was estimated from the total decrease in node impurities from splitting on the variable, averaged over all trees. Node impurity was measured by the Gini index for classification, and by residual sum of squares for regression. The significance of importance metrics for Random Forest models were estimated by permuting the response variable (https://github.com/EricArcher/rfPermute).

Features considered for identification, along with their highly correlated counterparts, were searched in Human Metabolome Database (HMDB), LipidMaps, Metlin, and Kegg. Compound identity was confirmed by comparison of retention times and MS/MS fragmentation against chemical standards when available, or otherwise against reference MS/MS spectra. Since the feature 240.1468@0.8929933 was strongly correlated with several features identified as TMAP (**Supplementary Table 11**), the integration of these features was inspected and corrected manually, and regressed against SBS40b using the same model applied to features selected for analysis. Creatinine was identified among the features by matching its retention time and MS/MS spectra against a reference standard and also regressed against SBS40b in the same fashion as other metabolites. Estimation of correlation between metabolic features was done using linear regression adjusting for batch and acquisition order.

### Targeted metabolomics analyses

Circulating levels of PFAS (Per- and Polyfluorinated Substances) and cystatin C compounds were investigated using targeted mass spectrometry-based methods as described previously^30,31^.

Out of the 962 subjects from the main analysis, plasma samples from 909 subjects (from all countries except Japan) were randomized and sent frozen in dry ice to each respective laboratory for analyses. Measurement of cystatin C from 906 subjects included its native form and isoforms (3Pro-OH cystatin C, cystatin C-desS, 3Pro-OH cystatin C-desS and cystatin C-desSSP) that were modeled individually and for the total concentration of cystatin C isoforms. Measurements of PFAS compounds included PFOA (Perfluorooctanoic Acid; total, branch, linear), PFOS (Perfluorooctanoic Acid; total, branch, linear), PFHxS (Perfluorohexane sulfonate), PFNA (Perfluorononanoic acid), PFDA (Perfluorodecanoic acid), MePFOSAA (n-methylperfluoro-1 octanesulfonamido acetic acid), EtPFOSAA (2-(N-Ethyl-perfluorooctane. sulfonamido) acetic acid).

Multivariable quasi-Poisson (for the least sparse signatures SBS1, SBS40a and SBS40b) and logistic regression were used to estimate the association between plasma concentrations of the aforementioned substances and mutational signatures. All compounds were modeled continuously (log2-transformed) and categorically, with adjustments made by sex, age, date of recruitment, country, BMI, tobacco and alcohol status in the case of PFAS molecules and by sex, age and BMI, in the case of cystatin C.

### Geospatial analyses

Geospatial analyses were performed to estimate the regional effect for signature attribution, particularly for signatures thought to be from exogenous exposure (SBS40b – unknown – and SBS22a/SB22b - Aristolochic acid). Residential history information was available for a large proportion of cases from the countries of interest: Czech Republic for SBS40b and Romania/Serbia for SBS22a/SBS22b. The 259 cases from Czech Republic within this study were recruited from 4 separate regions including Prague, České Budějovice (in Southern Bohemia), as well as Brno and Olomouc in the east of the country. Each individual residence was geocoded to its administrative region. All locations outside the country of recruitment were labeled as “Abroad”. A multi-membership mixed model was used to account for the full list of regions in which each subject resided, as well as the proportion of life spent in that region before diagnosis. As dependent variable, signatures were inverse-normal transformed. Models were adjusted for sex and age of diagnosis (fixed effects). The regional effect was treated as random effect.

## Data availability

Whole genome sequencing data and patient metadata are deposited in the European Genome-phenome Archive (EGA) associated with study EGAS00001003542. Aligned BAM files for all ccRCC cases included in the final analysis were deposited in dataset EGAD00001012102, variant calling files in dataset EGAD00001012222 and patient metadata in dataset EGAD00001012223. The metabolomics data are available upon request. All other data are provided in the accompanying **Supplementary Tables**.

## Code availability

All algorithms used for data analysis are publicly available with repositories noted within the respective method sections and in the accompanying reporting summary. Code used for regression analysis and figures is available at: https://gitlab.com/Mutographs/Mutographs_RCC.

## Acknowledgements

The authors would like to thank Laura O’Neill, Kirsty Roberts, Katie Smith, Maisie Farenden, Siobhan Austin-Guest and the staff of DNA Pipelines at the Wellcome Sanger Institute for their contribution. We are grateful for the support provided by Maja Milojevic, Christophe Lallemand, Helene Renard, Aude Bardot, Andreea Spanu and Nivonirina Robinot as well as IARC General Services, including the Laboratory Services and Biobank team led by Zisis Kozlakidis, the Section of Support to Research overseen by Tamas Landesz and the Evidence Synthesis and Classification Section led by Ian Cree, under IARC regular budget funding. The authors would like to thank Gislaine Bergo, Riley Cox and Juliana Oliveira for help with data/sample preparation and processing. The authors would also like to acknowledge the contributions of the Leeds Biobanking and Sample Processing Lab, the Leeds Multidisciplinary RTB and the Leeds NIHR BioRTB for provision of samples. The authors would like to thank Peter Campbell, Inigo Martincorena, Tim Butler, Daniela Mariosa, Laura Torrens Fontanals, Wellington Oliveira Dos Santos, Hana Zahed, Marc Gunter, Maggie Blanks and Mimi McCord for useful discussions. The authors would also like to thank all the patients and their families involved in this study.

## Contributions

The study was conceived, designed and supervised by M.R.S., P.B. and L.B.A. Analysis of data was performed by S.Senkin, S.Moody, M.D.-G., T.C., A.F.-I., J.W., S.F., M.K., R.V., A.P.L., E.N.B., A.K., B.O., S.C., E.T., J.A., K.S.-B., R.C.C.P., V.G., D.J., J.W.T. and J.M. Pathology review was carried out by B.A.-A., S.F. and M.A. Sample manipulation was carried out by C.L., C.C. and P.C. Patient and sample recruitment was led or facilitated by S.Sangkhathat, W.A., B.S., S.J., R.S., D.M., V.Jinga, S.R., S.Milosavljevic, M.M., S.Savic, J.M.S.B, M.A., L.P., P.A.-P., M.B., B.S.N., S.M.B., M.P.C., S.C.Z., R.M.R., E.F., N.S.M., R.S.F., R.B., N.V., D.Z., A.M., O.S., V.M., L.F., M.N., I.H., A.H., V.Janout, S.C. and C.L., M.P. P.K.-R., S.C., M.P., P.M.U. and M.J. contributed to data generation. Patient and sample recruitment for Japanese cases was led by T.S. and A.F. Scientific project management was carried out by L.H., E.C., G.S., A.C.D.C., A.F.-I. and S.P. S.Moody and S.Senkin jointly contributed and were responsible for overall scientific coordination. The manuscript was written by S.Senkin, S.Moody, M.R.S. and P.B. with contributions from all other authors.

## Disclaimer

Where authors are identified as personnel of the International Agency for Research on Cancer/ World Health Organization, the authors alone are responsible for the views expressed in this article and they do not necessarily represent the decisions, policy or views of the International Agency for Research on Cancer / World Health Organization.

## Corresponding author

Correspondence to Paul Brennan.

## EXTENDED DATA FIGURE AND TABLE LEGENDS

**Extended Data Fig. 1:**
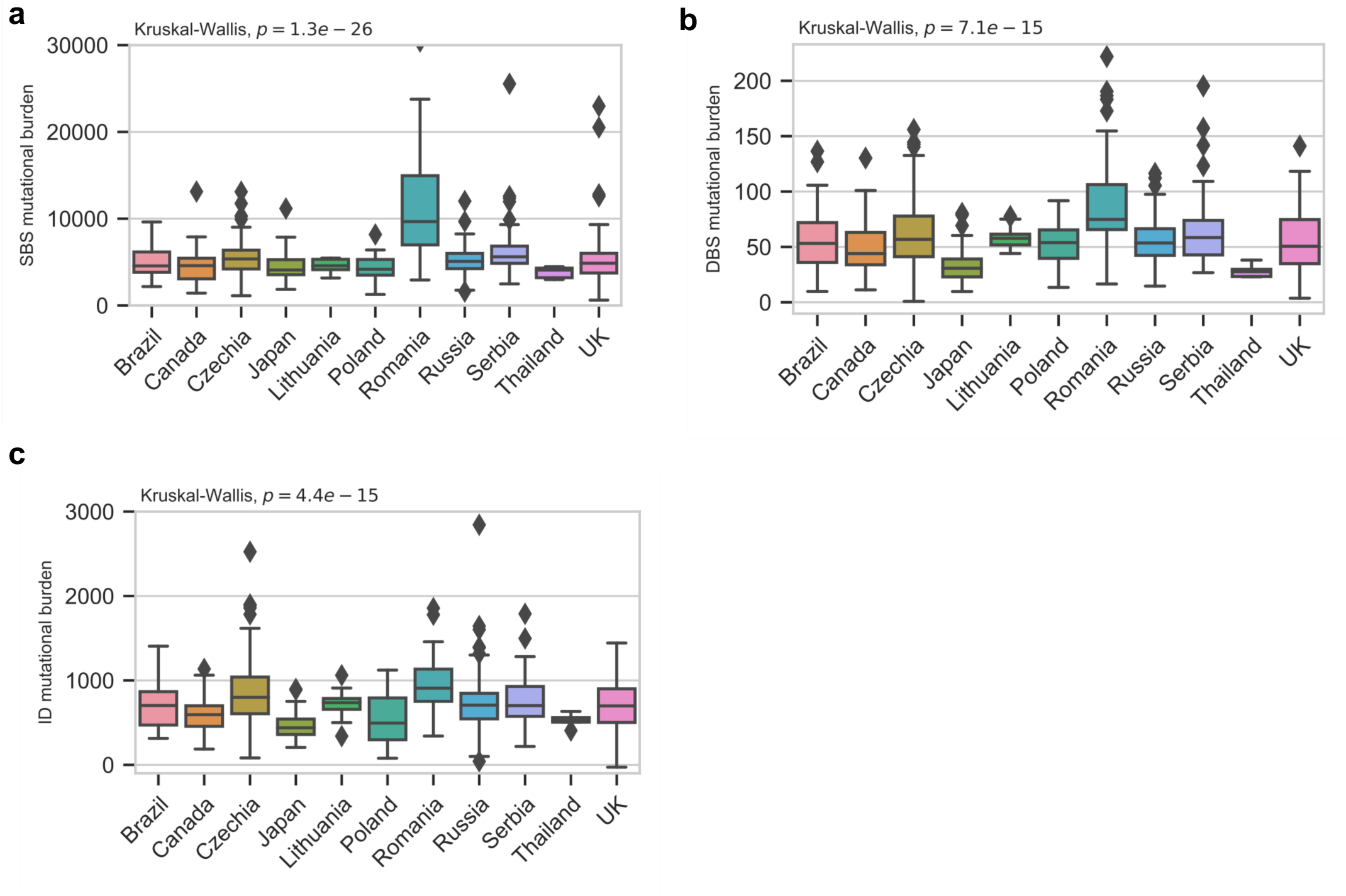
Mutation burdens in clear cell renal cell carcinomas across countries. Mutation burdens for single base substitutions (SBS) **(a)**, doublet base substitutions (DBS) **(b)** and small insertions and deletions (ID) **(c)** show significant differences between countries using the Kruskal-Wallis (two-sided) test (n=961 biologically independent samples). Four SBS hypermutators and four ID hypermutators above mutation burden of 30000 and 3000, respectively, were removed for clarity. Box and whiskers plots are in the style of Tukey. The line within the box is plotted at the median while the upper and lower ends are indicated 25th and 75th percentiles. Whiskers show 1.5*IQR (interquartile range) and values outside it are shown as individual data points.

**Extended Data Fig. 2:**
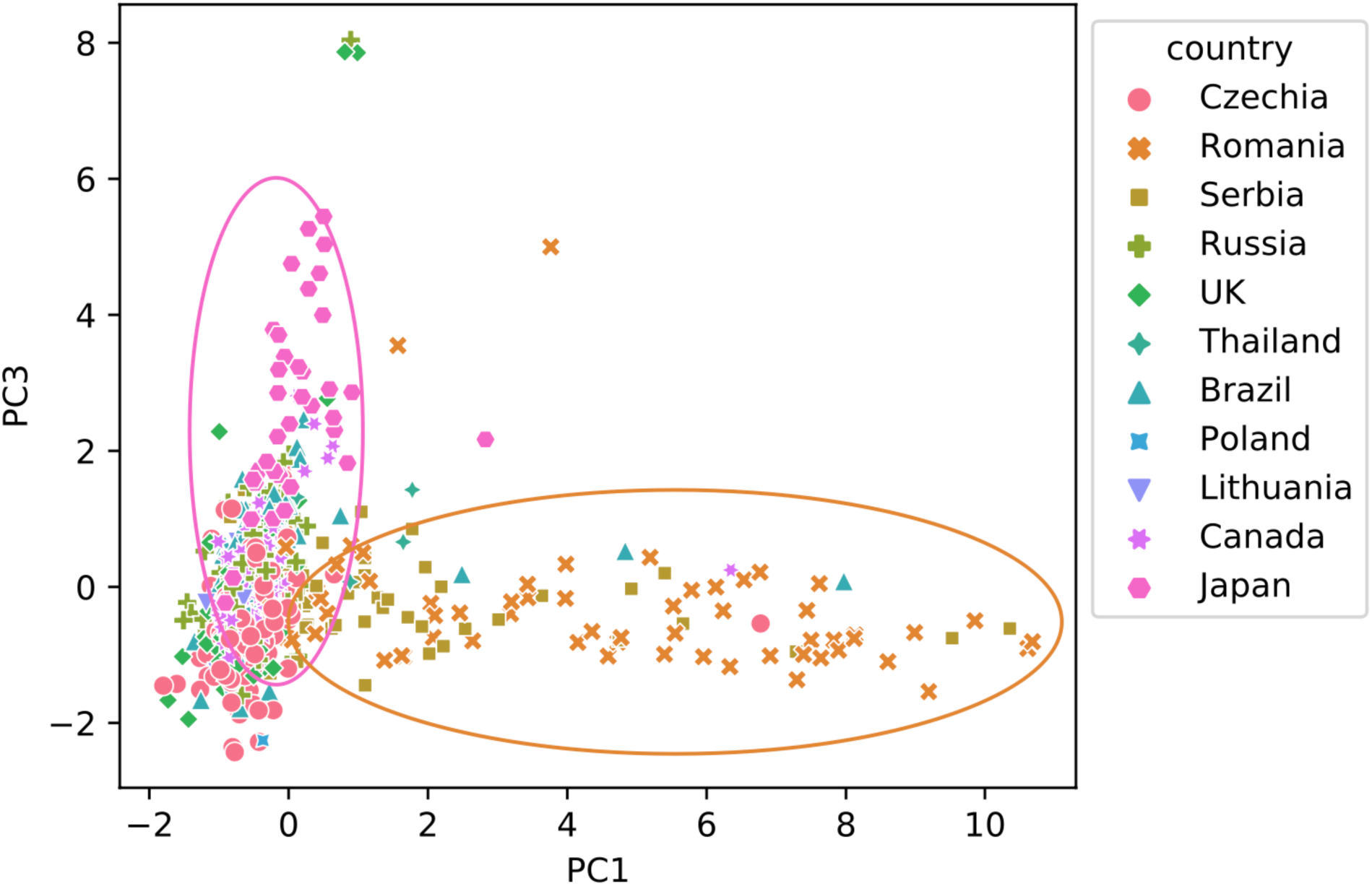
Principal component analysis of relative mutation counts. PCA performed on relative mutation counts of all ccRCC tumors incorporating the six mutation classes (C>A, C>G, C>T, T>A, T>C, T>G). Principal component 1 (PC1) clearly separates the cluster of mostly Romanian cases that are enriched with AA signatures, often at high mutation burdens. Principal component 3 (PC3) identifies a cluster of mostly Japanese cases, enriched with signature SBS12.

**Extended Data Fig 3:**
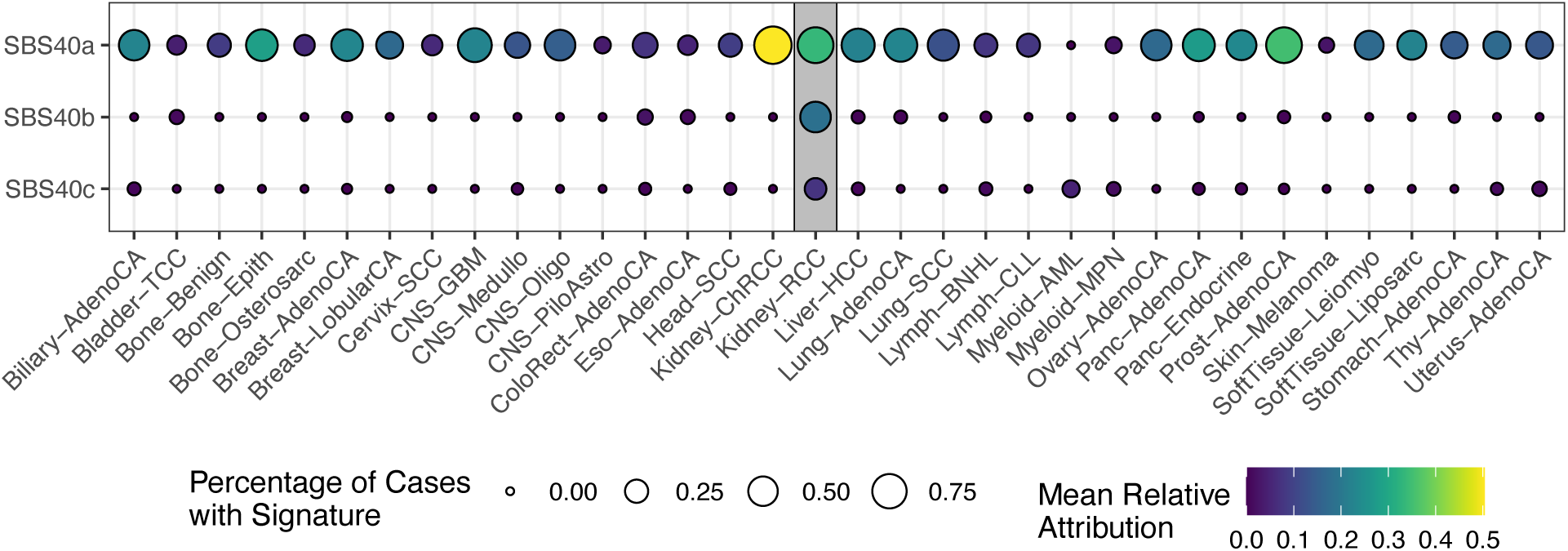
Attribution of signatures SBS40a, SBS40b, and SBS40c in a pan-cancer cohort. Attribution of signatures SBS40a, SBS40b, and SBS40c in a pan-cancer cohort, showing a widespread distribution for SBS40a whilst SBS40b and SBS40c are only seen consistently in clear cell renal cell carcinomas (ccRCC). The size of each dot represents the proportion of samples of each tumor type where the signature is present. The color of each dot represents the average mutation burden.

**Extended Data Fig. 4:**
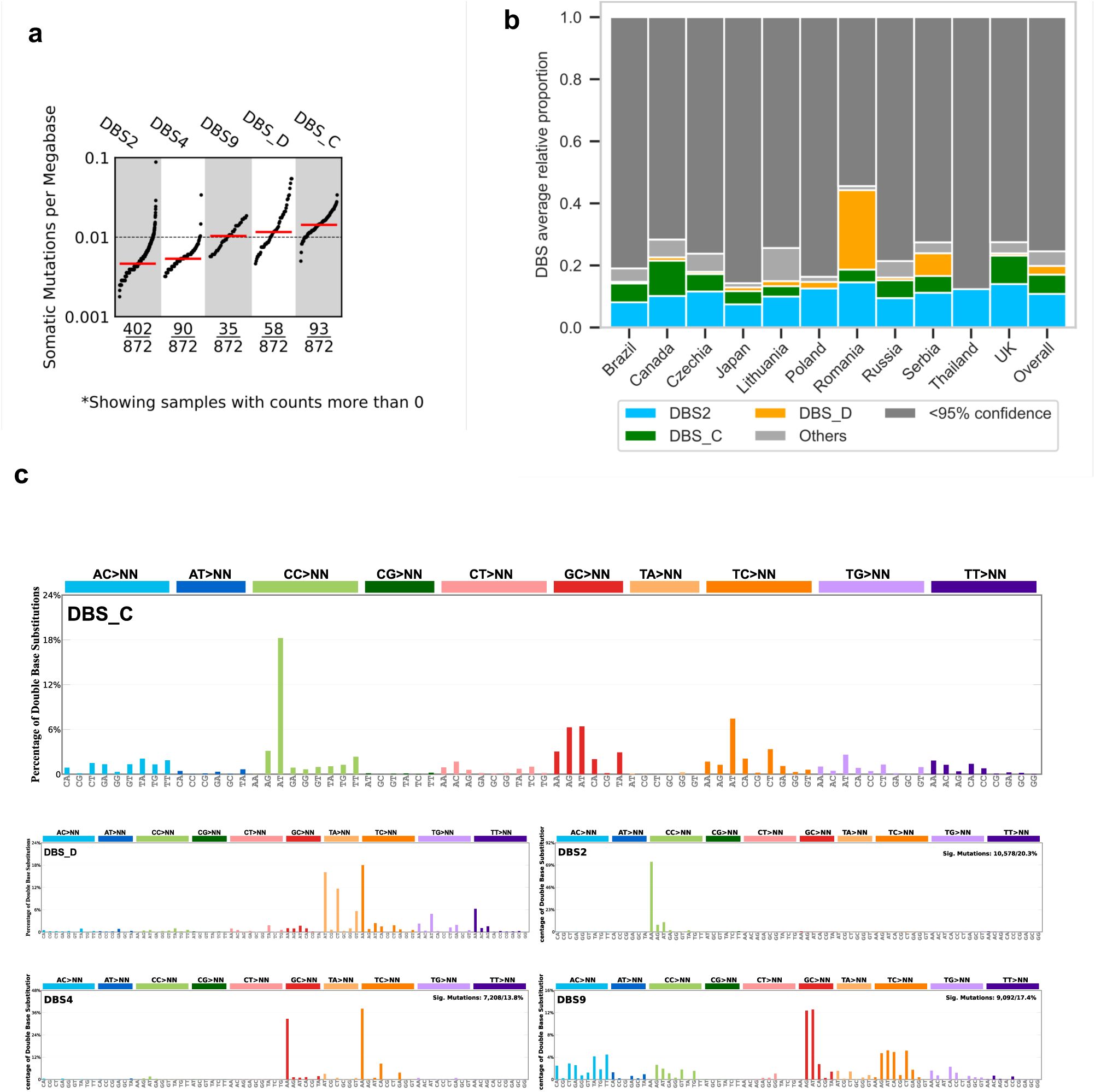
Doublet-base substitution signatures operative in clear cell renal cell carcinomas. **(a)** Tumour mutation burden (TMB) plot showing the frequency and mutations per Mb for each of the decomposed DBS signatures. **(b)** Average relative attribution for doublet-base substitution (DBS) signatures across countries. Signatures contributing less than 5% on average are grouped in the ‘Other’ category, apart from signature DBS_D. Category named ‘<95% confidence’ accounts for the proportion of mutation burden which could not be assigned to any signature with confidence level of at least 95%. **(c)** Decomposed DBS signatures, including reference COSMIC signatures as well as *de novo* signatures not decomposed into COSMIC reference signatures.

**Extended Data Fig. 5:**
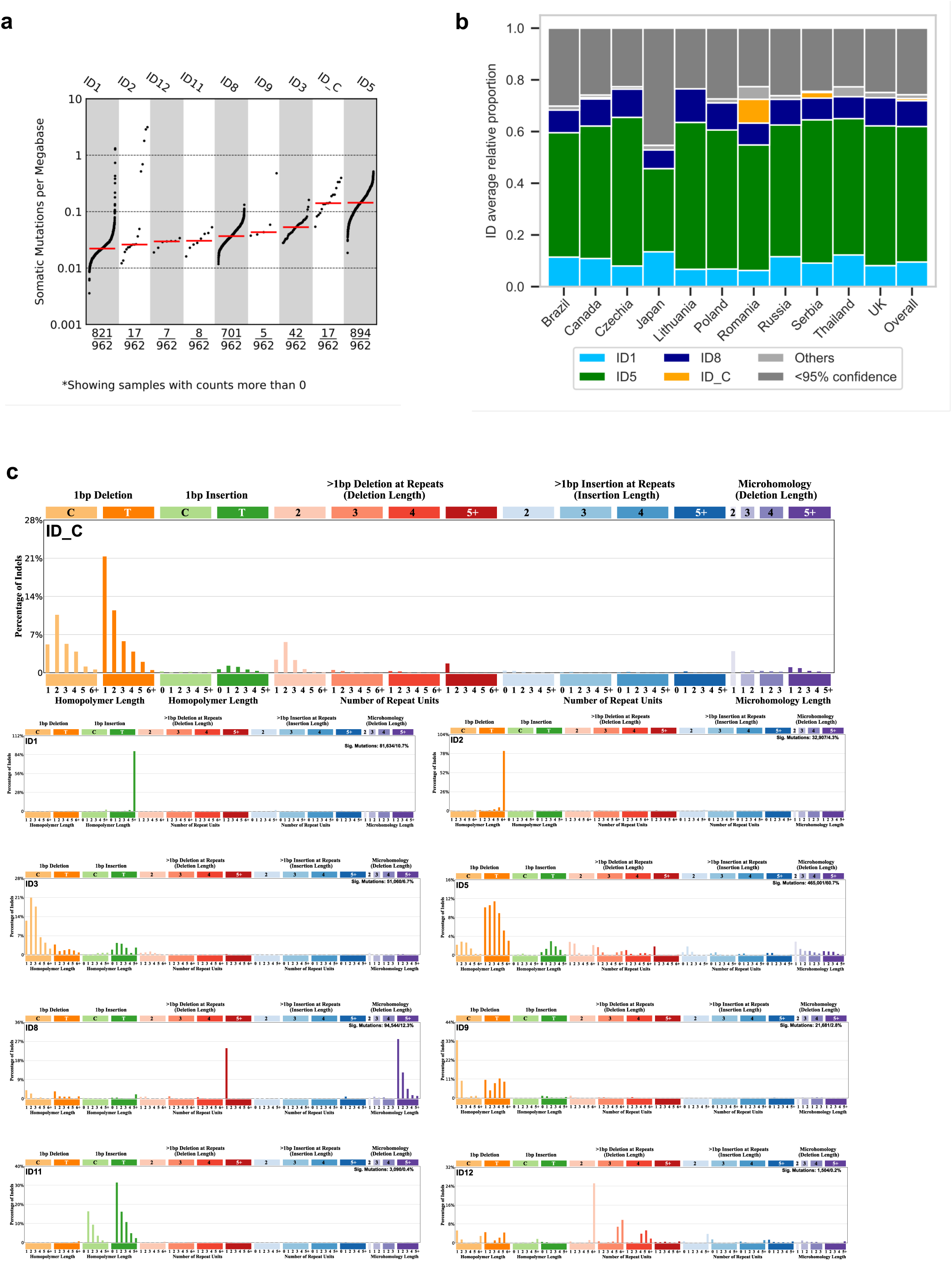
Small insertions and deletion signatures operative in clear cell renal cell carcinomas. **(a)** Tumour mutation burden (TMB) plot showing the frequency and mutations per Mb for each of the decomposed ID signatures. **(b)** Average relative attribution for small insertion and deletion (ID) signatures across countries. Signatures contributing less than 5% on average are grouped in the ‘Others’ category, apart from signature ID_C. Category named ‘<95% confidence’ accounts for the proportion of mutation burden which could not be assigned to any signature with confidence level of at least 95%. **(c)** Decomposed ID signatures, including reference COSMIC signatures as well as *de novo* signatures not decomposed into COSMIC reference signatures.

**Extended Data Fig. 6:**
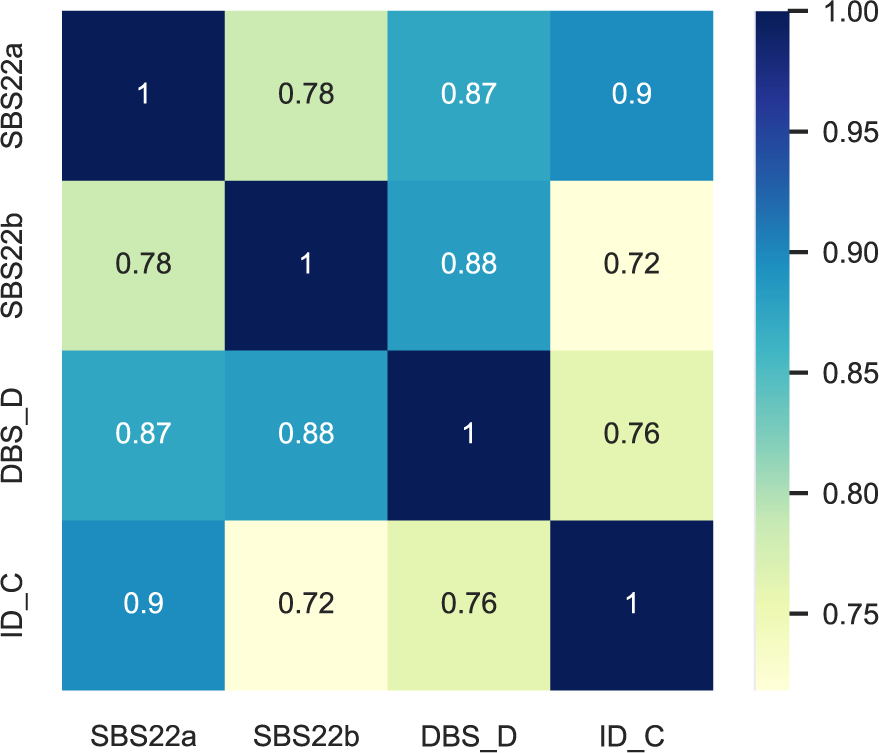
Correlation amongst signatures SBS22a, SBS22b, DBS_D, ID_C.

**Extended Data Table 1:** Presence of signatures SBS22a, SBS22b, DBS_D, ID_C across countries.

| Country | N cases | SBS22a (%) | SBS22b (%) | DBS_D (%) | ID_C (%) | SBS22a or SBS22b (%) | Any (%) |
| --- | --- | --- | --- | --- | --- | --- | --- |
| Romania | 64 | 45 (70.3) | 48 (75.0) | 42 (65.6) | 13 (20.3) | 53 (82.8) | 54 (84.4) |
| Serbia | 69 | 16 (23.2) | 33 (47.8) | 11 (15.9) | 3 (4.3) | 35 (50.7) | 36 (52.2) |
| Thailand | 5 | 3 (60.0) | 3 (60.0) | 0 (0.0) | 0 (0.0) | 4 (80.0) | 4 (80.0) |
| Brazil | 96 | 3 (3.1) | 3 (3.1) | 1 (1.0) | 0 (0.0) | 3 (3.1) | 3 (3.1) |
| Canada | 73 | 2 (2.7) | 0 (0.0) | 2 (2.7) | 1 (1.4) | 2 (2.7) | 3 (4.1) |
| Czechia | 259 | 1 (0.4) | 5 (1.9) | 32 (12.4) | 0 (0.0) | 6 (2.3) | 37 (14.3) |
| UK | 115 | 1 (0.9) | 0 (0.0) | 31 (27.0) | 0 (0.0) | 1 (0.9) | 31 (27.0) |
| Russia | 216 | 0 (0.0) | 1 (0.5) | 26 (12.0) | 0 (0.0) | 1 (0.5) | 27 (12.5) |
| Poland | 13 | 0 (0.0) | 0 (0.0) | 1 (7.7) | 0 (0.0) | 0 (0.0) | 1 (7.7) |
| Lithuania | 16 | 0 (0.0) | 1 (6.2) | 1 (6.2) | 0 (0.0) | 1 (6.2) | 2 (12.5) |
| Japan | 36 | 0 (0.0) | 1 (2.8) | 1 (2.8) | 0 (0.0) | 1 (2.8) | 1 (2.8) |

**Extended Data Fig. 7:**
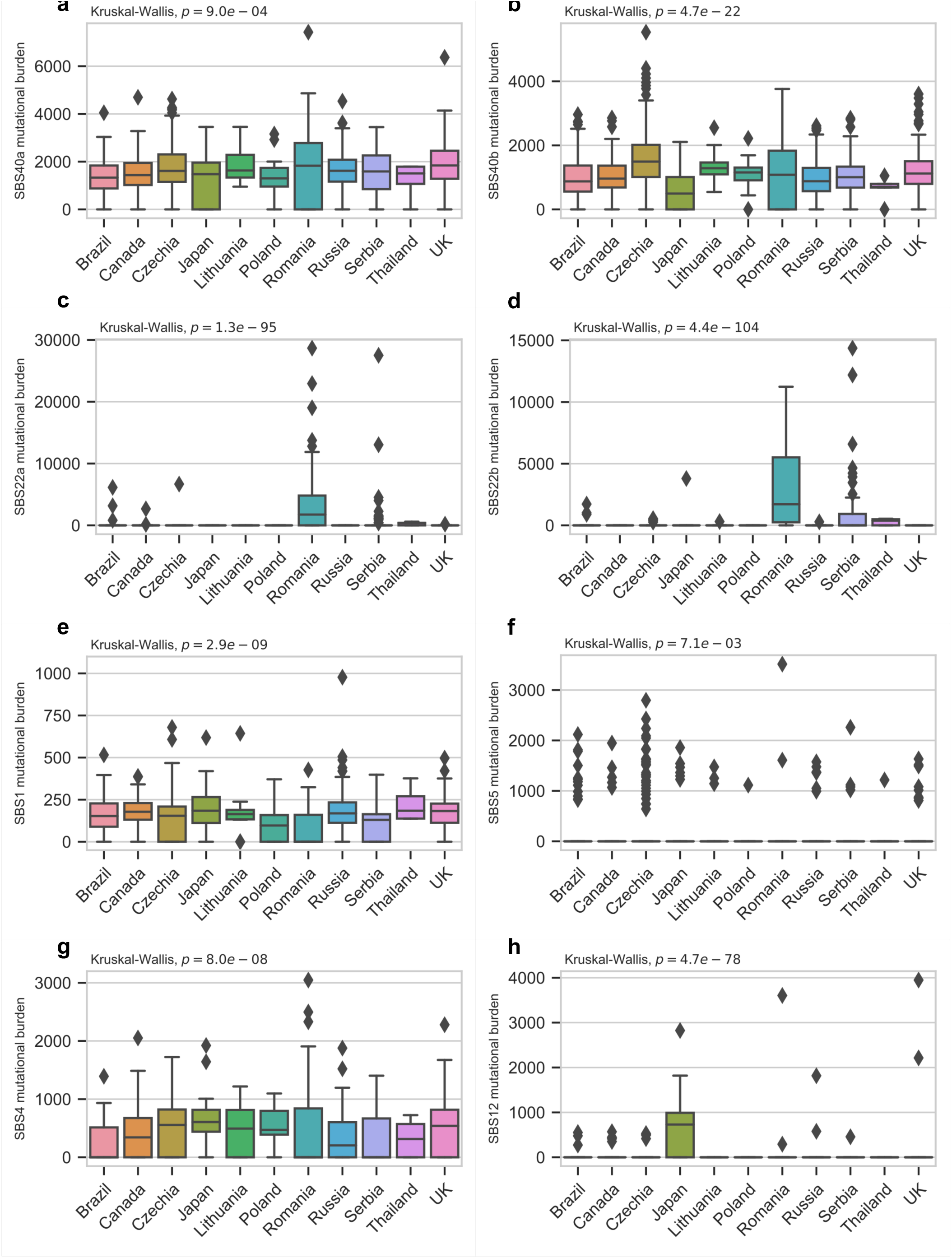
Single base substitution signatures showing significant differences in attributed mutation burden between countries. Signatures SBS40a **(a)** and SBS40b **(b)** were more prevalent in high-incidence regions of Czech Republic and Lithuania. Signatures SBS22a **(c)** and SBS22b **(d)** were enriched in Romania and Serbia. SBS1 **(e)**, SBS5 **(f)** and SBS4 **(g)** showed moderate differences across countries. Signature SBS12 **(h)** is highly prevalent in Japan. Five SBS1 hypermutators above mutation burden of 1000 were removed for clarity. Box and whiskers plots are in the style of Tukey. The line within the box is plotted at the median while the upper and lower ends are indicated 25th and 75th percentiles. Whiskers show 1.5*IQR (interquartile range) and values outside it are shown as individual data points.

**Extended Data Fig. 8:**
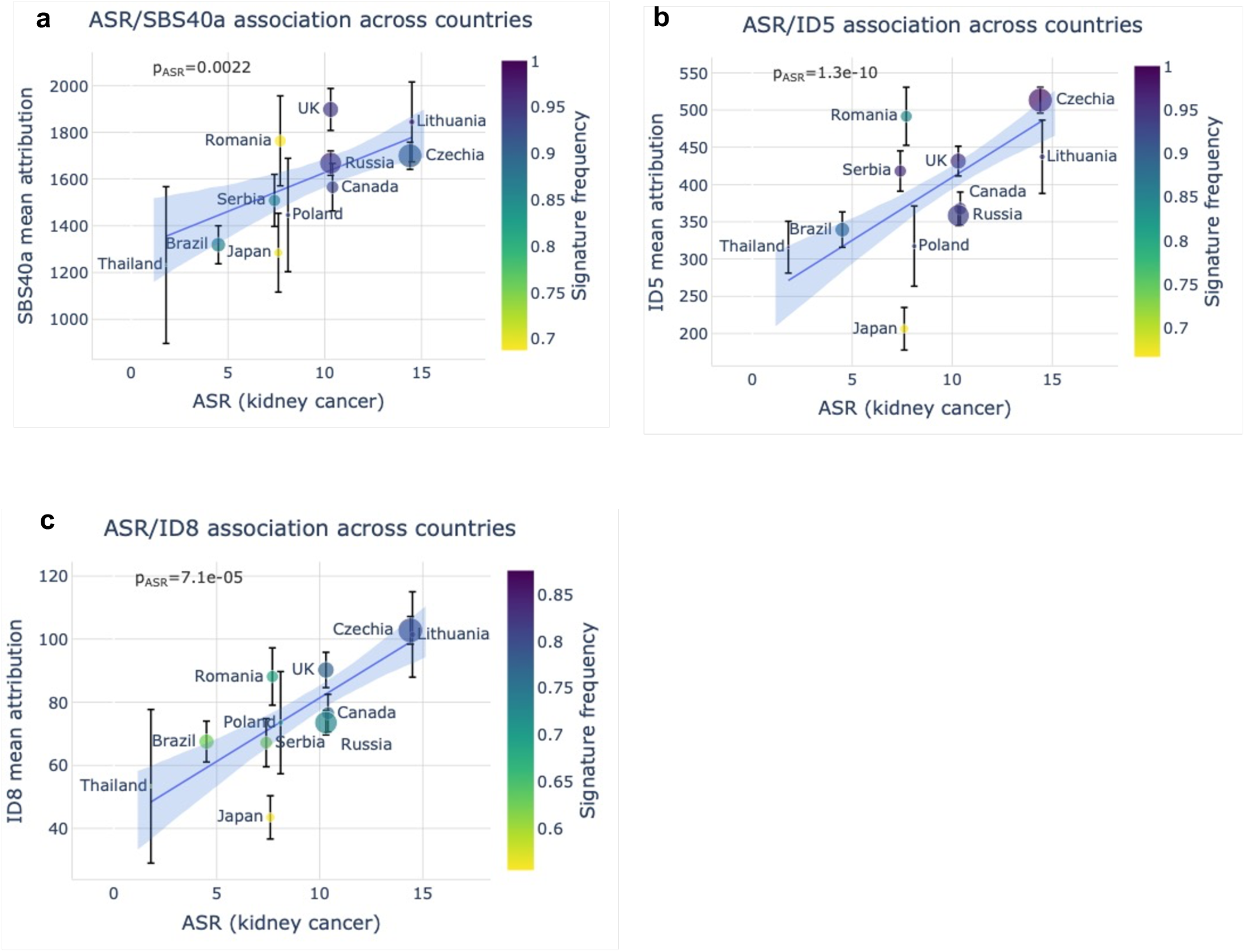
Association of mutational signatures with incidence of renal cancer. Number of mutations attributed to signatures **(a)** SBS40a, **(b)** ID5 and **(c)** ID8 against age-standardized incidence rate (ASR) of kidney cancer in each of the eleven countries represented in the cohort. Error bars represent standard errors of the mean. The p-values shown are for the ASR variable in linear regressions across all cases, adjusted for sex and age of diagnosis.

**Extended Data Fig. 9:**
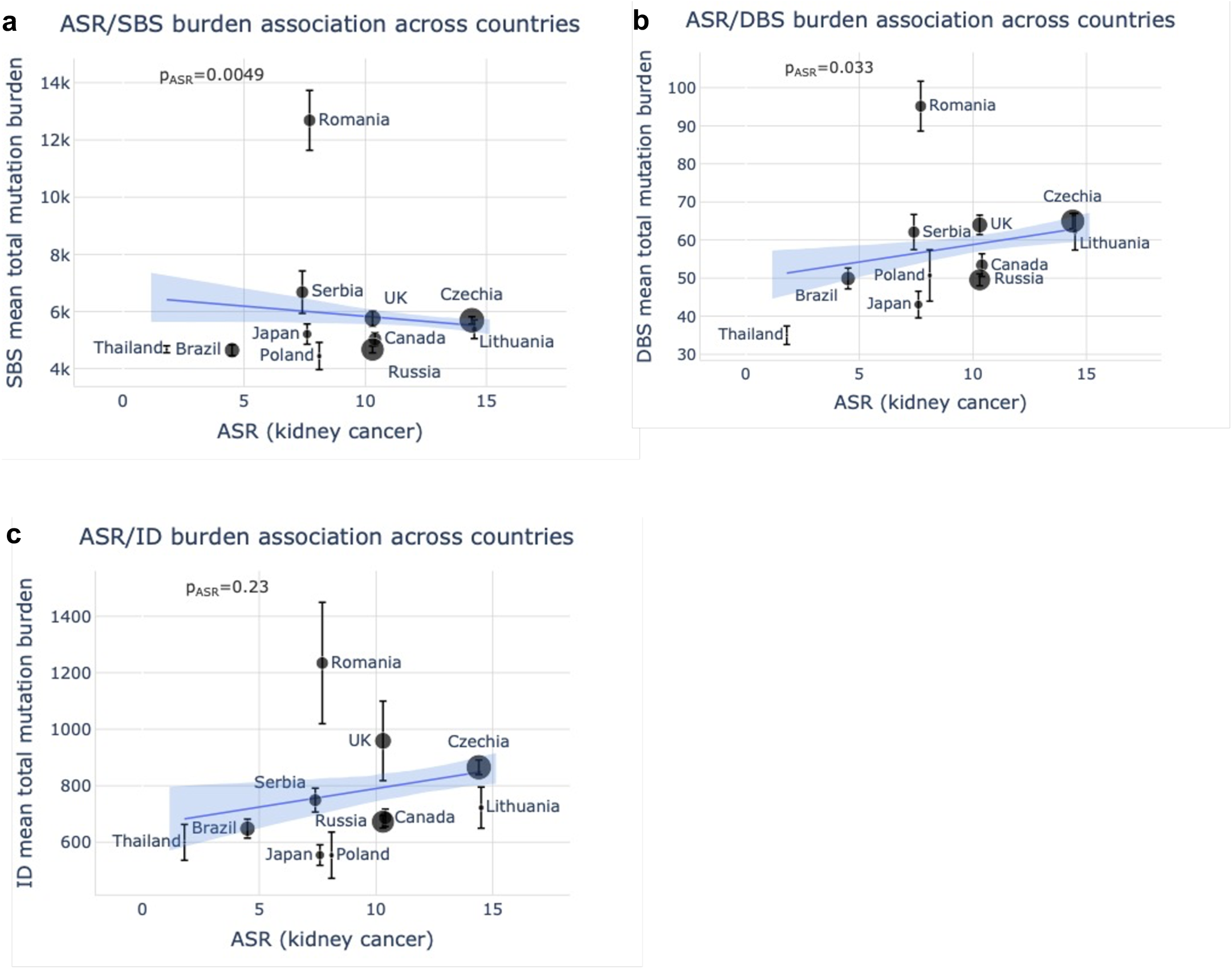
Association of mutation burden with incidence of renal cancer. Association of age-standardized rates (ASR) of kidney cancer incidence with SBS (**a)**, DBS (**b)** and ID (**c)** mutation burdens across countries. Error bars represent standard errors of the mean. The p-values shown are for the ASR variable in linear regressions across all cases, adjusted for sex and age of diagnosis.

**Extended Data Fig. 10:**
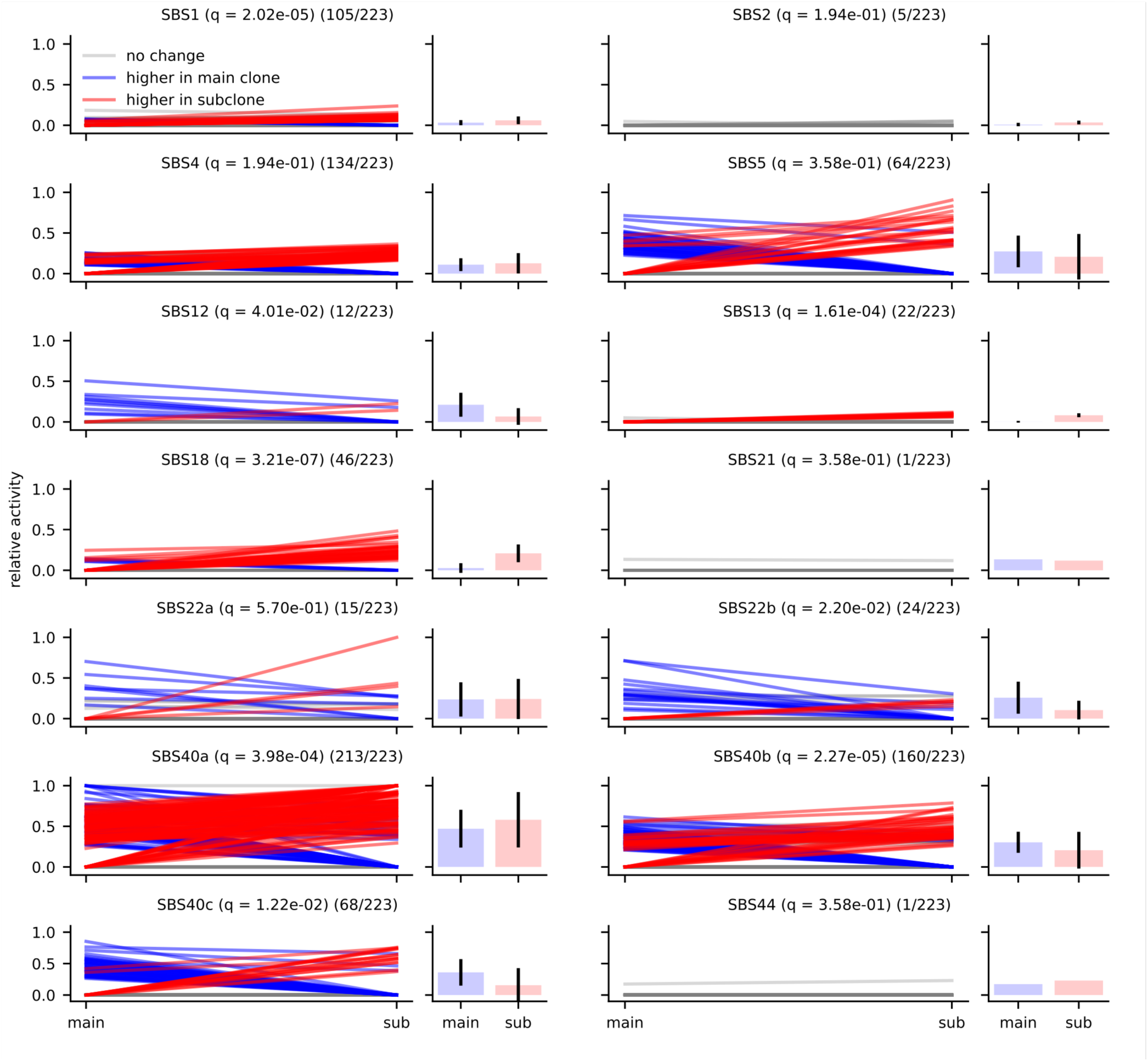
Evolutionary analysis of mutational signatures in ccRCC. Comparison of mutational signatures between clonal and subclonal mutations. Lines show the change in relative activity between the clonal mutations (main) and subclonal mutations (sub) within a sample. Blue and red lines represent an activity change of more than 6% (blue indicates higher in the clonal mutations; red indicates higher in the subclonal mutations). Bar plots show the distribution of activities in samples where the signature was present in the clonal and/or subclonal mutations; this number is represented in the title of each plot as X/223 for each signature. Black bars indicate one standard deviation away from the mean. Significance was assessed using a two-sided Wilcoxon signed-rank test, and q-values were generated using the Benjamini-Hochberg Procedure.

## REFERENCES

1. Brennan, P. & Davey-Smith, G. Identifying Novel Causes of Cancers to Enhance Cancer Prevention: New Strategies Are Needed. JNCI: Journal of the National Cancer Institute 114, 353–360 (2022).

2. Hsieh, J. J. et al. Renal cell carcinoma. Nat Rev Dis Primers 3, 17009 (2017).

3. Koh, G., Degasperi, A., Zou, X., Momen, S. & Nik-Zainal, S. Mutational signatures: emerging concepts, caveats and clinical applications. Nat Rev Cancer 21, 619–637 (2021).

4. Alexandrov, L. B. et al. The repertoire of mutational signatures in human cancer. Nature 578, 94–101 (2020).

5. Scelo, G. et al. Variation in genomic landscape of clear cell renal cell carcinoma across Europe. Nat Commun 5, 5135 (2014).

6. Mitchell, T. J. et al. Timing the Landmark Events in the Evolution of Clear Cell Renal Cell Cancer: TRACERx Renal. Cell 173, 611–623.e17 (2018).

7. Campbell, P. J. et al. Pan-cancer analysis of whole genomes. Nature 578, 82–93 (2020).

8. Degasperi, A. et al. A practical framework and online tool for mutational signature analyses show intertissue variation and driver dependencies. Nat Cancer 1, 249–263 (2020).

9. The Cancer Genome Atlas Research Network. Comprehensive molecular characterization of clear cell renal cell carcinoma. Nature 499, 43–49 (2013).

10. Mutographs Cancer Grand Challenge. https://cancergrandchallenges.org/teams.

11. Sung, H. et al. Global Cancer Statistics 2020: GLOBOCAN Estimates of Incidence and Mortality Worldwide for 36 Cancers in 185 Countries. CA Cancer J Clin 71, 209– 249 (2021).

12. Nik-Zainal, S. et al. Mutational Processes Molding the Genomes of 21 Breast Cancers. Cell 149, 979–993 (2012).

13. Alexandrov, L. B. et al. Signatures of mutational processes in human cancer. Nature 500, 415–421 (2013).

14. Drost, J. et al. Use of CRISPR-modified human stem cell organoids to study the origin of mutational signatures in cancer. Science (1979) 358, 234–238 (2017).

15. Hoang, M. L. et al. Mutational Signature of Aristolochic Acid Exposure as Revealed by Whole-Exome Sequencing. Sci Transl Med 5, (2013).

16. Poon, S. L. et al. Genome-wide mutational signatures of aristolochic acid and its application as a screening tool. Sci Transl Med 5, 197ra101 (2013).

17. Grollman, A. P. Aristolochic acid nephropathy: Harbinger of a global iatrogenic disease. Environ Mol Mutagen 54, 1–7 (2013).

18. Turesky, R. J. et al. Aristolochic acid exposure in Romania and implications for renal cell carcinoma. Br J Cancer 114, 76–80 (2016).

19. Wang, X.-M. et al. Integrative genomic study of Chinese clear cell renal cell carcinoma reveals features associated with thrombus. Nat Commun 11, 739 (2020).

20. Huang, M. N. et al. Genome-scale mutational signatures of aflatoxin in cells, mice, and human tumors. Genome Res 27, 1475–1486 (2017).

21. Haradhvala, N. J. et al. Mutational Strand Asymmetries in Cancer Genomes Reveal Mechanisms of DNA Damage and Repair. Cell 164, 538–549 (2016).

22. Nik-Zainal, S. et al. The genome as a record of environmental exposure. Mutagenesis gev073 (2015) doi:10.1093/mutage/gev073.

23. Sato, Y. et al. Integrated molecular analysis of clear-cell renal cell carcinoma. Nat Genet 45, 860–867 (2013).

24. Dempsey, D. et al. Nicotine metabolite ratio as an index of cytochrome P450 2A6 metabolic activity. Clin Pharmacol Ther 76, 64–72 (2004).

25. Velenosi, T. J. et al. Untargeted metabolomics reveals N, N, N-trimethyl-L-alanyl-L-proline betaine (TMAP) as a novel biomarker of kidney function. Sci Rep 9, 6831 (2019).

26. Sato, Y. et al. Integrated molecular analysis of clear-cell renal cell carcinoma. Nat Genet 45, 860–867 (2013).

27. Nik-Zainal, S. et al. The life history of 21 breast cancers. Cell 149, 994–1007 (2012).

28. Dentro, S. C., Wedge, D. C. & van Loo, P. Principles of Reconstructing the Subclonal Architecture of Cancers. Cold Spring Harb Perspect Med 7, (2017).

29. Shearer, J. J. et al. Serum Concentrations of Per- and Polyfluoroalkyl Substances and Risk of Renal Cell Carcinoma. J Natl Cancer Inst 113, 580–587 (2021).

30. Nik-Zainal, S. et al. The genome as a record of environmental exposure. Mutagenesis gev073 (2015) doi:10.1093/mutage/gev073.

31. Kucab, J. E. et al. A Compendium of Mutational Signatures of Environmental Agents. Cell 177, 821–836.e16 (2019).

32. Gabriel, A. A. G. et al. Genetic Analysis of Lung Cancer and the Germline Impact on Somatic Mutation Burden. JNCI: Journal of the National Cancer Institute 114, 1159– 1166 (2022).

33. Liu, Y., Gusev, A., Heng, Y. J., Alexandrov, L. B. & Kraft, P. Somatic mutational profiles and germline polygenic risk scores in human cancer. Genome Med 14, 14 (2022).

34. Moody, S. et al. Mutational signatures in esophageal squamous cell carcinoma from eight countries with varying incidence. Nat Genet 53, 1553–1563 (2021).

35. Abascal, F. et al. Somatic mutation landscapes at single-molecule resolution. Nature 593, 405–410 (2021).

36. Martincorena, I. et al. High burden and pervasive positive selection of somatic mutations in normal human skin. Science (1979) 348, 880–886 (2015).

37. Martincorena, I. et al. Somatic mutant clones colonize the human esophagus with age. Science (1979) 362, 911–917 (2018).

38. Fowler, J. C. & Jones, P. H. Somatic Mutation: What Shapes the Mutational Landscape of Normal Epithelia? Cancer Discov 12, 1642–1655 (2022).

## Methods references

1. Moody, S. et al. Mutational signatures in esophageal squamous cell carcinoma from eight countries with varying incidence. Nat Genet 53, 1553–1563 (2021).

2. Scelo, G. et al. Variation in genomic landscape of clear cell renal cell carcinoma across Europe. Nat Commun 5, 5135 (2014).

3. Campbell, P. J. et al. Pan-cancer analysis of whole genomes. Nature 578, 82–93 (2020).

4. Whalley, J. P. et al. Framework for quality assessment of whole genome cancer sequences. Nat Commun 11, 5040 (2020).

5. Bergmann, E. A., Chen, B.-J., Arora, K., Vacic, V. & Zody, M. C. Conpair: concordance and contamination estimator for matched tumor–normal pairs. Bioinformatics 32, 3196–3198 (2016).

6. Van Loo, P. et al. Allele-specific copy number analysis of tumors. Proceedings of the National Academy of Sciences 107, 16910–16915 (2010).

7. Nik-Zainal, S. et al. The life history of 21 breast cancers. Cell 149, 994–1007 (2012).

8. Jones, D. et al. cgpCaVEManWrapper: Simple Execution of CaVEMan in Order to Detect Somatic Single Nucleotide Variants in NGS Data. Curr Protoc Bioinformatics 56, (2016).

9. Raine, K. M. et al. cgpPindel: Identifying Somatically Acquired Insertion and Deletion Events from Paired End Sequencing. Curr Protoc Bioinformatics 52, (2015).

10. Kim, S. et al. Strelka2: fast and accurate calling of germline and somatic variants. Nat Methods 15, 591–594 (2018).

11. Bergstrom, E. N. et al. SigProfilerMatrixGenerator: a tool for visualizing and exploring patterns of small mutational events. BMC Genomics 20, 685 (2019).

12. Liu, M., Wu, Y., Jiang, N., Boot, A. & Rozen, S. G. mSigHdp: hierarchical Dirichlet process mixture modeling for mutational signature discovery. bioRxiv 2022.01.31.478587 (2022) doi:10.1101/2022.01.31.478587.

13. Islam, S. M. A. et al. Uncovering novel mutational signatures by de novo extraction with SigProfilerExtractor. Cell genomics 2, None (2022).

14. Alexandrov, L. B. et al. The repertoire of mutational signatures in human cancer. Nature 578, 94–101 (2020).

15. Senkin, S. MSA: reproducible mutational signature attribution with confidence based on simulations. BMC Bioinformatics 22, 540 (2021).

16. Martincorena, I. et al. Universal Patterns of Selection in Cancer and Somatic Tissues. Cell 171, 1029–1041.e21 (2017).

17. Dentro, S. C., Wedge, D. C. & van Loo, P. Principles of Reconstructing the Subclonal Architecture of Cancers. Cold Spring Harb Perspect Med 7, (2017).

18. Wilcoxon, F. Individual Comparisons by Ranking Methods. Biometrics Bulletin 1, 80 (1945).

19. Benjamini, Y. & Hochberg, Y. Controlling the False Discovery Rate: A Practical and Powerful Approach to Multiple Testing. Journal of the Royal Statistical Society: Series B (Methodological) 57, 289–300 (1995).

20. Sung, H. et al. Global Cancer Statistics 2020: GLOBOCAN Estimates of Incidence and Mortality Worldwide for 36 Cancers in 185 Countries. CA Cancer J Clin 71, 209– 249 (2021).

21. Dušek, L. et al. Epidemiology of Malignant Tumours in the Czech Republic [online]. Masaryk University, Czech Republic, [2005]. http://www.svod.cz/. *Version 7.0 [2007], ISSN 1802 –* 8861.

22. Liu, M. et al. Association studies of up to 1.2 million individuals yield new insights into the genetic etiology of tobacco and alcohol use. Nat Genet 51, 237–244 (2019).

23. Yengo, L. et al. Meta-analysis of genome-wide association studies for height and body mass index in ∼700000 individuals of European ancestry. Hum Mol Genet 27, 3641– 3649 (2018).

24. Lagou, V. et al. Sex-dimorphic genetic effects and novel loci for fasting glucose and insulin variability. Nat Commun 12, 24 (2021).

25. Bycroft, C. et al. The UK Biobank resource with deep phenotyping and genomic data. Nature 562, 203–209 (2018).

26. Alexander, D. H., Novembre, J. & Lange, K. Fast model-based estimation of ancestry in unrelated individuals. Genome Res 19, 1655–1664 (2009).

27. Shim, H. et al. A Multivariate Genome-Wide Association Analysis of 10 LDL Subfractions, and Their Response to Statin Treatment, in 1868 Caucasians. PLoS One 10, e0120758 (2015).

28. Choi, S. W. & O’Reilly, P. F. PRSice-2: Polygenic Risk Score software for biobank-scale data. Gigascience 8, (2019).

29. Loftfield, E. et al. Novel Biomarkers of Habitual Alcohol Intake and Associations With Risk of Pancreatic and Liver Cancers and Liver Disease Mortality. JNCI: Journal of the National Cancer Institute 113, 1542–1550 (2021).

30. Shearer, J. J. et al. Serum Concentrations of Per- and Polyfluoroalkyl Substances and Risk of Renal Cell Carcinoma. J Natl Cancer Inst 113, 580–587 (2021).

31. Gao, J., Meyer, K., Borucki, K. & Ueland, P. M. Multiplex Immuno-MALDI-TOF MS for Targeted Quantification of Protein Biomarkers and Their Proteoforms Related to Inflammation and Renal Dysfunction. Anal Chem 90, 3366–3373 (2018).

