## Supplementary Note for "Geographic variation of mutagenic exposures in kidney cancer genomes"

**Supplementary Information Inventory**

**This file contains the following:**

Supplementary Note

Supplementary References

Supplementary Figures

#### SUPPLEMENTARY NOTE

**Contents**


**1.Mutational signature analysis**

1.1 Extraction of *de novo* mutational signatures with SigProfilerExtractor

1.2 Extraction of *de novo* signatures with mSigHdp

1.3 Decomposition to reference signatures

1.4 Justification for non-decomposed signatures

1.5 Exploration of conditions required for SBS40a, SBS40b and SBS40c separation

1.6 Presence of tobacco-associated signature SBS4

1.7 Attribution of single base substitution signatures

**2. Attribution of mutational signatures in external data sets**

2.1 Attribution of SBS40 components in a pan-cancer cohort

2.2 Attribution of SBS12 in liver cancers

2.3 Validation of SBS12 in additional datasets

**3. Tumour Mutation Burdens and regression analysis**

3.1 Adjustment of tumour mutation burden by age of diagnosis, sex and stage

3.2 Relative signature attributions associations with incidence rates

3.3 Association of total mutational burden with ASR excluding countries with predominant AA signatures

3.4 Estimating FDR of ASR associations with signatures

3.5 Limiting regression analysis to early-stage cancers

**4. Copy number (CN) and structural variant (SV) signatures**

4.1 Extraction of CN and SV signatures

4.2 Associations of CN and SV signatures

**5. Germline Analysis**

5.1 Polygenic risk scores (PRS) across countries

5.2 SBS12 and common germline variants

**6. Untargeted metabolomics analysis – extended methods and quality control**

6.1 Sample preparation

6.2 Sample analysis

6.3 Data processing

6.4 Quality control

**7. Subclonal mutational signatures**

7.1 Validation of the detection of subclonal mutational signatures

#### 1. Mutational signature analysis

##### 1.1 Extraction of *de novo* mutational signatures with SigProfilerExtractor

Extractions were performed for single base substitutions (SBSs), doublet base substitutions (DBSs), and small insertions and deletions (IDs; **Supplementary Figs 1-3**). SBS extractions were performed using both SBS-288 and SBS-1536 contexts. These two contexts extend the SBS-96 classification in two independent ways. SBS-288 by considering the SBS-96 contexts on transcribed and untranscribed strands of protein coding genes as well as by including mutations on intergenic non-transcribed DNA. SBS-1536 is formed of a pentanucleotide context formed of the two flanking bases on both the 5' and 3' of the mutated base. Although using different information to extract mutational signatures, the two extractions were largely concordant (**Supplementary Note Table 1**) with two observed differences:

1) SBS-1536 was able to extract an additional signature (SBS1536C/SBS_C, **Supplementary Fig. 1**). Visual inspection of this signature showed a similarity to SBS1536H/SBS_H, a signature which was driven by a single hypermutator case (**Supplementary Fig. 4**). On this basis, we hypothesised that these two signatures were more distinct at the SBS-1536 level than at the SBS-288 level which would lead to a failure to extract them separately in the SBS-288 extraction. To test this, we performed an SBS-288 extraction with the hypermutator case removed, which was able to extract a signature corresponding to SBS1536C/SBS_C, thus supporting its existence.

2) SBS-1536 extracted an additional 'flat' signature (SBS1536E/SBS_E), in addition to three extracted in both SBS-288 and SBS-1536 format (**Supplementary Fig. 1**). Flat signatures are termed so because they lack distinct peaks in specific contexts but rather have variable peaks spread across all substitution types. The consequence of this is that they are very difficult to accurately distinguish between, and again, the

difference between the two extractions is very likely to be due to the signature being more distinct at the SBS-1536 level compared to the SBS-288.

For our dataset, the SBS-1536 format was selected for further analysis due to its ability to extract additional signatures. In principle, whether SBS-288 or SBS-1536 is more effective is likely to vary between datasets, depending on which signatures are present and the amount of overlap between substitution types.

##### 1.2 Extraction of *de novo* signatures with mSigHdp

To ensure that the mutational signature landscape was fully reflected in the SigProfilerExtractor results, extraction of mutational signatures was also performed with mSigHdp. In contrast to SigProfilerExtraction, which utilises nonnegative matrix factorization, mSigHdp leverages a hierarchical Dirichlet process.[1] However, unlike SigProfilerExtractor, mSigHdp has only been benchmarked for use with SBS-96 and ID-83 contexts which prevents a comprehensive direct comparison. mSigHdp extracted 11 SBS signatures and 6 ID signatures (**Supplementary Figs 5-6**). The 11 SBS signatures were largely concordant with the SBS-288 signatures (**Supplementary Note Table 1**), and the same differences compared to SBS-1536 described above were observed. The exception to this was the second Aristolochic acid signature hdp11, which only had a cosine similarity of 0.75 and 0.77 to SBS1536I (SBS_I) and SBS288J respectively. Despite the differences in the signature extracted, the mSigHdp results support the existence of a second SBS Aristolochic acid signature. Given that this second Aristolochic acid signature has not been identified in prior studies which have been largely focused on SBS-96 contexts, it is likely that the extended contexts are important for distinguishing between the two signatures. mSigHdp extracted 6 ID-83 signatures, one less than SigProfilerExtractor. Overall, the results were similar, with mSigHdp extracting ID_A and ID_B as a single signature (hdp2) explaining the difference in the number of signatures extracted.

##### 1.3 Decomposition to reference signatures

Decomposition of SBS *de novo* mutational signatures to the Catalogue of Somatic Mutations in Cancer (COSMIC) reference signatures was performed with SigProfilerAssignment (https://github.com/AlexandrovLab/SigProfilerAssignment) Currently, the method for distinguishing between a novel mutational signature and those composed of multiple COSMIC reference signatures has limitations. Firstly, as the reference signatures are currently only available in SBS96 contexts, de novo signatures in extended context must be collapsed to SBS96 format. This leads to the loss of information which may have assisted in distinguishing novel signatures from the COSMIC reference signatures. Secondly, the default decomposition settings use the entire COSMIC reference set even if it is not plausible for some signatures to be present in a given tissue type. Lastly, the large number of SBS-96 signatures in the current COSMIC reference set means that it is increasingly difficult to declare a mutational signature novel, as most decompositions can achieve a cosine similarity of 0.80 using combinations of the existing reference signatures. This is a particular problem for flat signatures which are difficult to distinguish between due to the lack of unique features at the SBS96 context level. Both previous studies and ours have shown that the flat signatures SBS5 and SBS40 are present in clear cell renal cell carcinomas (ccRCC).[2,3]

SigProfilerAssignment has two parameters that allow the optimisation of the decomposition process. The first (exclude_signature_subgroups) allows the exclusion of sets of signatures which share a common aetiology from the pool of signatures used in the decomposition. Justifications for excluding subgroups may include reviewing the mutational spectra and germline /somatic mutation calls to exclude signatures associated with DNA repair deficiencies, review of clinical data to exclude the possibility of prior chemotherapy and other treatments/exposures and lastly removing subgroups unlikely to be present in the tissue type based on biological mechanisms and/or previous literature. The second parameter which can be altered is newsignature_threshold, which determines the cosine similarity threshold at which a signature is considered novel. The default value for this is 0.8, meaning that if a signature can be reconstructed using COSMIC reference signatures with a cosine similarity to the *de novo* signature of 0.8 or more, then the signature will be decomposed. Adjusting this parameter in order to define a signature as novel should only be used when there is strong evidence to reject the default decomposition. Examples of such justifications may include; association of the with metadata/risk factors or in exposure studies, where the signature is found only in the treated subset, the presence of individual mutational spectra which support the existence of the signature, extraction of the mutational signature using multiple signature extraction methodologies, the presence of specific context peaks which are not explained by the decomposition and/or the presence of incompatible features such as transcriptional strand bias.

Using default settings all the SBS *de novo* signatures could be decomposed using the default parameters. The final decompositions were performed using the SBS-1536 *de novo* signatures with custom parameters. For SBS signatures we increased the newsignature_threshold from 0.80 to 0.95. Secondly, the following groups of signatures were excluded; POL deficiency signatures (SBS10a, SBS10b, SBS10c, SBS10d, SBS28), homologous recombination (HR) deficiency signatures (SBS3), base excision repair (BER) deficiency signatures (SBS30, SBS36), iatrogenic signatures (SBS11, SBS25, SBS31, SBS32, SBS35, SBS86, SBS87, SBS90), ultraviolet (UV) signatures (SBS7a, SBS7b, SBS7c, SBS7d, SBS38), lymphoid signatures (SBS9, SBS85, SBS86), and artefact signatures (SBS27, SBS43, SBS45, SBS46, SBS47, SBS48, SBS49 , SBS50, SBS51, SBS52, SBS53, SBS54, SBS55, SBS56, SBS57, SBS58, SBS59, SBS60). These parameters result in five signatures (SBS_A, SBS_B, SBS_F, SBS_H, and SBS_I) remaining non-decomposed (**Fig.2, Supplementary Table 8, Supplementary Note Table 2**). For ID signatures, the decision was made to increase the threshold for a novel signature to 0.90, which results in a single signature (ID_C) remaining non-decomposed (**Extended Data Fig.5, Supplementary Table 8**). No alternation was deemed necessary for the decomposition of DBS78 *de novo* signatures, with one signature (DBS_D) remaining non-decomposed on default parameters (**Extended Data Fig.4, Supplementary Table 8**).

**1.4 Justification for non-decomposed signatures**

Although deviating from the default parameters during decomposition is an arbitrary decision, we consider this justified in the light of the additional evidence supporting the fact that these signatures genuinely represent distinct mutational processes.

Due to the nature of the flat signatures, including COSMIC reference signatures SBS5 and SBS40, it has been speculated that they do not represent a singular mutagenic process but instead multiple processes which are extremely difficult to separate due to their high level of correlation. Our results extracted 4 flat signatures. One of these, SBS_E is decomposed whereas SBS_A, SBS_B, and SBS_F remained non-decomposed in the optimised decomposition. The original decompositions for SBS_A included SBS1, SBS5, SBS7c, SBS28 and SBS40 (**Supplementary Note Table 2**). SBS7c is associated with UV light exposure. This is unlikely given that there is no plausible biological mechanism that could explain the presence of this signature in this cancer type. SBS28 is also not likely, as this signature is usually found in samples with signatures SBS10a/10b which are associated with POLE mutations. For SBS_B the original decomposition used SBS30, which is associated with a deficiency in base excision repair due to NTHL1 mutations. Again, this is unlikely due to the high number of cases positive for the signature and the lack of detectable NTHL1 mutations. Lastly SBS_F was decomposed to SBS5, however this was considered unlikely considering the differences in the T>C peaks and the transcription bias observed in the SBS288 counterpart was not consistent with that observed in SBS5.[4] Given that we did not extract a single mutational signature that directly matched SBS40 in any of the three extractions performed, whereas the combination of SBS_A, SBS_B, and SBS_F matches the COSMIC SBS40 with a cosine similarity of 0.96, we have provisionally named these signatures SBS40a, SBS40b, and SBS40c. (**Supplementary Fig. 7**). The finding that SBS40b (and to a lesser extent SBS40a) is associated with incidence, whereas SBS40c is not, in addition to the unique association of SBS40b with N, N, N-trimethyl-L-alanyl-L-proline betaine (TMAP) imply that these are distinct mutational processes which have biologically meaningful implications in ccRCC. There are multiple factors which contribute to the ability to extract a mutational signature which could explain why SBS40 was extracted as a single signature in previous analysis. These include the use of extended contexts, cohort size, variation between samples and the continual improvement of the methodology. However, supplementary analysis from the PCAWG study from which SBS40 was originally extracted show that signatures similar to SBS40a/b/c can be extracted from the ccRCC cohort when analysed individually (**Supplementary Fig.8**).[3] This observation suggests that some signatures are more difficult to extracted separately in the presence of multiple cancer types, and since this analysis was performed in SBS96 format with a limited number of samples this is likely an important factor in this scenario. Further exploration of the conditions required to separate SBS40 is included in the section 1.5 of the supplementary note. While our analysis has enabled the separation of SBS40 into three components, it is important to note that the SBS40a/b/c components may still represent multiple mutational process which potentially could split further in future studies.

The original decomposition for SBS_I included SBS27 and SBS90, the former of which is an artefact signature while the latter is associated with chemotherapy, however, this is extremely unlikely that so many individuals from a specific country would show evidence of artefacts and chemotherapy associated signatures. There is strong evidence to consider SBS_I (in addition to DBS_D and ID_C) as novel signatures associated with Aristolochic exposure. SBS_I was extracted in both SigProfilerExtractor 288 and 1536 formats, and while the mSigHdp version of the signature was not an exact match it did nonetheless agree that there was a second SBS signature in addition to the COSMIC signature 22. It is also possible to identify mutational spectra from individual tumours which are dominant in each signature (**Supplementary Fig. 9**). All four signatures correlate strongly with each other, and show enrichment in the same countries (Romania, Serbia and Thailand, **Extended Data Fig.6-7**). Finally, the original decomposition for SBS_H signature included SBS8 which has an unknown aetiology and SBS32 which is associated with azathioprine treatment. Given the presence of a hypermutator whose mutational spectra is a close match to the signature (**Supplementary Fig. 4**), rejection of this decomposition is justified. The cancer of this individual has a mutational burden exceeding the mutational burdens of cancers with the strongest Aristolochic exposure; however, with only a single case, it is impossible to determine whether this is due to an environmental exposure or due to a currently unknown defect in DNA repair.

##### 1.5 Exploration of conditions required for SBS40a, SBS40b and SBS40c separation.

To further explore the conditions required to separate SBS40 into SBS40a, SBS40b and SBS40c, additional extractions were preformed using 500, 250 or 125 randomly sampled cases from the whole cohort in SBS96 and SBS288 context, each repeated 5 times for a total of 30 scenarios. The cosine similarity between the extracted signatures from each scenario and the SBS40a/b/c signatures used for final analysis was then calculated (SBS288 scenario signatures were first collapsed into SBS96 context to enable these comparisons).

The results show that signatures corresponding to SBS40a and SBS40b could be extracted from all but one scenario with an overall average cosine similarity of 0.94 and 0.97 respectively (**Supplementary Note Table 3**). The single scenario where SBS40 could not be separated was a SBS96 scenario with only 125 samples. SBS40c extraction was more variable, being extracted in the majority of the 500 samples extractions (5/5 SBS288 and 4/5 SBS96) but less frequently in the 250 (3/5 SBS288 and 1/5 SBS96) and rarely in the 125 (2/5 SBS288 and 0/5 SBS96) scenarios (**Supplementary Note Table 3**). This behaviour is expected for two reasons, firstly that SBS40c is found in fewer samples and at lower proportions compared to SBS40a and SBS40b but also considering that SBS40c is the most similar component to the flat signature SBS5 (cosine similarity 0.91 compared to 0.71 and 0.81 for SBS40a and SBS40b). Both factors make SBS40c the most difficult of the three components to extract.

The average cosine similarity in scenarios where SBS40c could be extracted was 0.95 (**Supplementary Note Table 3**).

The benefits of using extended contexts were demonstrated in two ways, firstly SBS40c was extracted more frequently in SBS288 scenarios, as the extended context information enhances the ability to distinguish between mutational signatures which are highly similar in SBS96 context. Secondly, the average cosine similarity across all scenarios was slightly higher for SBS40b and SBS40c in SBS288 context compared to SBS96 context (0.96 vs 0.98 and 0.93 vs 0.97 respectively), but equal for SBS40a (both 0.94, **Supplementary Note Table 3**). In terms of cohort size, for all three signatures the average cosine similarity improved with increasing cohort size with the closest match to the original SBS40a/b/c obtained in the 500 sample runs for both SBS96 and SBS88 contexts **(Supplementary Note Table 3)**. It is worth noting however, that for SBS40b the average cosine similarity was the same in the 250 and 500 samples scenarios whereas for SBS40a and SBS40c the 500 sample scenarios had increased average cosine similarity **(Supplementary Note Table 3)**, which reflects the fact that the exact number of samples required to extract any mutational signature reproducibly will be variable. Overall, the set of scenarios which recreated the original SBS40a/b/c with the highest average cosine similarity was the SBS288 500 sample set (0.98, 0.99 and 0.99 respectively). SBS40c was extracted from all scenarios in this set and for all three signatures cosine similarity was consistently high (0.97- 0.99) between individual scenarios, which demonstrates that at this point conditions were sufficient to stably extract each signature with minimal contamination from the other components.

Overall, these results show that the splitting of SBS40 is highly reproducible. While the impact of sample numbers, context and cohort diversity are unique to each mutational signature and the cohort that they are extracted from, such investigations are useful particularly when analysing the potential splitting of an existing mutational signature. By testing the reproducibility of a split in this manner, if conditions can be identified where each component is consistently extracted with high average cosine similarity and low variability (as is the case for the SBS288 500 sample set of scenarios for SBS40 components), then a split could be considered reproducible and likely to represent distinct mutational processes. If components are extracted inconsistently within a set of scenarios and/or variability in the cosine similarity is observed, then this would instead suggest that the extraction conditions are still influencing the extraction process and thus may not be generating reproducible results.

##### 1.6 Presence of tobacco-associated signature SBS4

Whilst tobacco has been shown to be a risk factor for RCC, SBS4 has not been previously identified in RCC.[2,5] In this study SBS4 was identified as a component of the *de novo* signature SBS_C, which is decomposed into COSMIC signatures SBS4 and SBS40. To provide confidence in the presence of SBS4 the following steps were performed. Firstly, the signature was re-decomposed using a custom COSMIC reference signature list where SBS40 is replaced with SBS40a, SBS40b, and SBS40c. This shows that the *de novo* signature is composed solely of SBS4 (34.52%) and SBS40a (65.48%). Whilst it is not possible to completely remove the SBS40a component, subtracting the SBS40a contexts at the above ratio should leave a signature which is a closer match to the COSMIC reference SBS4, and indeed, the resulting adjusted signature has a cosine similarity of 0.90 compared to 0.80 in the original *de novo* signature (**Supplementary Fig. 10**). The adjusted signature notably lacks the T>A peaks present in the reference SBS4, whilst comparing just the C>A compartment increases the cosine similarity of the adjusted signature even further to 0.95. In a previous study of environmental exposures, which included many compounds found in tobacco smoke, the compounds dibenzo[*a,l*]pyrene diol-epoxide (DBPDE) and dibenzo[*a,l*]pyrene (DBP) generated a profile which strongly resembles the T>A peak observed in SBS4 (**Supplementary Fig. 10**).[6] We can speculate that the absence of these peaks in the adjusted signature indicates that not all mutagenic components of tobacco smoke are present in the kidney, however this would require additional study to confirm. For the purposes of this study, the adjusted signature provides sufficient evidence of an SBS4-like signature in ccRCC.

##### 1.7 Attribution of single base substitution signatures

For *de novo* SBS signatures, the decision was made to exclude SBS_H. This signature was driven by a single hypermutator, without which the signature is not extracted. However, when included in the attribution panel, this signature present in 208 /962 cases (22%) likely due to overlap in contexts with multiple other signatures. Therefore, SBS_H was removed from the final *de novo* attribution panel. For COSMIC reference SBS signatures, attributions were performed on the subset of COSMIC reference signatures which are present in the dataset (as determined by SigProfilerAssignment following decomposition of the *de novo* signatures), in addition to any non-decomposed signatures. Two changes were made to this for the final attributions. Specifically, SBS40 was removed from the panel of signatures. SBS40 was found in several decompositions, likely where there is a low-level background of mutations in the *de novo* signatures. However, it does not make sense to include it the final panel given the presence of SBS40a, SBS40b and SBS40c. Additionally, SBS_H was removed from the final COSMIC attribution panel for the same reasoning as for *de novo* signatures.

#### 2. Attribution of mutational signatures in external data sets

##### 2.1 Attribution of SBS40 components in a pan-cancer cohort

In order to investigate the patterns of attribution of SBS40a, SBS40b, and SBS40c, SigProfilerAssignment was used to attribute an altered COSMIC reference set where SBS40 is replaced with SBS40a, SBS40b, and SBS40c on a pan-cancer cohort dataset.[3] This showed that SBS40a was found in the majority of tumour types, whilst SBS40b and SBS40c were only seen consistently in clear cell RCC (**Extended Data Fig 3**). Notably, the chromophobe RCC dataset did not show attribution to either SBS40b or SBS40c, suggesting that these signatures are likely further restricted to certain niches within the kidney. These results provide additional support for SBS40a, SBS40b, and SBS40c representing distinct mutational processes.

##### 2.2 Attribution of SBS12 in liver cancers

The COSMIC reference signature SBS12 was originally extracted from liver cancers.[2,3] The liver cancers used in this extraction consisted of three cohorts, one of which (LINC-JP) is from Japan. In order to determine whether SBS12 enrichment is also present in Japanese liver cancers, SigProfilerAssignment was used to attribute the COSMIC v3.3 reference signatures. This shows that SBS12 is enriched in the LINC-JP cohort compared to the LICA-FR (France) and LIHC-US (US) cohorts (**Supplementary Fig. 11**).

##### 2.3 Validation of SBS12 in additional datasets

In order to validate the presence of SBS12 in Japanese ccRCC we attributed our mutational signatures in two additional cohorts. The first cohort consisted of 14 whole genome sequenced cases, for which the provided variant calls were used to generate an SBS96 matrix to which the ccRCC mutations signatures were attributed.[7] While additional cases with whole exome sequencing were available from this study, the number of mutations is too low to reliably detect the presence of SBS12 in exomes. The second cohort consisted of 61 Japanese ccRCC sequenced by the same centre which provided the primary cohort. Variant calling on this cohort was performed using the same methodology as the primary cohort. SBS12 was detected at similar levels in both validation cohorts at similar frequencies (72%, 85% and 75%, **Supplementary Fig.12a**). There were no significant differences in age of diagnosis and the overall mutational profile was similar in all three cohorts (**Supplementary Fig.12b-c**).

#### 3. Tumour Mutation Burdens and regression analysis

##### 3.1 Adjustment of tumour mutation burden by age of diagnosis, sex and stage

In order to detect potential bias due to variations in age of diagnosis, sex and stage of cases across countries, tumour mutation burden (TMB) was adjusted by these variables. This was performed by calculating residuals in TMB ~ age + sex + stage linear regression model, and adding them to the overall mean TMB across all countries in order to get the corrected TMB. The resulting values are independent of age, sex and stage distribution for each country. Since stage information was missing for some cases, for example 5 cases from Thailand, these cases were only corrected by age and sex. Nevertheless, all these adjustments had minimal impact on the plots and did not alter the messages conveyed in the study, as shown in the **Supplementary Note Fig. 13** illustrating the differences between adjusted and unadjusted TMB. Therefore, uncorrected TMB plots are shown in the main text, to be consistent with the literature[8], while the adjusted plots are shown here for reference.

##### 3.2 Relative signature attributions associations with incidence rates

The associations of incidence rates with relative signature attributions (i.e., the proportion of mutations assigned to signatures with respect to tumour mutation burden) are largely similar to those with absolute mutation counts, particularly for significant associations with signatures SBS40b and ID5 (**Supplementary Fig. 14 and Supplementary Note Table 4**).

##### 3.3 Association of total mutational burden with ASR excluding countries with predominant AA signatures

Although the association if total mutation burden with ASR was not significant when all countries were present, it did appear significant when countries with predominant AA signatures (Romania, Thailand and Serbia) were excluded. These results are summarised in **Supplementary Note Table 5 and Supplementary Fig. 15**.

##### 3.4 Estimating FDR of ASR associations with signatures

To estimate false discovery rate (FDR) of associations of ASR incidence rates with mutational signatures, two approaches were considered. In the first one, randomly generated signatures were added to the attribution process. Poisson processes were used to generate the signatures for SBS and ID mutation types (**Supplementary Fig. 16**), for which significant associations were found (signatures SBS40b, ID5 and ID8). During the attribution process performed with the MSA tool that uses parametric bootstrap to derive confidence intervals of attributions, only 2.3% of cases were attributed with the randomly generated SBS signature, and 0.1% for the randomly generated ID signature. No associations with incidence rates were found for such sparse signatures.

In an alternative approach, signature attributions were permuted, and regressed with incidence rates. The process was performed 1000 times. The distribution of p-values was uniform for all permuted signatures and mutation burdens reported as significant (**Supplementary Fig. 17**). False positive rate (FPR) was consistent with the conservative p-value threshold of 0.05 divided by the number of tests (signatures), yielding the FDR ≤ 0.05.

##### 3.5 Limiting regression analysis to early-stage cancers

To elucidate whether mutational signature associations with epidemiological data become more apparent in early-stage tumours, regression analysis was performed on stage I and stage II tumours only (495 cases). The associations remained largely similar to the analysis with the full cohort, albeit with lower significance due to lower sample size. The regression results are summarised in **Supplementary Note Table 6.**

#### 4. Copy number (CN) and structural variant (SV) signatures

##### 4.1 Extraction of CN and SV signatures

Copy number (CN) and structural variant (SV) signatures were extracted and assigned using the same methods and tools as with SBS, DBS and ID signatures, i.e. SigProfiler and MSA. Copy number signatures were generated using the 48-channel copy number classification scheme, as per COSMIC reference set. SV signatures were extracted and assigned *de novo* based on a previously developed classification in breast cancer[45,46]. The reference set of SV signatures was not available in COSMIC at the time of publication, hence only *de novo* signatures were analysed. The signatures are summarised in **Supplementary Fig 18-19.**

##### 4.2 Associations of CN and SV signatures

No significant associations were found with recorded risk factors, as shown in **Supplementary Note Table 7**, apart from the association of signature CN1 with sex. Of note, signatures CN1 and SV_B were associated with stage.

Associations of CN and SV signatures with ASR are summarised in **Supplementary Note Table 8**. Two significant associations were observed, namely, for signature CN9 which is a signature of chromosomal instability on a diploid background, as well as signature SV_A, which mainly comprises non-clustered translocations. CNV burden was also negatively associated with ASR, with low-incidence countries having a higher CNV burden on average. These associations are also illustrated in **Supplementary Fig 20**.

#### 5. Germline Analysis

##### 5.1 Polygenic risk scores (PRS) across countries

Polygenic risk scores for instruments shown in **Supplementary Table 18** did not vary meaningfully across countries (**Supplementary Fig. 21**) and did not associate with incidence rates of ccRCC (**Supplementary Note Table 9**).

##### 5.2 SBS12 and common germline variants

Given the ubiquity of signature SBS12 in Japan, as well as a well-known prevalence of a germline variant in ALDH2, rs671 (G>A) polymorphism in Japanese population, it is reasonable to question whether there is any association between SBS12 and rs671. A logistic regression analysis adjusted by age, sex, alcohol ever-drinking status and the first five principal components of genetic ancestry was performed, yielding no significant association of the SBS12 signature attribution with the rs671 germline variant (p=0.81). The null finding was also replicated using the validation cohort of additional 61 ccRCC cases from Japan (p=0.28). Additionally, an exploratory analysis was performed to evaluate whether any common germline genetic variants (MAF>1%) were associated with SBS12 within the

Japanese cohort. Similar logistic regressions revealed no statistically significant associations between common genetic variants and SBS12, neither at genome-wide significant level nor passing multiple-testing correction. As such, at this point there is little clear evidence for the influence of germline variants on SBS12.

#### 6. Untargeted metabolomics analysis – extended methods and quality control

##### 6.1 Sample preparation

Samples (901) were thawed at room temperature and prepared by mixing 30 µL of plasma with 200 µL of cold acetonitrile and filtering the precipitate with 0.2 µm Captiva ND plates (Agilent Technologies). A 100-µL aliquot of the filtrate was then mixed with equal volume of ultrapure water in a well plate that was then sealed (Rapid EPS, BioChromato, Fujisawa, Japan), frozen at -80 C, and thawed at room temperature before analysis. Quality control (QC) samples were prepared from a plasma pool that was made by mixing small aliquots of 100 randomly selected study samples and processed together with the study samples. Blank samples were also prepared along the plasma samples in an identical manner, only leaving the plasma out of the process. Each well plate included four individually prepared QCs and two blanks.

##### 6.2 Sample analysis

Samples were analyzed as two independent analytical batches both consisting of five 96-well plates. Samples were randomized across the batches. Analysis was performed with a UHPLC-QTOF-MS system that consisted of a 1290 Binary LC and a 6550 QTOF mass spectrometer equipped with Jet Stream electrospray ionization source (Agilent Technologies). Samples were kept at 4°C and 2 µL was injected. An ACQUITY UHPLC HSS T3 column (2.1 × 100mm, 1.8 µm) was maintained at 45 °C and the mobile phase consisted of ultrapure water and LC-MS grade methanol, both with 0.05 % (v/v) of formic acid. Following gradient profile was used: 0–6 min: 5% to 100% methanol, 6–10.5 min: 100% methanol, 10.5–13 min: 5% methanol. Mobile phase flow rate was 0.4 ml/min.

Mass spectrometer drying gas temperature was 175°C and flow 12 L/min, with capillary, nozzle, and fragmentor voltages of 3500 V, 300 V, and 175 V, respectively. The sheath gas temperature was 350°C and flow 11 L/min, and nebulizer pressure 45 psi. Continuous mass axis calibration was employed using lock mass ions *m/z* 121.0509 and 922.0098. Data was acquired in centroid format using an extended dynamic range mode, and acquisition rate of 1.67 Hz, over the mass range of 50-1000 Da (MassHunter Acquisition 10.1, Agilent Technologies).

**6.3 Data processing**

Pre-processing was performed using Profinder 10.0.2.162 and Mass Profiler Professional B.14.9.1 software (Agilent Technologies). A “Batch recursive feature extraction (small molecules)” process was employed for samples and blanks to find $[M+H]^+$ ions. Height thresholds of 2000 and 8000 counts for mass and chromatographic peaks were used, respectively, and a minimum quality score of 70. Feature alignment between samples was performed with retention time and mass windows of ±0.04 min and ±(10 ppm + 1 mDa), respectively. A target list for the recursive extraction was created by including features found in at least 12 of the samples. For recursive feature extraction ±35 ppm width was used for the *m/z* values to draw chromatographic peaks, with Agile 2 integrator and no smoothing, and the mass calculated as an average from spectra >60% peak heights. No filtering was applied. The two batches were processes separately and the resulting features were aligned in Mass Profiler Professional. Features present in every blank sample were excluded, unless 5-fold greater in average intensity in samples, for both batches separately. Remaining features present in at least one study sample were used as targets in a “Batch targeted feature extraction” process for study samples and QC samples in Profinder. The two batches were processed independently and all resulting output (.cef) files were aligned in Mass Profiler Professional and exported as a single .csv file. Chromatographic peak areas were used as a measurement of intensity. No normalization or transformation of raw data was performed prior to the downstream data analysis.

##### 6.4 Quality control

Quality control was performed using data from QC samples.

The assessment was based on following attributes:

- Response stability: in chronological order, log2-normalized average response of features found in all QC samples
- Response variability: distribution of relative standard deviations (RSD%) of features found in all QC samples
- Response variability: RSD% of 10 known compounds in all QC samples

The analytical run was initiated with priming injections of a QC sample to achieve stable instrument response, followed by a blank sample and the study samples, with one QC sample after every 10-12 injections. Quality assessment of the raw data before any transformation or adjustment was based on monitoring of known target metabolites in the QC samples. Intra-batch RSD% of ten representative metabolites listed **in Supplementary Note Table 10** ranged from 8.3% to 15.9% (N=79), with retention times within 0.03 min across the data. The QC results are illustrated in **Supplementary Note Fig. 22 and 23**.

#### 7. Subclonal mutational signatures

##### 7.1 Validation of the detection of subclonal mutational signatures

In order to quantify the power to detect subclonal mutations and, subsequently, subclonal mutational signatures, we calculated the number of reads per tumor chromosomal copy (NRPCC), as described in a recent report characterizing the intra-tumor heterogeneity and subclonal architecture in the Pan-Cancer Analysis of Whole Genomes (PCAWG) cohort[9], as this is not directly dependent on the number of detected SNVs (**Supplementary Fig. 24**).

This metric considers the sample purity, tumor ploidy, and sequencing coverage, in order to quantify the power to detect subclonal clusters of mutations uniformly. We calculated the NRPCC for the whole cohort with copy number calling available (complete pipeline, n=857), as well as for the set of 223 samples considered for the evolutionary analysis after applying the filters previously described in the Methods section (at least 40% purity, at least one subclone, at least 256 clonal and subclonal mutations, no samples with poor separation between clonal and subclonal clusters, and no clusters centered at a CCF>1.5 where chromosome X contributed the highest number of mutations).

The vast majority of the filtered samples (218, 98%) had a NRPCC above 10 (**Supplementary Fig. 25**), which was the power threshold used in the PCAWG study, and that was sufficient to detect subclonal mutation clusters at CCFs from 20 to 80% (**Supplementary Note Fig. 26**). Thus, we validated that our filtering strategy was consistent with previous studies and allowed us to accurately identify subclonal mutational signatures in the sufficiently powered samples.

#### SUPPLEMENTARY REFERENCES

1. Liu, M., Wu, Y., Jiang, N., Boot, A. & Rozen, S. G. mSigHdp: hierarchical Dirichlet process mixture modeling for mutational signature discovery. *NAR Genom Bioinform* **5**, (2023).
2. Alexandrov, L. B. *et al.* The repertoire of mutational signatures in human cancer. *Nature* **578**, 94–101 (2020).
3. Islam, S. M. A. *et al.* Uncovering novel mutational signatures by de novo extraction with SigProfilerExtractor. *Cell Genomics* **2**, 100179 (2022).
4. Otlu, B. *et al.* Topography of mutational signatures in human cancer. *Cell Rep* **42**, 112930 (2023).
5. Alexandrov, L. B. *et al.* Mutational signatures associated with tobacco smoking in human cancer. *Science (1979)* **354**, 618–622 (2016).
6. Kucab, J. E. *et al.* A Compendium of Mutational Signatures of Environmental Agents. *Cell* **177**, 821-836.e16 (2019).

7. Sato, Y. *et al.* Integrated molecular analysis of clear-cell renal cell carcinoma. *Nat Genet* **45**, 860–867 (2013).
8. Moody, S. *et al.* Mutational signatures in esophageal squamous cell carcinoma from eight countries with varying incidence. *Nat Genet* **53**, 1553–1563 (2021).
9. Dentro, S. C. *et al.* Characterizing genetic intra-tumor heterogeneity across 2,658 human cancer genomes. *Cell* **184**, 2239-2254.e39 (2021).

#### SUPPLEMENTARY FIGURE LEGENDS

**Supplementary Note Table 1: Comparison of single base substitution signatures extracted by SigProfilerExtractor and mSigHdp.**

**Supplementary Note Table 2: Rejected SBS signature decompositions**

**Supplementary Note Table 3: Exploration of conditions required for SBS40a, SBS40b and SBS40c separation.**

**Supplementary Note Table 4. Associations of relative attribution of COSMIC mutational signatures with age-standardised incidence rates (ASR) of ccRCC**

**Supplementary Note Table 5. Associations of attribution of COSMIC mutational signatures and mutational burdens with age-standardised incidence rates (ASR) of ccRCC excluding countries with predominant AA signatures**

**Supplementary Note Table 6. Associations of ccRCC risk factors with COSMIC mutational signatures and mutational burden for early-stage cases only**

**Supplementary Note Table 7. Associations of ccRCC risk factors with copy number (CN) and structural variant (SV) mutational signatures and mutational burden**

**Supplementary Note Table 8. Associations of copy number (CN) and structural variant (SV) mutational signatures and mutational burden with age-standardised incidence rates (ASR) of ccRCC**

**Supplementary Note Table 9. Associations of polygenic risk scores with age-standardised incidence rates (ASR) of ccRCC**

**Supplementary Note Table 10. Intensity variability of 10 known compounds across all QC samples**

**Supplementary Fig.1: Single base substitution signatures extracted by SigProfilerExtractor.**
All single base substitution (SBS) *de novo* signatures extracted in SBS-288 (11 signatures) and SBS-1536 (13 signatures) format, shown side by side for comparison. Equivalent signatures where not extracted in SBS-288 format for SBS1536C and SBS1536E. For clarity, the signatures context is retained in the signature names in this figure.

**Supplementary Fig.2: Doublet base substitution signatures extracted by SigProfilerExtractor.**
Four doublet base substitution (DBS) *de novo* signatures extracted by SigProfilerExtractor.

**Supplementary Fig.3: Small insertion and deletion signatures extracted by SigProfilerExtractor.**
Seven small insertion and deletion (ID) *de novo* signatures extracted by SigProfilerExtractor

**Supplementary Fig.4: Single base substitution mutational signature driven by a hypermutated kidney cancer.**
**(a)** A single base substitution signature extracted in SBS-1536 format (SBS_H) and **(b)** the mutational spectra of a clear cell renal cell carcinomas (ccRCC) patient which corresponds to the extracted signature. The mutation burden in this patient was the highest observed in the cohort.

**Supplementary Fig.5: Single base substitution mutational signatures extracted by mSigHdp.**
Eleven single bases substitution (SBS) *de novo* signatures extracted by mSigHdp.

**Supplementary Fig.6: Small insertion and deletion mutational signatures extracted by mSigHdp.**
Six small insertion and deletion (ID) *de novo* signatures extracted by mSigHdp.

**Supplementary Fig.7: Reconstruction of COSMIC reference signature SBS40.**
The combination (SBS_ABF) of *de novo* signatures SBS_A, SBS_B and SBS_C at equal ratios **(a)** can reconstruct the profile of COSMIC signature SBS40 **(b)** with a cosine similarity of 0.96.

**Supplementary Fig.8: Comparison of signatures previously extracted from ccRCC.**
Supplementary analysis of kidney cancers from the original study reporting SBS40 shows that mutational signatures were extracted (**a**) which are similar to those reported in this study (**b**).

**Supplementary Fig.9: Aristolochic acid mutational signatures in kidney cancers.**
Examples of individual RCC mutational spectra which support the existence of both SBS22a **(a)** and SBS22b **(b)**.

**Supplementary Fig.10: Presence of tobacco-associated signature SBS4 in in kidney cancers.**
SBS4 was identified as a component of SBS_C **(a)** which also contains SBS40a. Subtracting the SBS40a component results in SBS_C_adjusted **(b)** which has a higher overall cosine similarity (CS) to COSMIC reference signature SBS4 **(c)**. The previously determined mutational signatures of the compounds dibenzo[*a,l*]pyrene (DBP) **(d)** and dibenzo[*a,l*]pyrene diol-epoxide (DBPDE) **(e)** generate peaks which correspond to the T>A peaks observed in SBS4, and the absence of these compounds in kidney may explain the remaining difference of SBS_C_adjusted compared to SBS4.

**Supplementary Fig.11: Attribution of signature SBS12 in liver cancers**
Attribution of SBS12 in liver cancers, showing enrichment of COMSIC reference signature SBS12 in the LIRI-JP cohort (Japan) compared to those in LIRI-US (USA) and LIRI-FR (France) cohorts.

**Supplementary Fig.12: Attribution of signature SBS12 in validation cohorts**
Attribution of SBS12 in two validation cohorts, showing no significant difference in either the attribution of SBS12 (**a**) or the age of diagnosis by sex (**b**) between the main and validation cohorts. The overall mutational signatures landscape was similar between all cohorts (**c**).

**Supplementary Fig.13**: **Difference between unadjusted and adjusted tumour mutation burdens across countries**

Unadjusted (left), adjusted (middle) and the difference between unadjusted and adjusted (right) tumour mutation burdens across countries for SBS (top), DBS (middle) and ID (bottom) mutation types.

**Supplementary Fig.14: Associations of relative attributions of mutational signatures with incidence of kidney cancer**

**Supplementary Fig.15: Associations of total mutation with incidence of kidney cancer excluding countries with predominant AA signatures.**

**Supplementary Fig.16: Random signature profiles for SBS and ID mutation types, generated using Poisson distribution**

**Supplementary Fig.17: Regression of permuted signature attributions and burdens with ASR**
**(a)** Log10-transformed p-values for randomized signature attributions bootstrapped 10000 times. Box-and-whisker plots are in the style of Tukey. The line within the box is plotted at the median, while upper and lower ends indicate 25th and 75th percentiles. Whiskers show 1.5 × interquartile range (IQR), and values outside it are shown as individual data points. The red dashed line represents the Bonferroni threshold used to determine significance. **(b-h)** Quantile-quantile diagrams for signatures and burdens of interest, comparing the distribution of p-values from permutation tests with the uniform distribution.

**Supplementary Fig.18: Copy number signatures extracted by SigProfilerExtractor**

**Supplementary Fig.19: Structural rearrangement signatures extracted by SigProfilerExtractor**

**Supplementary Fig.20: Associations of copy number and structural variant signatures with incidence of kidney cancer**

**Supplementary Fig. 21: Polygenic risk scores**
Polygenic risk scores for RCC risk, BMI, diastolic and systolic blood pressure, fasting glucose and insulin as well as tobacco smoking initiation across countries.

**Supplementary Fig 22: Log2-normalized intensity distribution in QC samples**
Log2-normalized intensity distribution plots for features found in all 79 QC samples. Boxplots: median with 25th and 75th percentiles. Whiskers at 1.5 IQR, outliers in red. Samples in chronological order from left to right, batch numbers shown below the plots.

**Supplementary Fig.23: Intensity variability in QC samples**
Intensity variability for features found in all 79 QC samples

**Supplementary Fig.24: Number of reads per tumor chromosomal copy (NRPCC) with respect to the total number of SNVs** Number of reads per tumor chromosomal copy (NRPCC) with respect to the total number of SNVs, without and with filtering applied.

**Supplementary Fig.25: Number of reads per tumor chromosomal copy by filtering status**

**Supplementary Fig.26: Number of reads per tumor chromosomal copy with respect to the minimum CCF of detected clusters**
Number of reads per tumor chromosomal copy with respect to the minimum CCF of detected clusters, with and without filtering applied.

### Supplementary Fig.1: Single base substitution mutational signatures extracted by SigProfilerExtractor

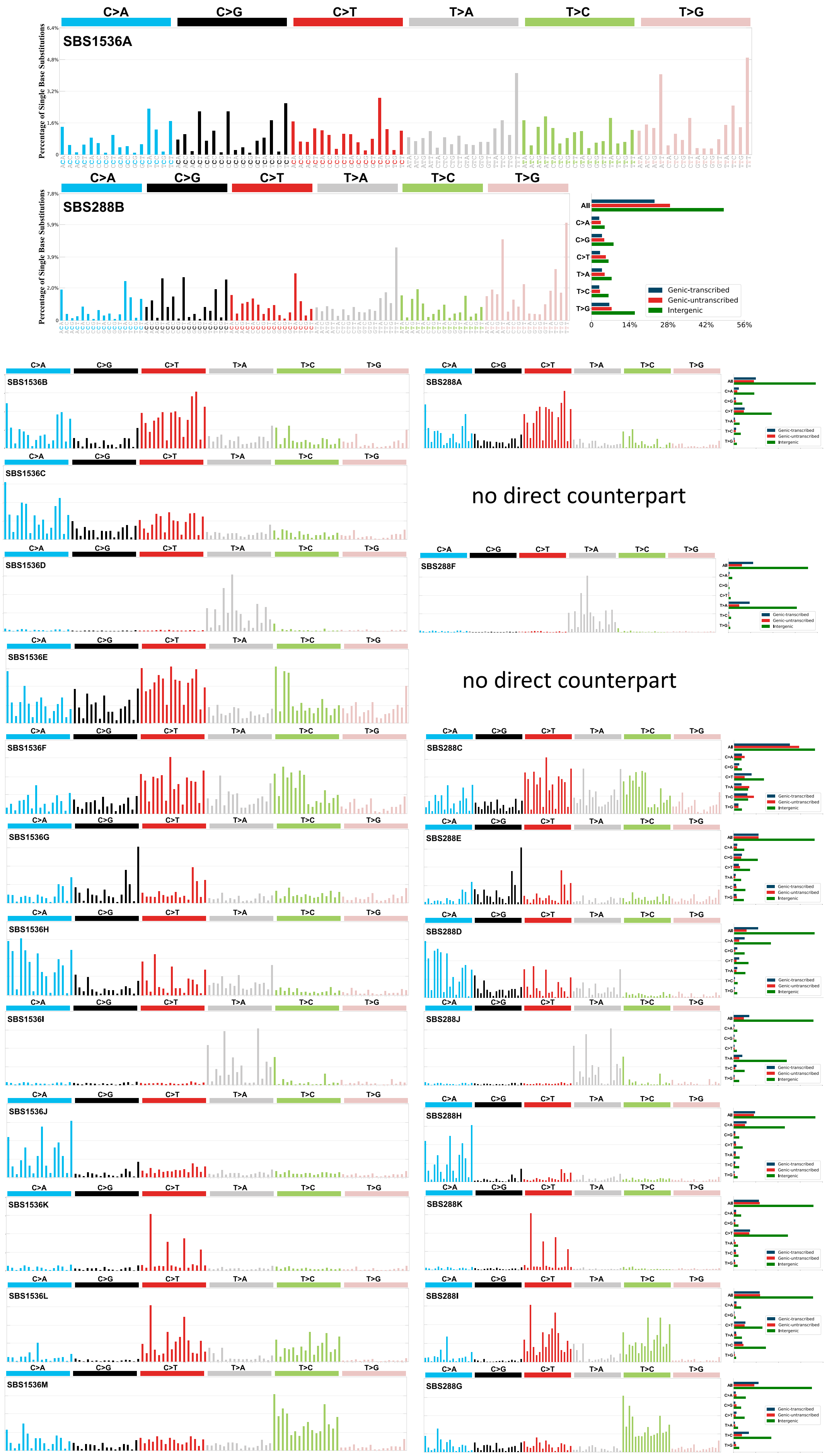

**Supplementary Fig.2: Doublet base substitution mutational signatures extracted by SigProfilerExtractor**

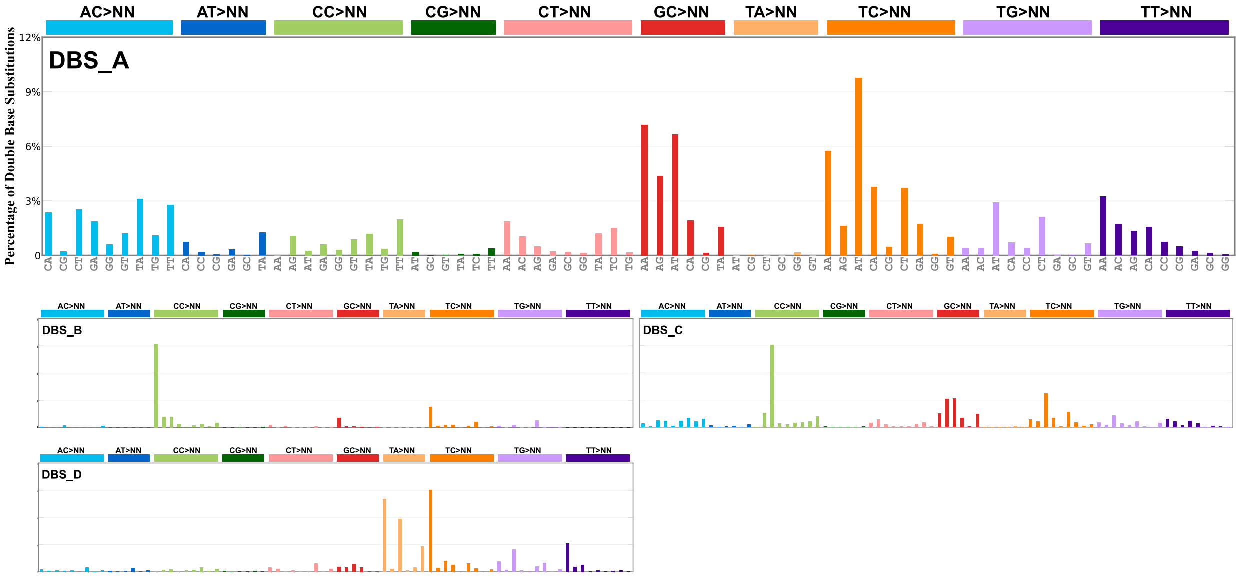

### Supplementary Fig.3: Small insertion and deletion mutational signatures extracted by SigProfilerExtractor

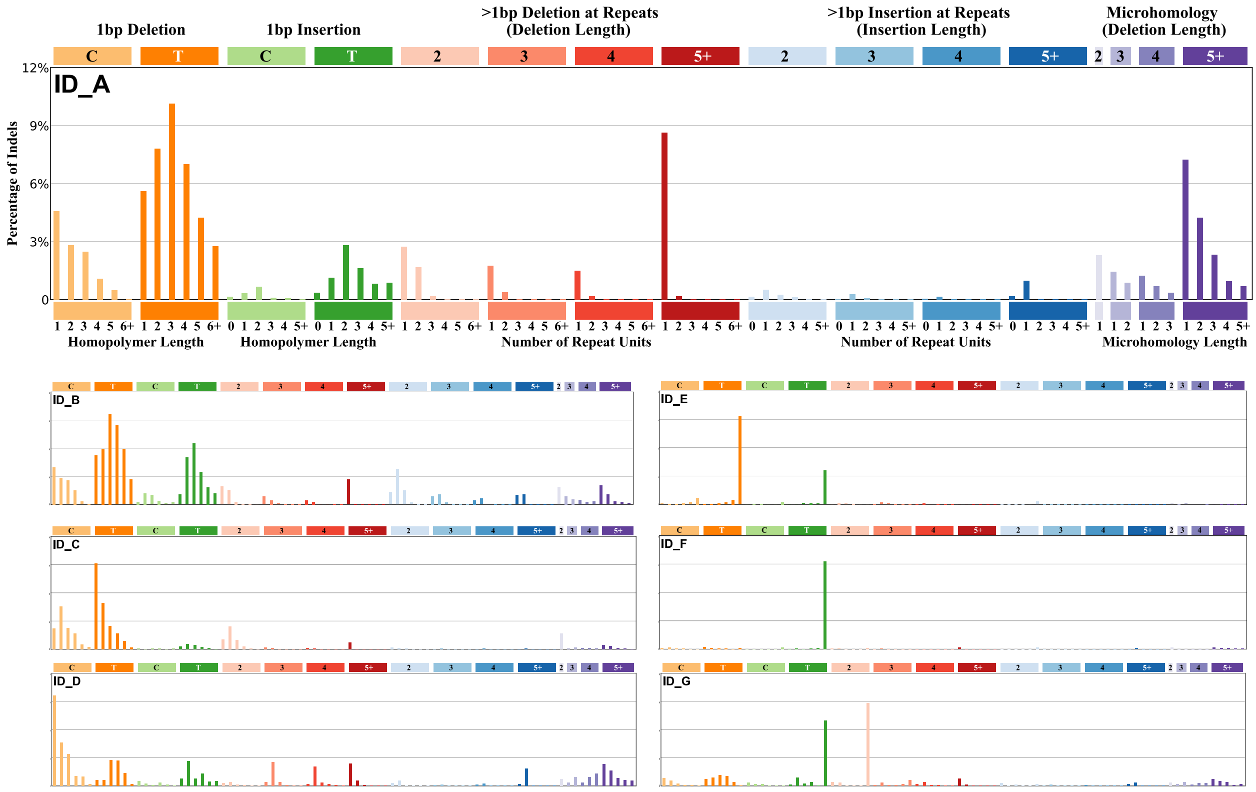

### Supplementary Fig.4: Single base substitution mutational signature driven by a hypermutated kidney cancer

A

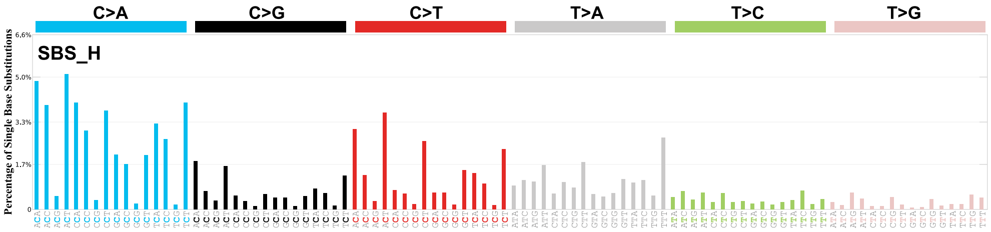


B

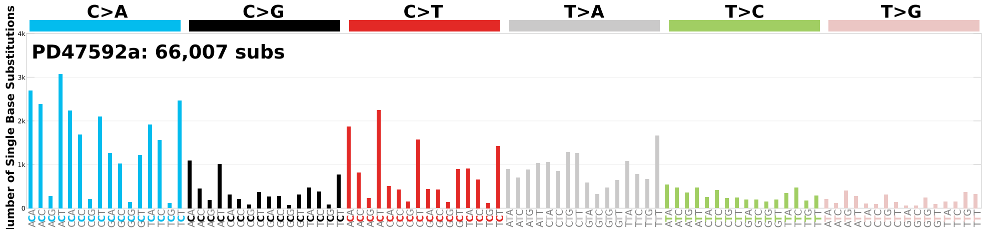

#### Supplementary Fig.5: Single base substitution mutational signatures extracted by mSigHdp

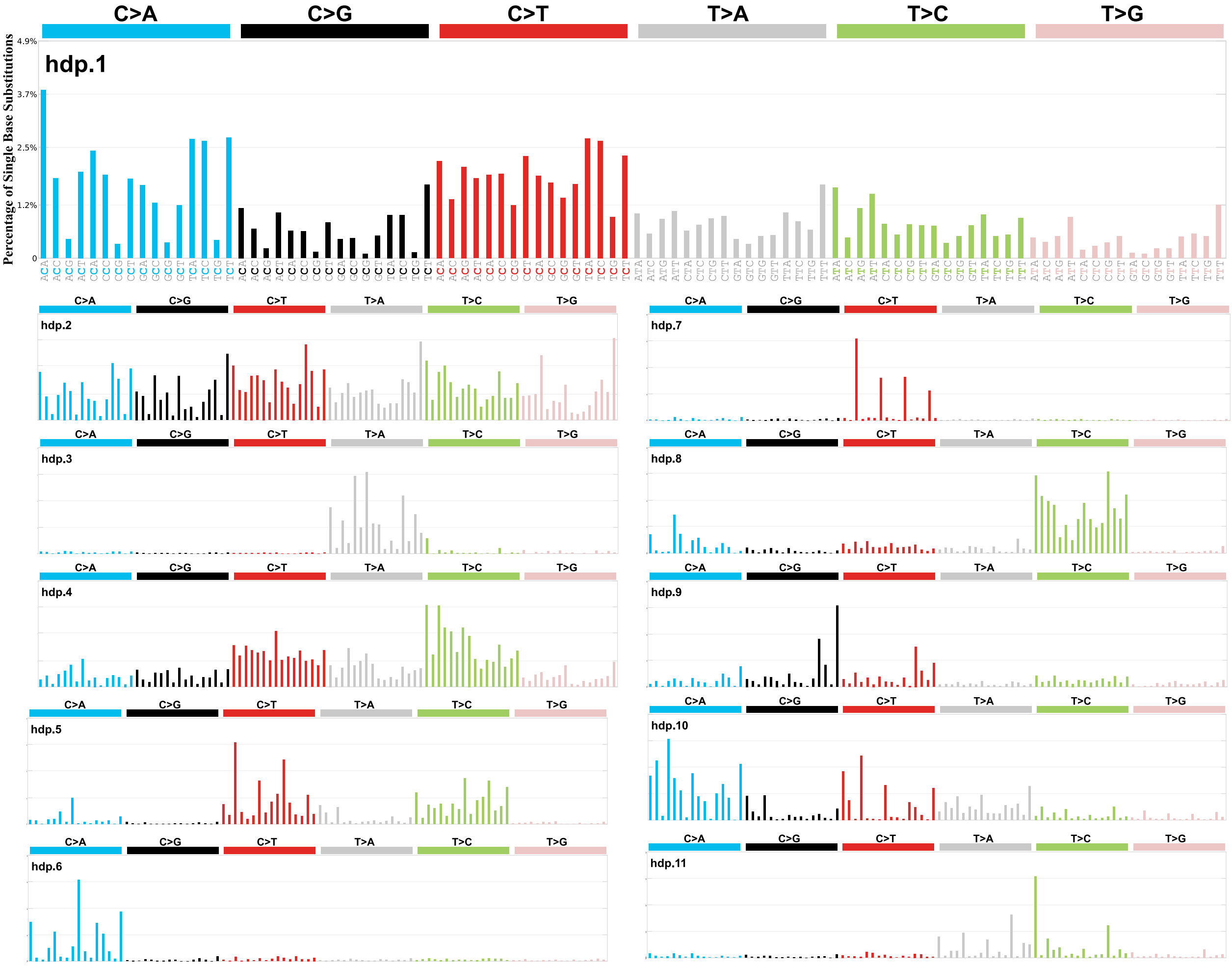

### Supplementary Fig.6: Small insertion and deletion mutational signatures extracted by mSigHdp

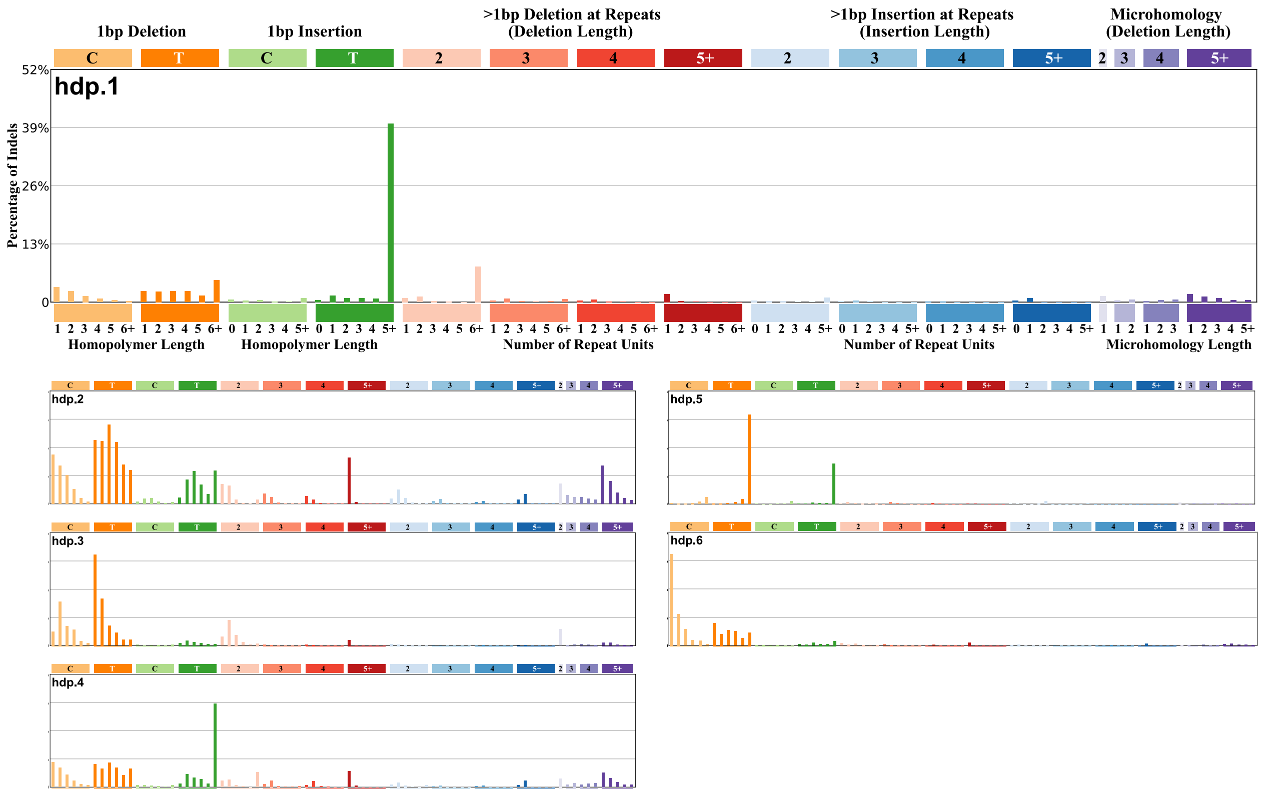

### Supplementary Fig.7: Reconstruction of COSMIC reference signature SBS40

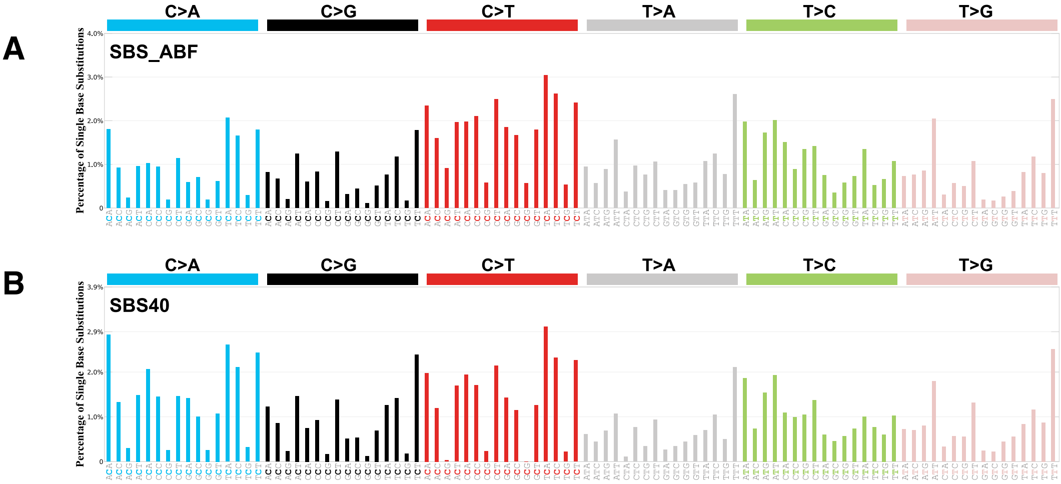

### Supplementary Fig.8: Comparison of signatures previously extracted from ccRCC

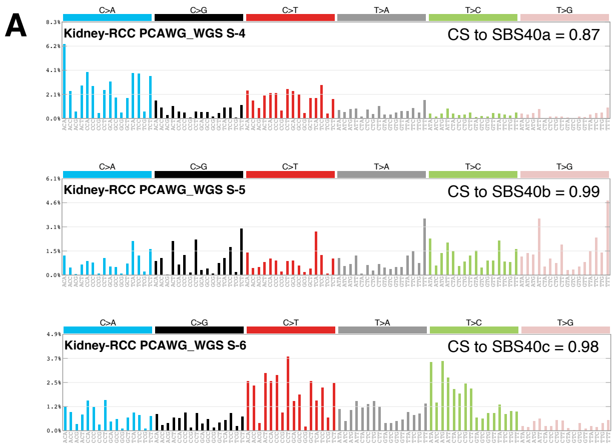


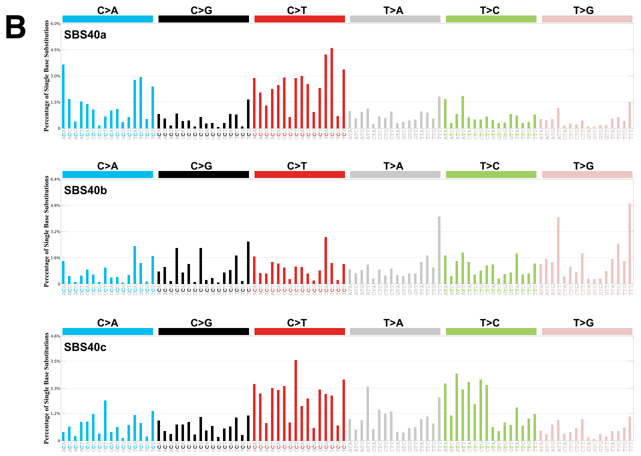

### Supplementary Fig.9: Aristolochic acid mutational signatures in kidney cancers

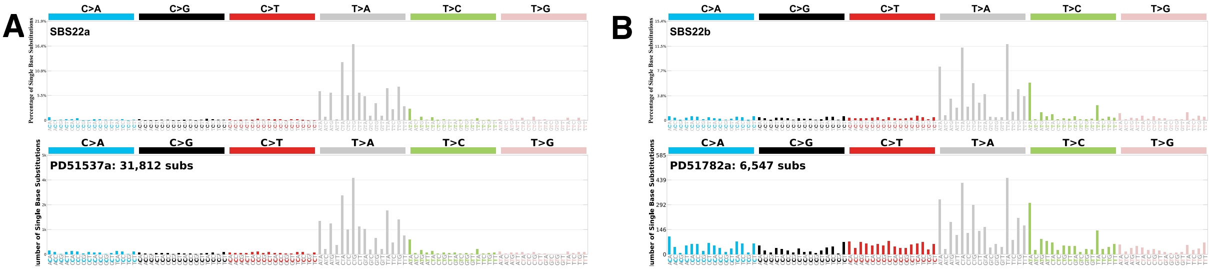

#### Supplementary Fig.10: Presence of tobacco-associated signature SBS4 in in kidney cancers

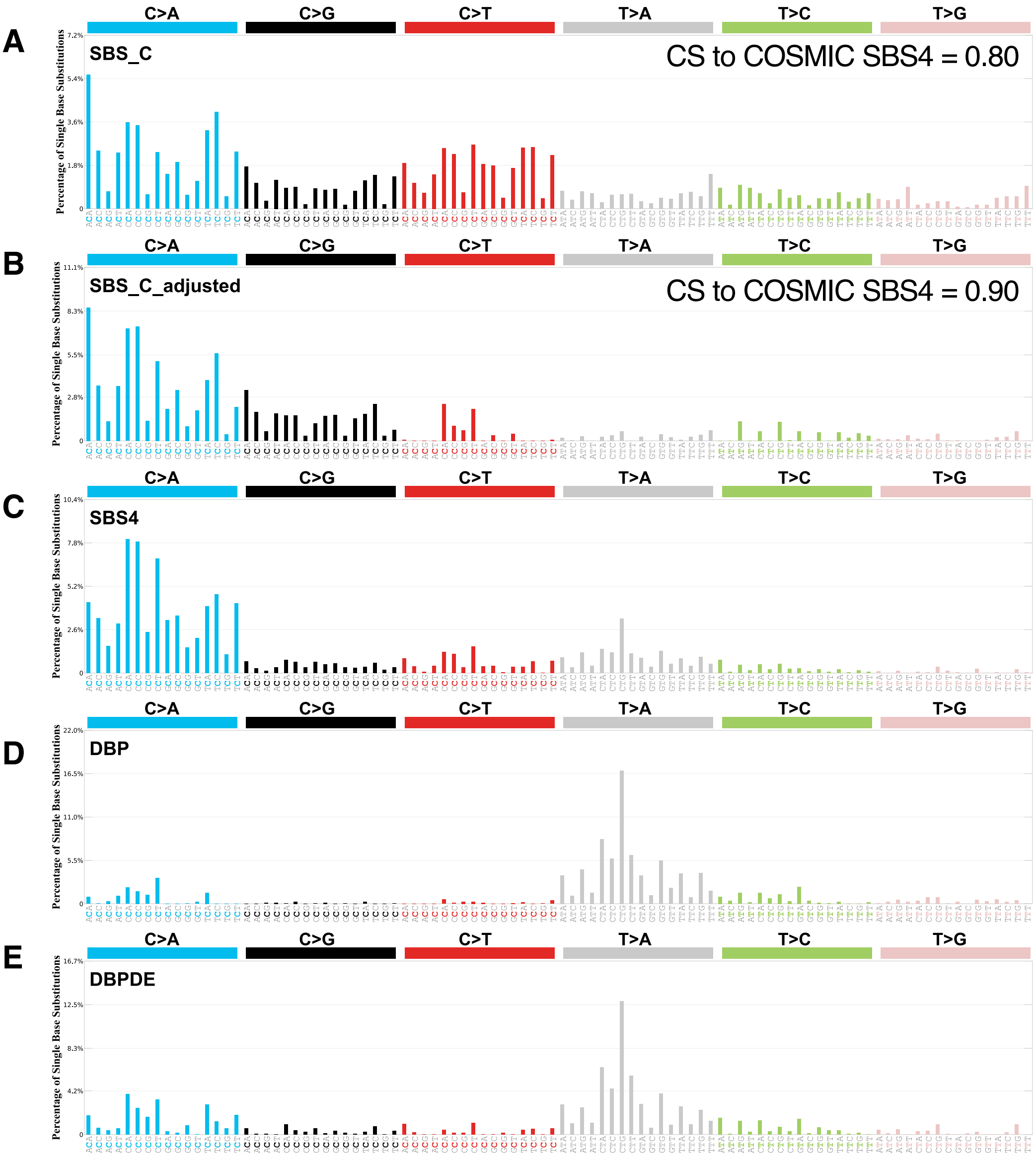

#### Supplementary Fig.11: Attribution of signature SBS12 in liver cancers

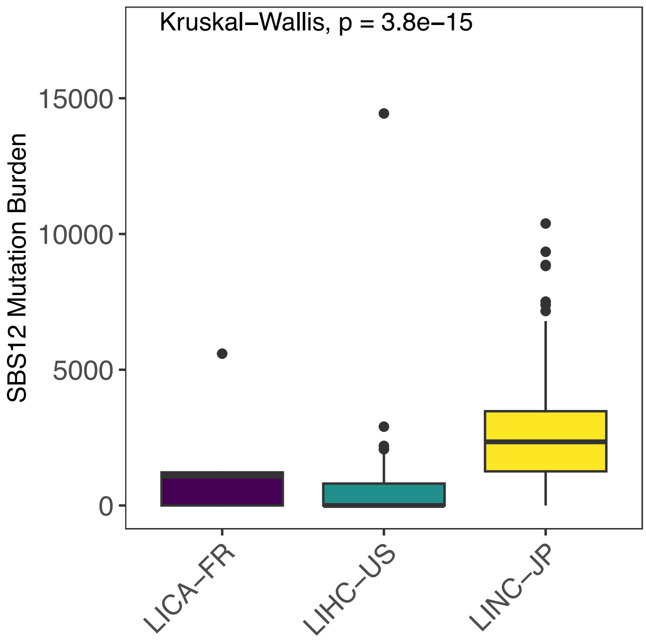

### Supplementary Fig.12: Attribution of signature SBS12 in validation cohorts

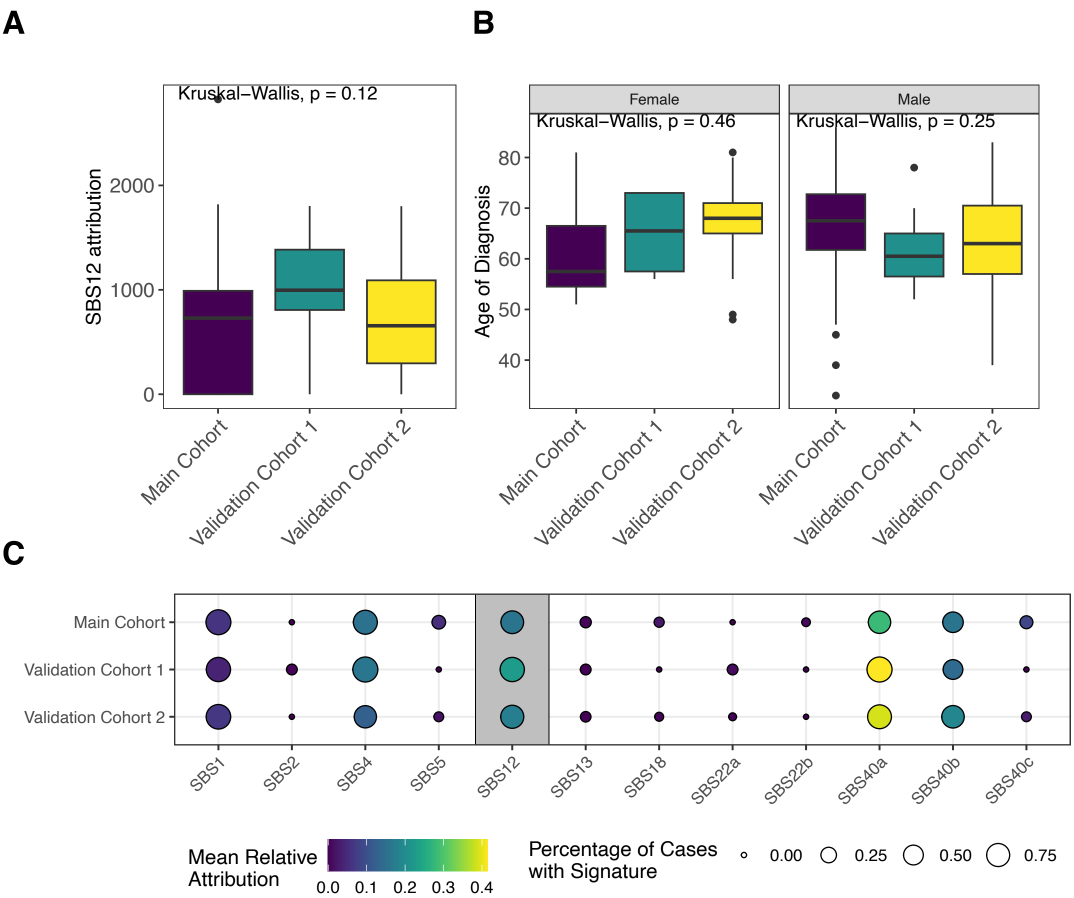

### Supplementary Fig.13: Difference between unadjusted and adjusted tumour mutation burdens across countries

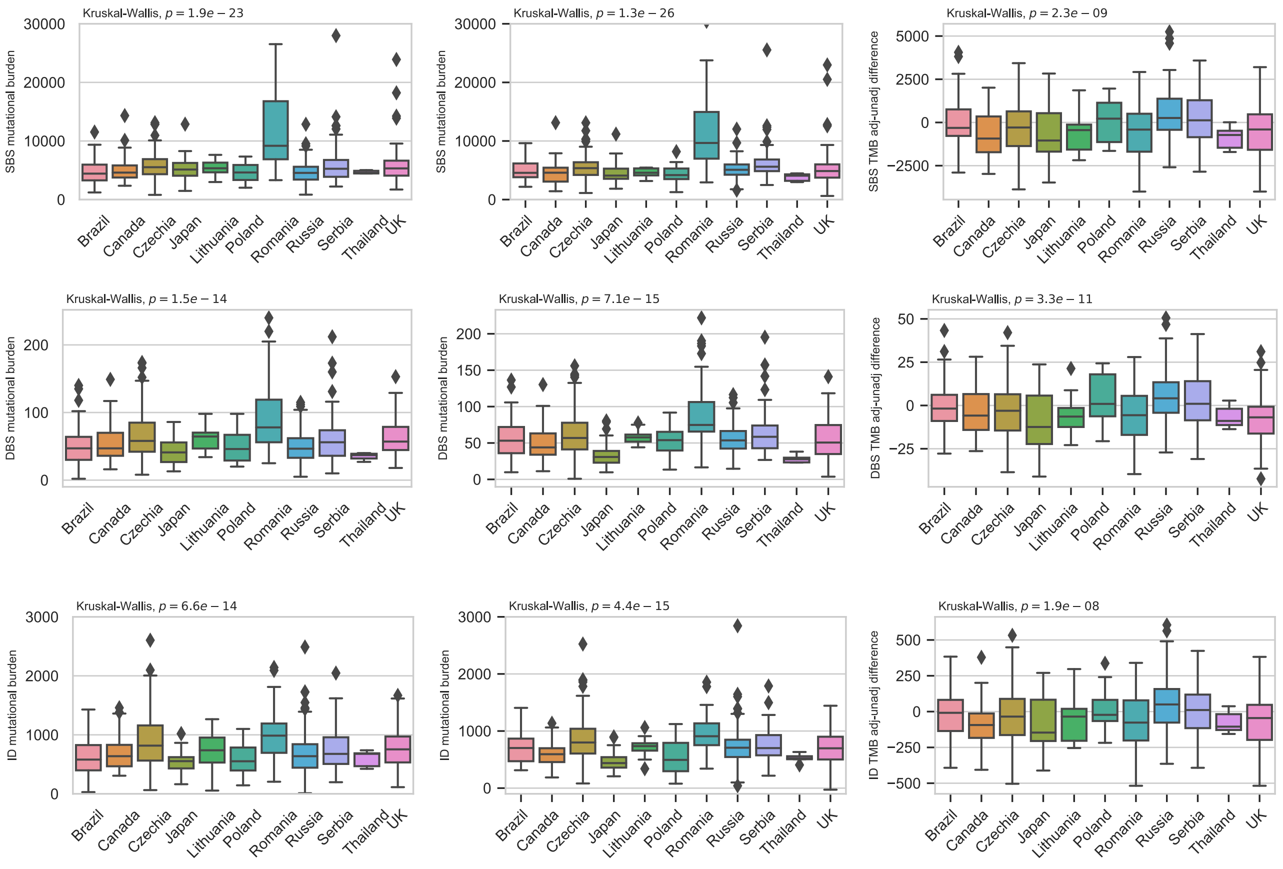

#### Supplementary Fig.14: Associations of relative attributions of mutational signatures with incidence of kidney cancer

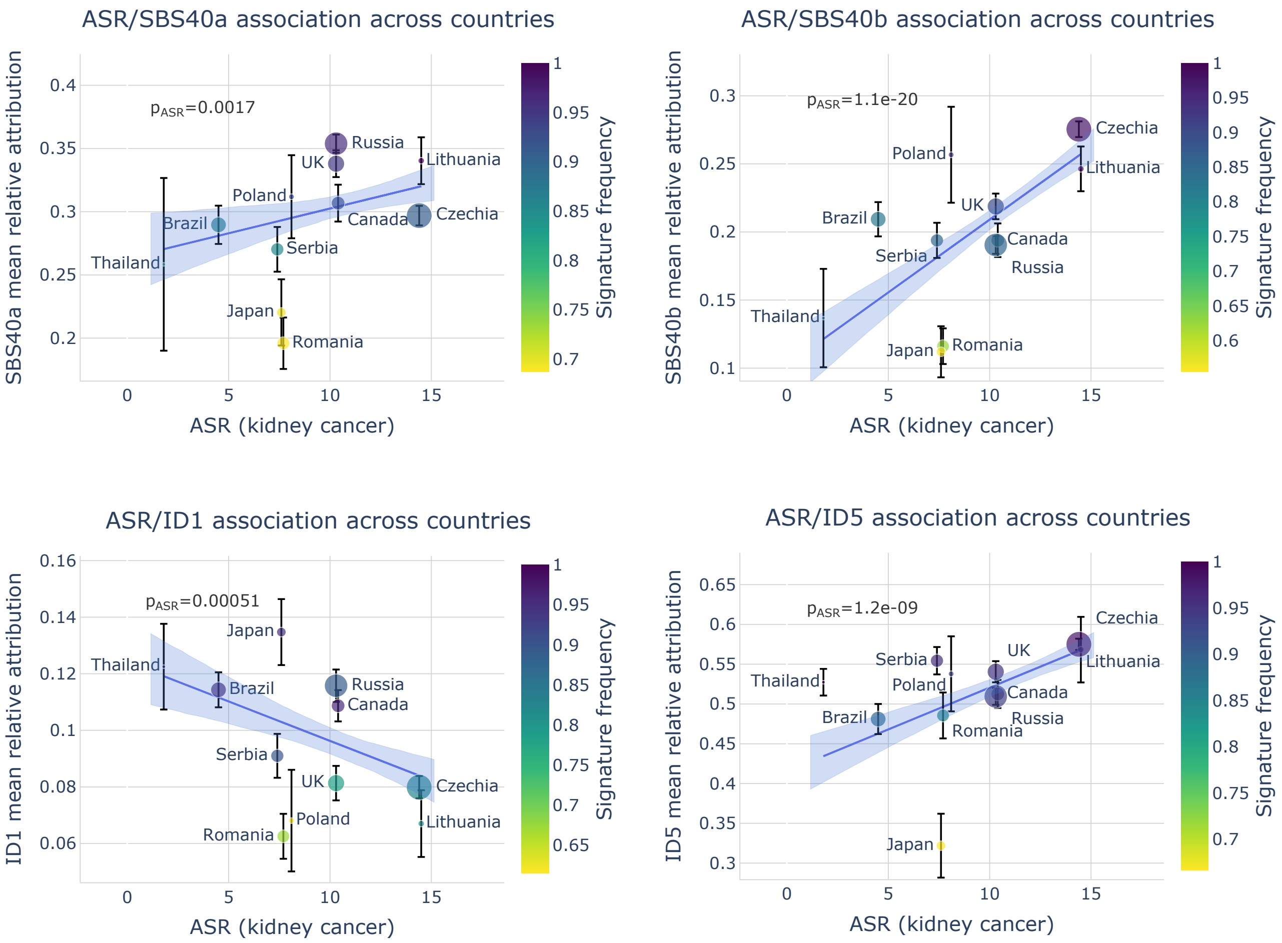

#### Supplementary Fig.15: Associations of total mutation with incidence of kidney cancer excluding countries with predominant AA signatures

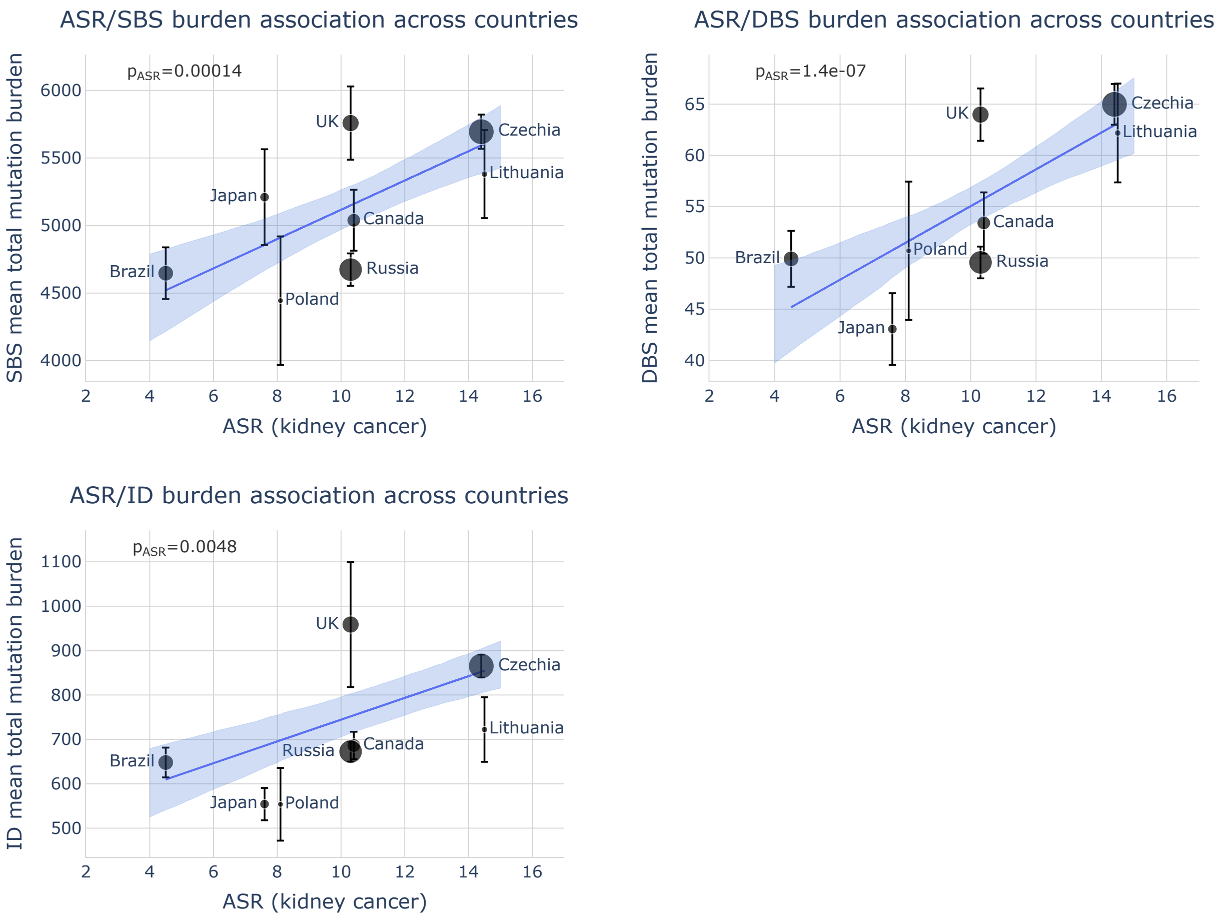

### Supplementary Fig.16: Random signature profiles for SBS and ID mutation types, generated using Poisson distribution

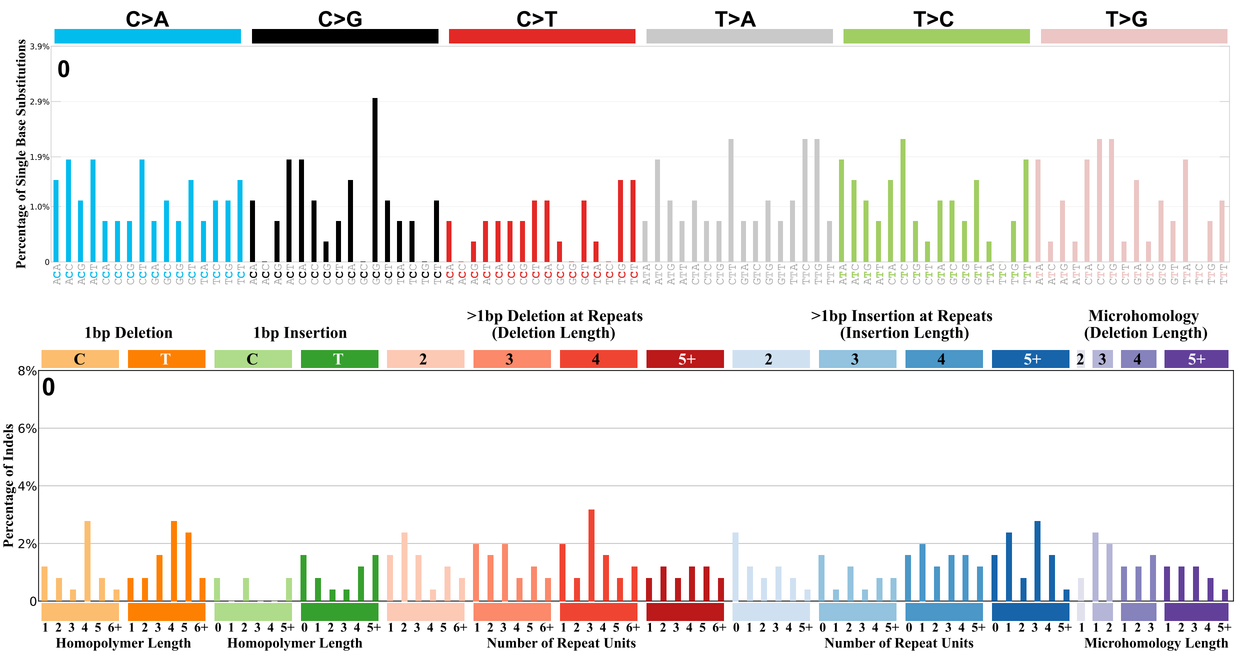

#### Supplementary Fig.17: Regression of permuted signature attributions and burdens with ASR

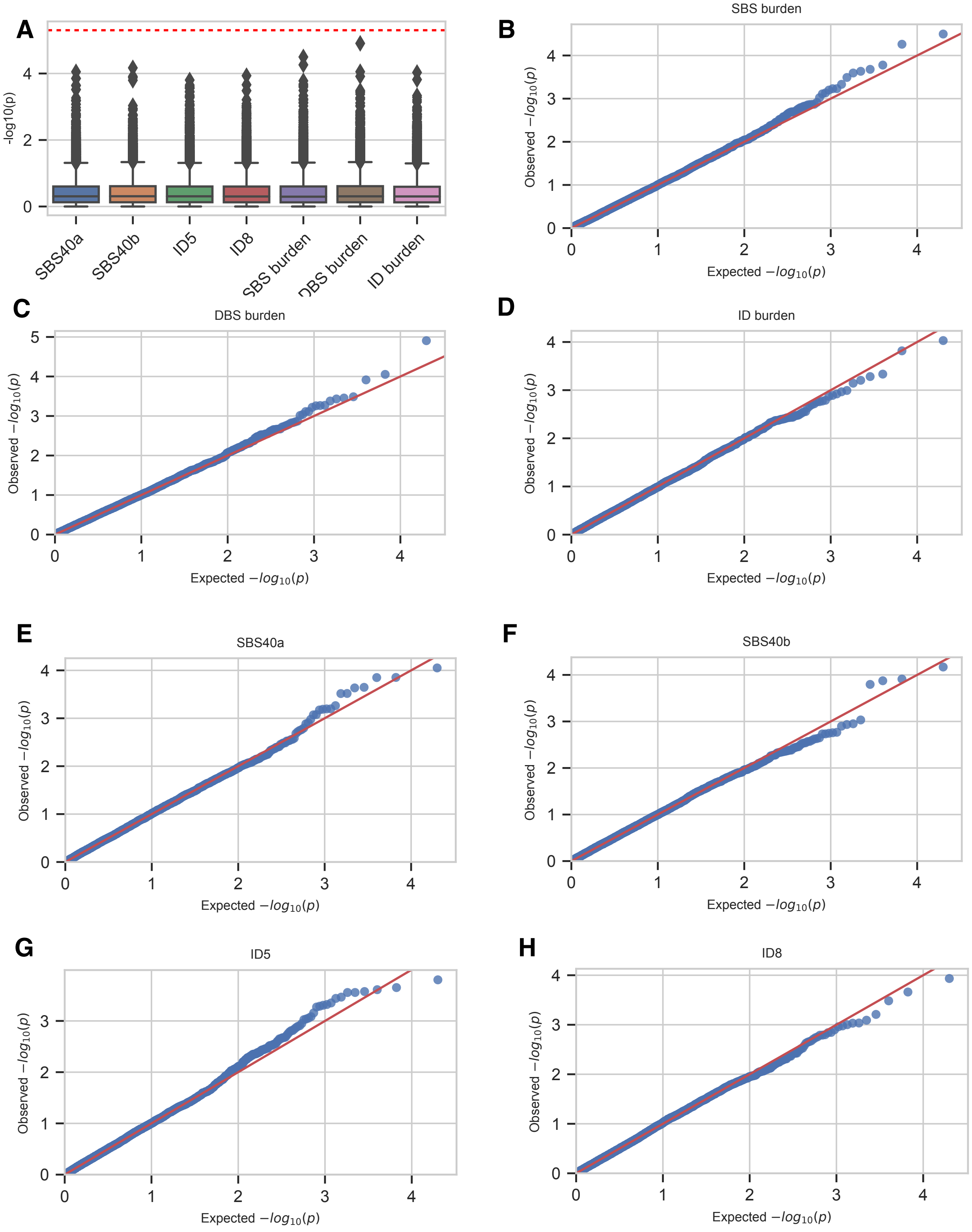

### Supplementary Fig.18: Copy number signatures extracted by SigProfilerExtractor

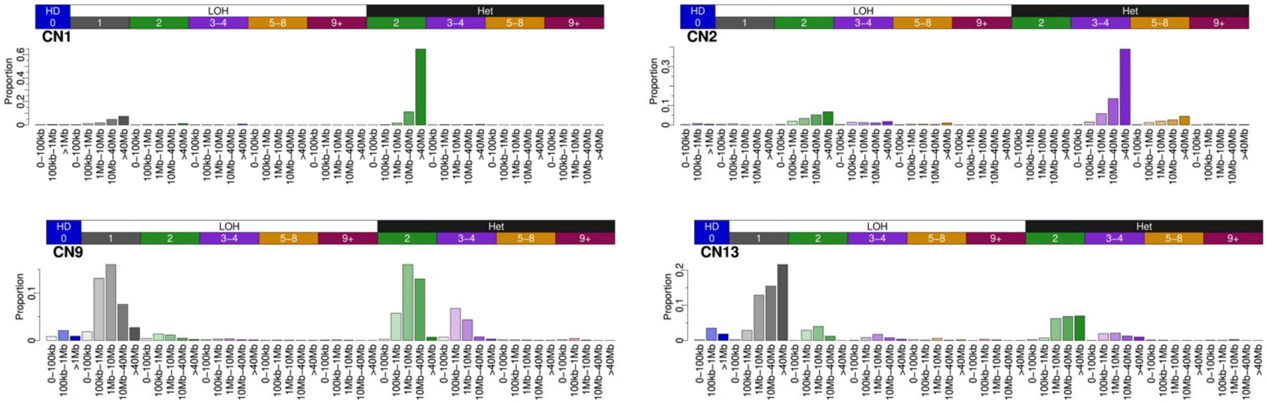

### Supplementary Fig.19: Structural rearrangement signatures extracted by SigProfilerExtractor

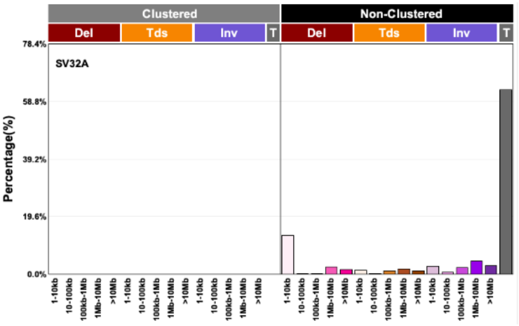


Non-clustered translocations

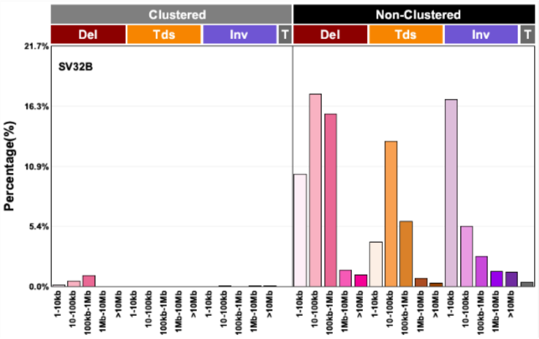


Small non-clustered events

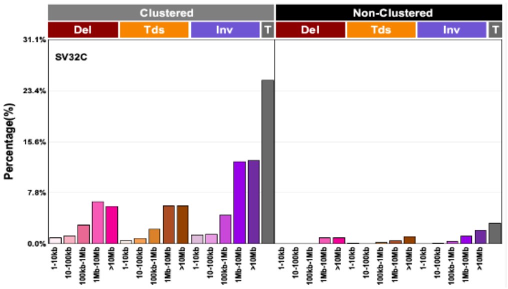


Clustered events

#### Supplementary Fig.20: Associations of copy number and structural variant signatures with incidence of kidney cancer.

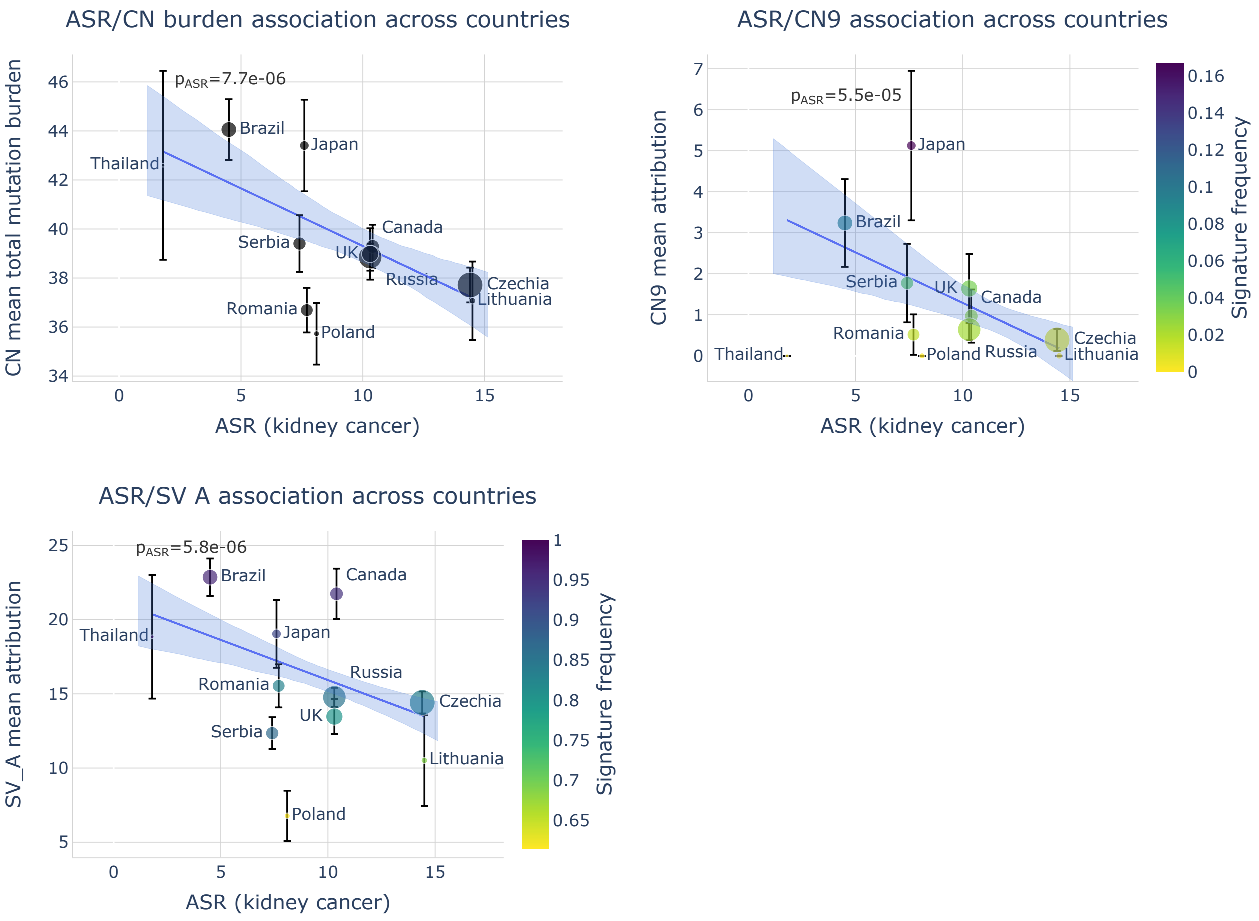

#### Supplementary Fig.21: Polygenic risk scores

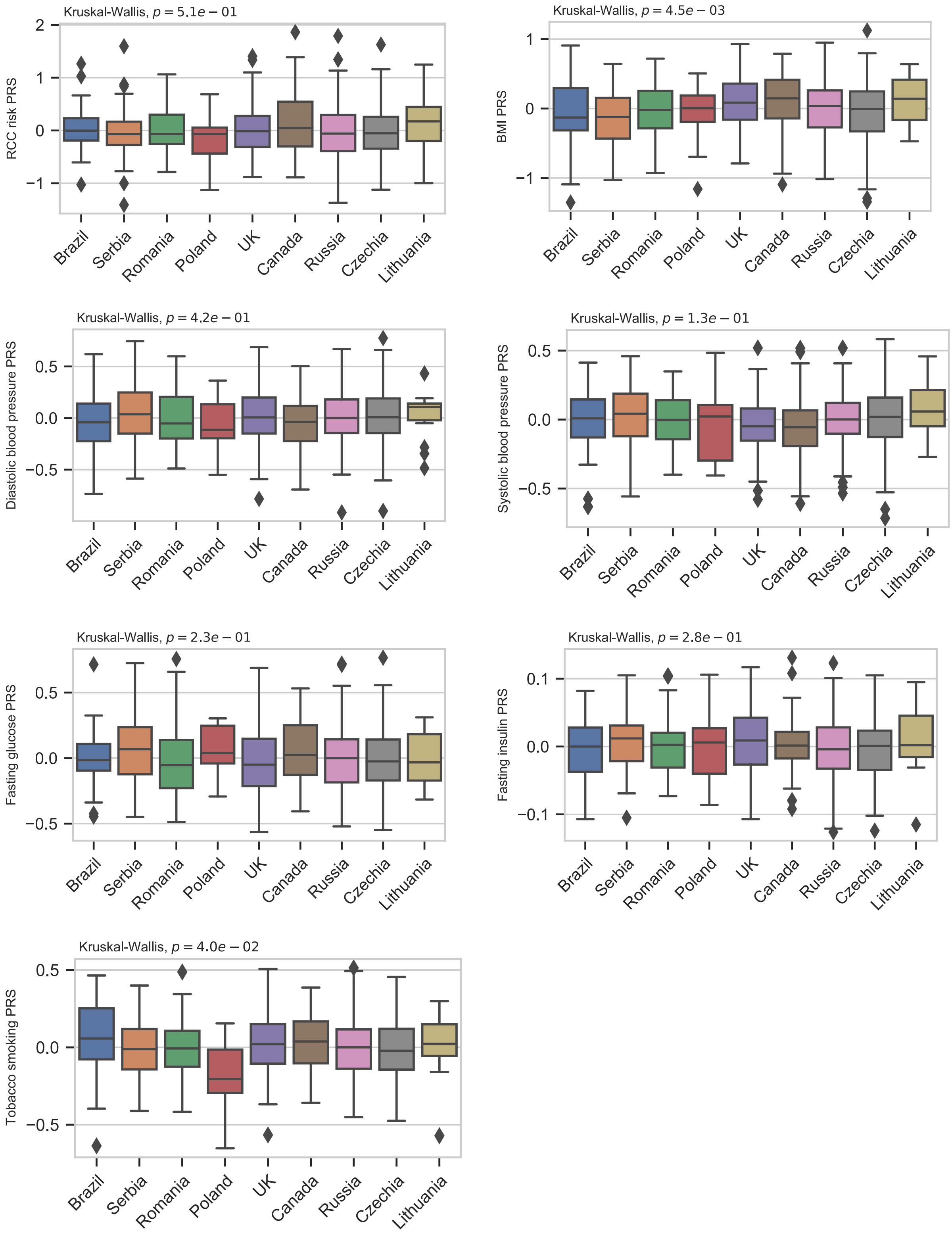

#### Supplementary Fig.22: Log2-normalized intensity distribution in QC samples

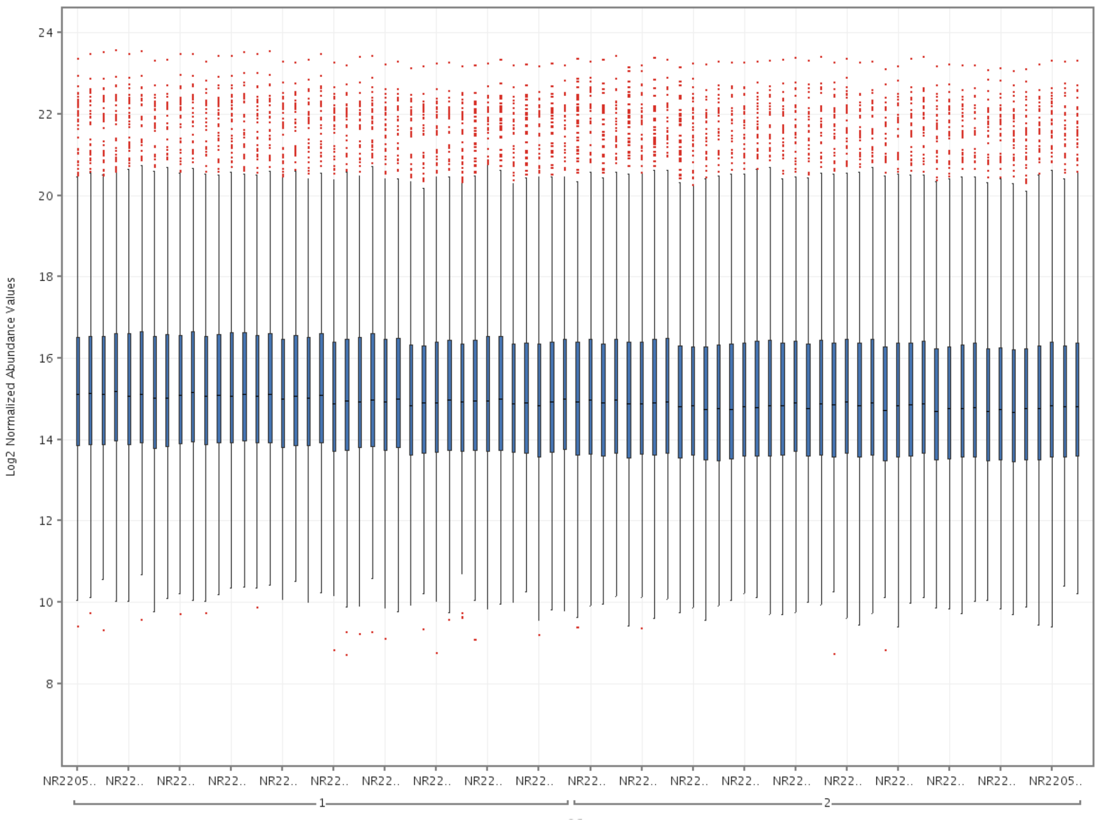

**Supplementary Fig.23: Intensity variability in QC samples**

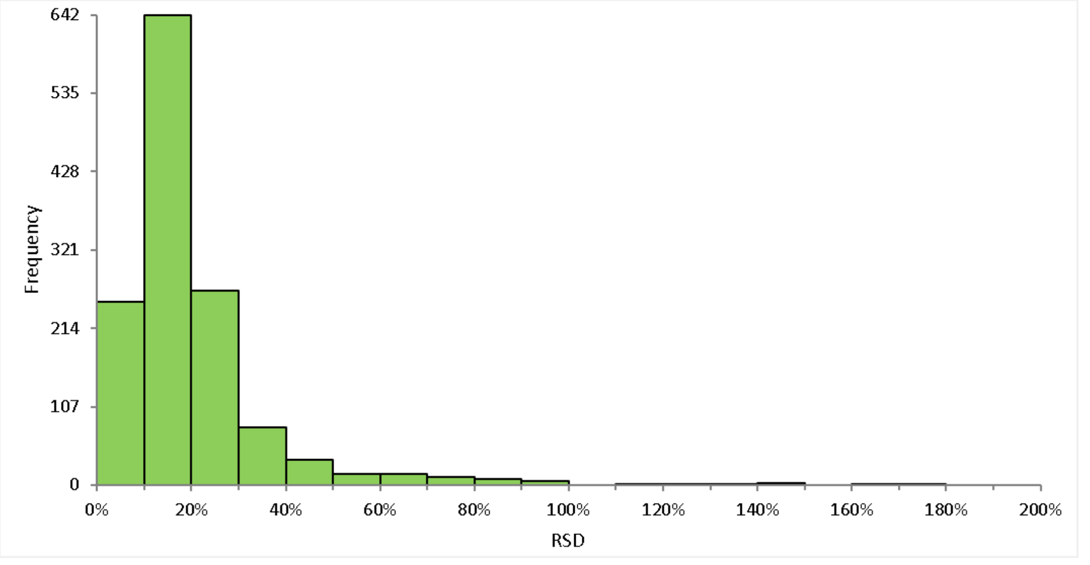

#### Supplementary Fig.24: Number of reads per tumor chromosomal copy (NRPCC) with respect to the total number of SNVs

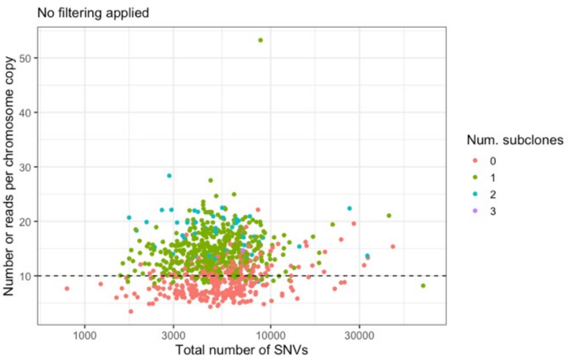


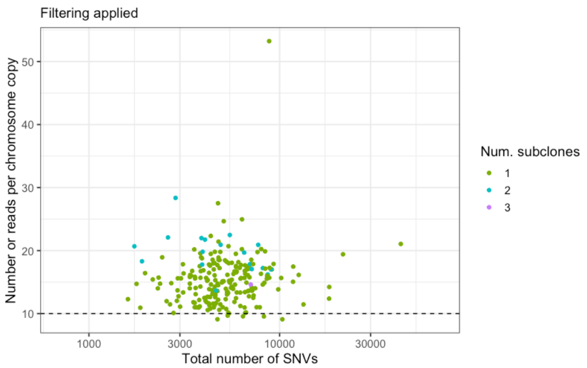

**Supplementary Fig.25: Number of reads per tumor chromosomal copy by filtering status**

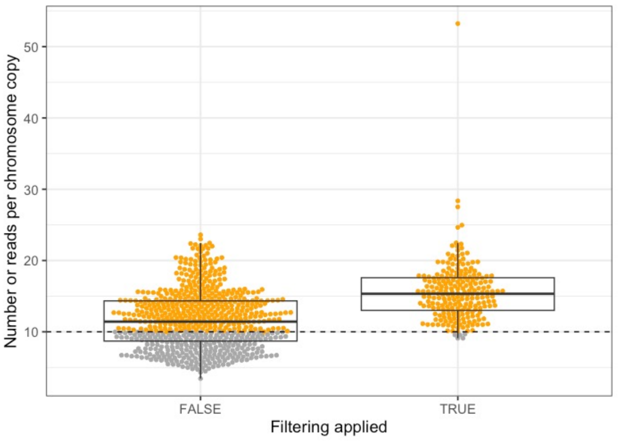

### Supplementary Fig.26: Number of reads per tumor chromosomal copy with respect to the minimum CCF of detected clusters

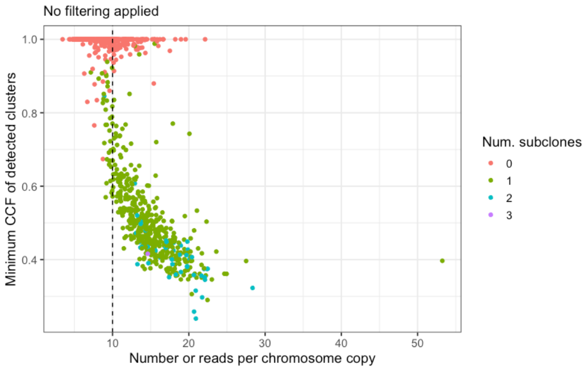


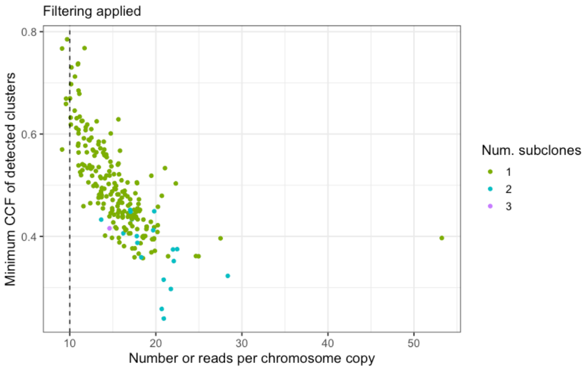
